# REMI: A Free Tool for Truncated Proximity Searching in PubMed

**DOI:** 10.64898/2026.11.19.26368485

**Authors:** Remigius van Alebeek, Rinus Verdonschot

## Abstract

**Background:** PubMed is the most comprehensive freely available database for medical and health sciences publications, making it an essential platform for literature searching in budget-constrained settings (e.g. non-academic hospitals). However, unlike subscription platforms (e.g. Ovid MEDLINE), it cannot combine truncation with proximity operators. Searchers must therefore either enumerate every word form manually, risking missed variants, or fall back on Boolean AND, which retrieves many irrelevant records. We developed REMI (Related Expression Machine Identification), a free, open-source web application which emulates truncated proximity searching in PubMed.

**Case Presentation:** REMI was developed at Maastricht University Library. Through the NCBI E-utilities, it identifies the variants of truncated terms that occur in PubMed records. An iterative exclusion procedure, which reruns the search for each concept while excluding that concept’s variants already found, surfaces progressively rarer forms. Users select relevant variants from frequency-ranked lists, and REMI combines them into native PubMed proximity clauses that can be documented, peer reviewed, and rerun without REMI. Performance of REMI was evaluated by comparison to four Ovid MEDLINE search strategies from published reviews. REMI’s proximity search was able to identify all included articles found with the original Ovid MEDLINE query.

**Conclusions:** REMI makes truncated proximity searching available in PubMed while keeping searches transparent, reliable, and reproducible, which particularly benefits searchers without access to subscription platforms. Because variant discovery is a sampling procedure, completeness is not guaranteed, and long searches require variant selection. Other libraries can use, host, or adapt the tool, however, native support in PubMed would remain the preferable solution.

## 1 Background

PubMed is a widely used bibliographic database for retrieving literature in the biomedical and health sciences [1]. Although much of its coverage comprises citations indexed in MEDLINE, PubMed also contains additional records, including those from PubMed Central [2]. Searching PubMed or MEDLINE is considered an essential component of systematic reviews in the health sciences and provides excellent coverage within the biomedical domain [3, 4, 5]. Compared with MEDLINE accessed through subscription-based platforms, PubMed offers several advantages: it is freely available worldwide, provides a user-friendly interface, and facilitates flexible searching through Automatic Term Mapping [6]. Nevertheless, its search interface has a notable limitation: unlike many other bibliographic databases and subscription-based MEDLINE platforms, PubMed does not permit the simultaneous use of truncation and proximity operators [7].

Proximity searching is an alternative searching method compared to utilizing the Boolean AND. With the Boolean AND, terms entered and combined in such a way may appear at any locations within the defined field, which may result in the terms occurring in unrelated contexts. With proximity searching, a predefined distance can be set for these terms. This improves precision and reduces the workload during screening. Combining proximity searching with truncation can support this balance further. Truncation retrieves multiple word forms sharing a common stem (e.g. plural and singular forms), without requiring each variant to be specified individually. When used within a proximity search, it allows these variants to be retrieved while retaining the specified distance constraint.

However, as previously indicated, PubMed does not support truncation within proximity searches: if a wildcard is included, the proximity operator is ignored [7]. Users must therefore explicitly and exhaustively combine the required word forms and combine separate proximity expressions with Boolean OR. As the number of variants increases, constructing these queries manually becomes more laborious and prone to omissions (such as uncommon inflections and spelling variants), potentially reducing sensitivity. Therefore, Boolean AND remains useful when maximizing sensitivity takes priority. However, as the published literature continues to expand, proximity searching is increasingly valuable for reducing the number of records requiring screening while maintaining adequate sensitivity [8].

Support for truncation within proximity searches could therefore improve the efficiency of systematic searching in PubMed. Although existing tools can generate proximity queries from user-specified terms [9], a need remains for functionality that more closely reproduces the behavior of proximity searches with truncation. To address this need, we developed REMI, a transparent and user-friendly tool that generates PubMed queries designed to emulate proximity searches with truncation. Here, we describe REMI and report the results of its validation.

## 2 Case Presentation

### 2.1 REMI - Info and Purpose

REMI (Related Expression Machine Identification; current version 2.1) is a free, open-source web application developed at Maastricht University Library to support systematic searching in PubMed. Its purpose is to emulate a truncated proximity search, such as vaccin* adj3 hesitan* in Ovid MEDLINE, by translating it into an equivalent set of proximity clauses written in PubMed syntax. To do so, REMI first discovers which variants from a truncated term are found in PubMed records and then combines these variants into proximity clauses that PubMed can execute.

REMI offers two functions, each on its own page. The function “Keyword Search” takes a single truncated term or phrase and lists the variants that the truncation retrieves in the titles and abstracts of PubMed records, ranked by frequency; selected variants can be copied or exported to a CSV file. This function can be used, for example to judge whether a truncation point is too broad or too narrow. The function “Proximity Search”, the core of the tool, takes two concepts, each entered in a separate field as one or more terms, truncated or not, separated by OR (e.g., `hesitan* OR reluctan*`). Terms can also be phrases, entered with quotation marks or a hyphen (e.g., `“hand off*”` or `hand-off*`). REMI identifies the variants of each truncated term that occur in records in which both concepts are present, presents the variant list to the user for selection, and generates a PubMed search string in which every selected variant of the first concept is combined with every selected variant of the second at a user-specified proximity distance (1 to 99 words; default 3). Further options restrict variant discovery to a publication date range, control the handling of hyphenated words, and enable a more exhaustive Precise Proximity mode, described below. The date range applies to variant discovery only and is not added to the final generated search, so the same limits must be applied in PubMed (if necessary). Input is restricted to letters, digits, spaces, hyphens, quotation marks, and the asterisk. As in PubMed, the asterisk can be used at the end of a term, in the middle (e.g., `randomi*ation`), or in several words of a phrase, and at least four characters must precede the first wildcard [6].

REMI does not execute the final search itself. Its output is an ordinary PubMed query that the user pastes into PubMed, and that can therefore be documented, peer reviewed, and rerun by others without REMI. The tool is available as a hosted web application at https://github.com/MaastrichtU-Library/remi, and the source code is released under the GNU Affero General Public License v3.0 at https://github.com/MaastrichtU-Library/remi, together with step-by-step instructions for running a local copy with a personal NCBI application programming interface (API) key. No programming experience or user account is required.

### 2.2 REMI - Methodological Backbone

REMI rests on a logical property of proximity searching: a record can be retrieved by a proximity search only if both concepts occur somewhere in its title or abstract. Every record that a truncated proximity search would retrieve is therefore also retrieved when the two truncated concepts are combined with Boolean AND, a search that PubMed can execute. Any variant that could contribute to a proximity hit must consequently occur in this co-occurrence set. Variants of a stem that never co-occur with the other concept cannot produce a proximity hit and would only lengthen the final search string. For example, `vaccin*` also matches *vaccinium*, the plant genus that includes cranberries, but unless *vaccinium* occurs in a record that also contains a word beginning with *hesitan*, it never enters REMI’s variant list. REMI therefore mines variants exclusively from the co-occurrence set, using an iterative exclusion procedure (Figure 1; Appendix A, Table S1).

**Figure 1:**
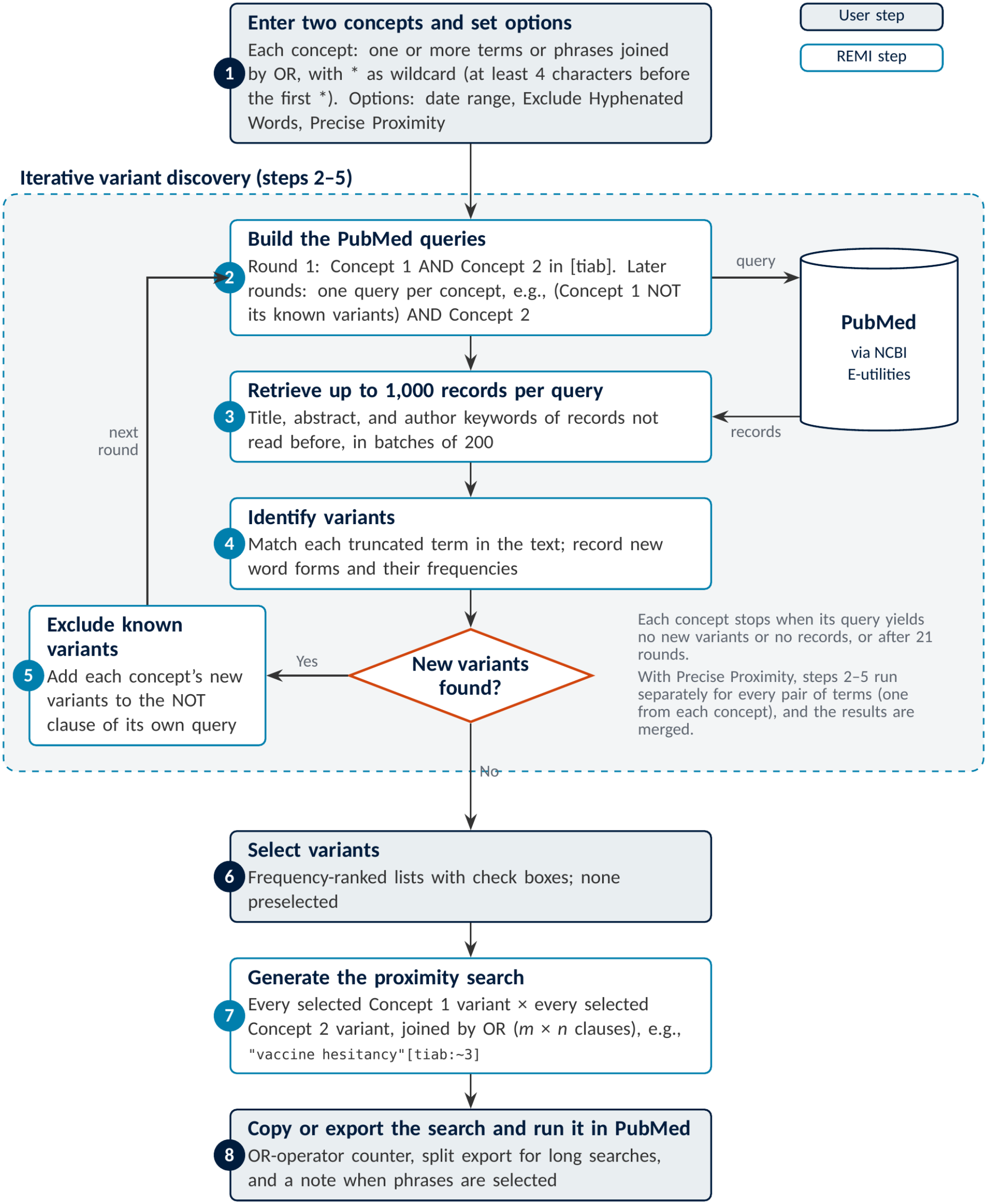
Workflow of REMI 2.1 Proximity Search. Gray boxes are steps performed by the user; white boxes are performed automatically by REMI, which queries PubMed through the NCBI E-utilities.

First, each term is tagged with the title/abstract field (`[tiab]`), terms within a concept are combined with OR, and the two concepts are combined with AND. REMI submits this query to PubMed through the NCBI Entrez Programming Utilities (E-utilities) [10], retrieves up to 1,000 PubMed identifiers (PMIDs), and downloads the corresponding records in batches of 200. For each record, the title, the abstract (including the section labels of structured abstracts), and the author keywords are combined into a single lower-case text.

Second, each truncated term is converted into a regular expression in which every asterisk matches any sequence of letters, digits, or hyphens and the match must begin at the start of a word, mirroring the PubMed wildcard, which substitutes for zero or more characters [6]. In phrases, the words may be separated by a space or a hyphen, so `“hand off*”` matches both *hand off* and *hand-offs*. Because both spellings retrieved the same records in PubMed in our tests, phrase variants are always recorded with a hyphen and their frequencies combined. Every distinct form matched in the downloaded text is recorded as a variant of the term, together with the number of times it occurs.

Third, the search is repeated with the variants found so far excluded with NOT. From the second round on, REMI runs one query per concept, each excluding only that concept’s known variants (Appendix A, Table S1). The next batch therefore consists of records in which that concept matched only through forms REMI has not yet seen. Excluding the known variants of both concepts at once would also skip every record that combines a new variant of one concept with a known variant of the other. Records that REMI has already read are not downloaded again. Successive rounds thereby reach progressively rarer variants without REMI having to download the entire co-occurrence set, which for common concepts can comprise many thousands of records. For each concept, iteration stops when a round yields no new variants or the query returns no records, with a maximum of 21 rounds (10 rounds in Keyword Search). Terms entered without truncation are carried into the final search unchanged and are not used for variant discovery or exclusion.

Two options modify this procedure, the “Exclude Hyphenated Words option” and the “Precise Proximity” option. The Exclude Hyphenated Words option, which is enabled by default, does not list hyphenated forms (e.g., *vaccine-related*) as separate variants; instead, the segment preceding the hyphen is recorded as a variant if it has not already been found. In our tests, PubMed treated the parts of a hyphenated word as separate words within a proximity clause, so a hyphenated variant does not restrict the search to the hyphenated form, and folding it into its first segment keeps the variant lists short. Phrases entered by the user, such as `hand-off*` , keep their hyphen. When the option is disabled, hyphenated forms are listed as separate variants and labeled as hyphenated. The Precise Proximity option addresses a different issue. When a concept contains several terms, the most frequent terms can dominate the 1,000 records retrieved in each round, leaving less frequent terms underexplored. Precise Proximity therefore runs the complete procedure separately for every pairing of a term from the first concept with a term from the second and merges the results, with a progress bar showing the number of completed pairings. It also gives every term its own exclusion list, whereas in the default mode all terms of a concept share one. This is more exhaustive but multiplies the number of PubMed requests and, therefore, the run time.

After discovery, REMI lists the variants of each truncated term, ranked by frequency, each with a check box for selection by the user. These frequencies reflect occurrences in the records REMI sampled, not in PubMed as a whole, and are intended to help users judge relevance. No variant is selected by default: users select the variants to include, individually or with select-all buttons per term or for all terms, and leave out, for example, unrelated words that happen to share the stem. REMI then combines each selected variant of the first concept with each selected variant of the second into a clause such as `“vaccine hesitancy”[tiab:∼3]` , in which the number specifies the maximum number of intervening words, and joins all clauses with OR. Because PubMed proximity searching ignores word order [7], one clause per pair suffices; the final string contains *m × n* clauses for *m* and *n* selected variants.

The generated search is displayed in a text box and can be copied or exported as a .txt or .doc file. Because the number of clauses can increase rapidly, REMI displays a running count of OR operators and issues a warning when this count exceeds 2,048, beyond which PubMed rejects the query. PubMed also rejects queries containing more than 200,000 characters (Appendix B, Figure S1 and S2).

During export, searches exceeding the operator limit are divided into consecutive subqueries, each within the 2,048 operator limit. However, running these subqueries separately and subsequently combining them in PubMed’s search history (e.g., `#1 OR #2`) does not circumvent this restriction: the combined search will still fail if it exceeds the operator limit. Users should therefore select term variants so that the complete search remains within PubMed’s operator and character limits.

When a selected variant is a phrase or a hyphenated word, REMI also displays a note explaining that PubMed searches the words of a phrase separately within a proximity clause, illustrated with a clause from the user’s own search.

REMI was designed to use the E-utilities responsibly. Requests are made with an NCBI API key, spaced at least one second apart within a user session, globally throttled to stay within NCBI’s rate limits, and retried with exponential back-off when they fail.

### 2.3 REMI - Validation

To assess how accurately REMI reproduces conventional proximity search results in databases such as MEDLINE, four validation searches were conducted for both the Keyword Search and the Proximity Search function. The Keyword Search validation was performed on 15 October 2026, while the Proximity Search functionality was validated on 19 October 2026. Both validations made use of REMI version 2.1.

The Keyword Search validation used four truncated terms: `tumo*, polic*, health*` , and `semaglut*`. Each term was first searched within PubMed individually to establish a reference hit count. REMI’s Keyword Search function was then used to identify variants of each term. These variants were combined using the Boolean operator OR, and the resulting hit count was compared with that of the corresponding truncation search. Overall, REMI Keyword Searches showed nearly 100% agreement with regular truncation searches in terms of hit counts. Although complete agreement would be expected, two factors account for the observed discrepancies. First, REMI’s variant retrieval does not include terms appearing in the “Other Abstract” (plain language summary). Second, PubMed occasionally retrieves hyphenated terms that do not match the intended truncation pattern; for example, *tumo*[tiab]* retrieved an article containing *“TU-modified”* in the abstract [11]. Such discrepancies reflect how PubMed indexes hyphenated words rather than shortcomings of REMI. Results for individual terms are presented in Table 1, and the complete queries generated by the Keyword Search function are provided in Appendix C. No date-limitations were used, and *Exclude hyphenated words* option was enabled.

**Table 1:**
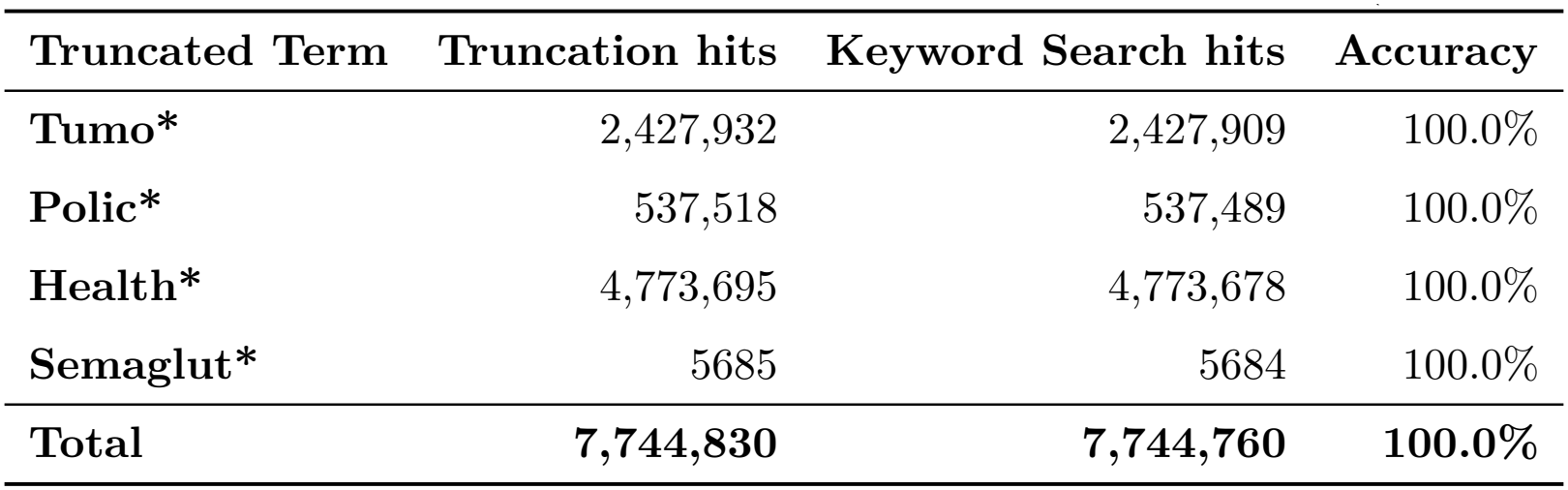
Comparison of truncation hits and REMI 2.1 keyword search hits (15 October 2026)

To validate the “Proximity Search” function, its ability to retrieve articles included in four published systematic reviews and retrieved by their original Ovid MEDLINE search strategy was assessed. [12, 13, 14, 15]. To enable a consistent comparison, the PubMed proximity query generated by REMI was translated into Ovid MEDLINE syntax while preserving the variant pairs generated by REMI. For proximity searches which would exceed 2,048 Boolean OR operators, a selection of variants was performed. Selection of variants was based on frequency and suitability to the context, as determined by one of the authors. Retrieval of included articles in the corresponding reviews was assessed only for articles that had a Ovid MEDLINE record which was also retrieved by the original proximity search.

REMI retrieved no unique articles relative to the original Ovid MEDLINE search, except for 8 articles in the first validation search. This discrepancy arose from differences in how REMI handles proximity within phrases. Specifically, REMI applies proximity operators between individual words within a phrase as well as between the phrase and other search terms. For example, the Ovid MEDLINE query *“vaccine-injection* ADJ4 hesitanc*”* is translated by REMI into a PubMed query equivalent to *“vaccine ADJ4 injection* ADJ4 hesitanc*”*, because PubMed does not support exact phrase matching within proximity searches [7]. Across all four validations, the queries generated by REMI retrieved all included articles that met these criteria (Table 2).

**Table 2:**
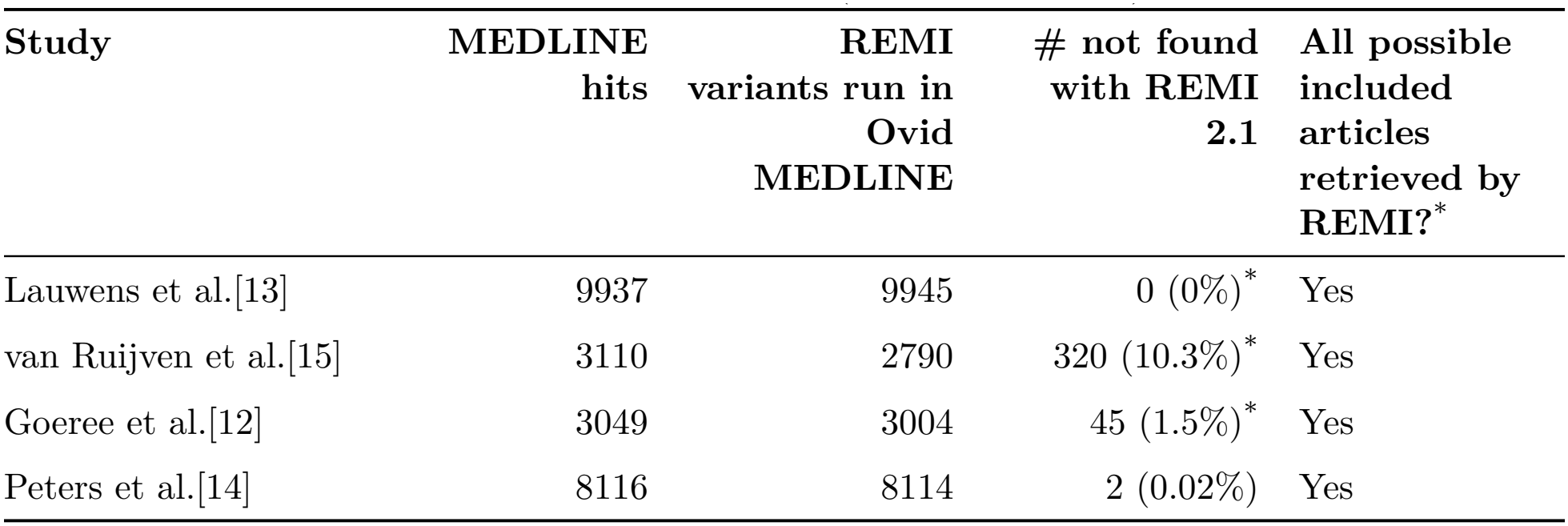
Comparison of Ovid MEDLINE and REMI 2.1 search results. Articles were only considered eligible if the original Ovid MEDLINE query retrieved them as well. *For these searches, variant selection had to be performed. (19 October 2026)

The complete search strategies, including selected and unselected terms and, where applicable, the articles retrieved, are provided in Appendix D. No date limits were applied. The *Exclude hyphenated words* option was enabled, and the *Precise proximity* option was disabled.

## 3 Discussion

### 3.1 Accomplishments

REMI makes a search technique that PubMed does not support natively usable in practice, while keeping the final search entirely within PubMed’s own syntax. This design choice has several consequences. Because REMI produces a standard query, the resulting strategy can be reported in full, as PRISMA-S requires [16]. It can be assessed by a peer reviewer, and rerun by anyone with access to PubMed, with or without REMI. The exports of the selected variants with their frequencies and of the generated search further support documentation of how the search was developed. Because variants are taken from the records themselves rather than from the searcher’s memory, REMI reduces the main risk of enumerating variants manually: forms that the searcher did not anticipate, such as uncommon inflections, spelling variants, or hyphenated compounds, are otherwise silently lost. And because variants are mined only from records in which both concepts co-occur, the resulting lists contain only forms that can actually contribute to a proximity hit, which keeps the final search string as short as the data allow.

The frequency-ranked variant lists are also useful beyond proximity searching. They make visible what a truncation actually retrieves, which supports decisions about truncation points in any database and can serve as a teaching aid in search instruction for students and researchers.

Finally, REMI retains the main advantage of PubMed: free, worldwide access. Searchers without subscriptions to platforms that support truncated proximity searching gain a comparable technique at no cost, and the open-source license allows other libraries to inspect, host, and adapt the tool.

### 3.2 Limitations

REMI emulates truncated proximity searching; it does not replicate it exactly. Most importantly, variant discovery is a sampling procedure without a guarantee of completeness. Each round inspects at most 1,000 records per query, and the number of rounds is capped. Moreover, because each query excludes every record that contains an already known variant of the concept being explored, a rare variant that occurs only in records also containing a common variant of the same concept can remain undiscovered. In the default mode this applies across all terms of a concept; Precise Proximity limits it to variants of the same term. Records in which only such a rare variant lies within the specified distance of the other concept may therefore be missed.

Second, REMI identifies variants in the record text returned by the E-utilities, whereas PubMed searches its own index. Differences in how the two handle hyphens, special characters, and individual record fields mean that REMI’s matching approximates, rather than reproduces, PubMed’s wildcard behavior. The reported frequencies, moreover, describe the sampled records rather than PubMed as a whole and should be read as relative indicators only. In addition, because PubMed cannot keep an exact phrase together within a proximity search, phrase variants such as *hand-off* are searched as separate words, which can retrieve some irrelevant records; REMI warns users when a generated search contains such variants.

Third, the number of clauses grows multiplicatively with the number of selected variants. Broad truncations of two common concepts can produce searches with hundreds or thousands of clauses, which are difficult to peer review and may exceed what PubMed accepts in a single query. The OR counter helps users stay within PubMed’s limits (Appendix B), but careful selection of variants remains part of the method rather than an optional extra, and it relies on the searcher’s judgment.

Fourth, the scope of REMI 2.1 is deliberately narrow. It combines exactly two concepts, applies a single distance to all clauses, generates clauses for the title/abstract field only, and accepts only unaccented letters, digits, hyphens, quotation marks, and the asterisk, so terms containing diacritics or other characters cannot be entered. It also depends on the availability of the E-utilities, and run time increases with the size of the co-occurrence set, particularly in precise mode.

Finally, a REMI search is a snapshot. New word forms enter the literature over time, so variant discovery should be repeated when a search is updated rather than the original search string being reused unchanged.

### 3.3 Conclusion and next steps

REMI shows that the absence of truncated proximity searching in PubMed can be largely bridged with publicly available infrastructure. By obtaining the variants that actually cooccur in PubMed records and translating them into native proximity clauses, REMI produces searches that are fully transparent, more focused than Boolean AND, less dependent on the searcher anticipating every variant than manual enumeration, and able to retrieve all possible included articles in the validation sets. Native support for combining truncation and proximity in PubMed would remain the preferable long-term solution, however until it exists, REMI offers searchers a transparent and freely available alternative. In future versions of REMI, more functionalities that are not available in PubMed can be expected. For example, truncation searching for less than four characters and proximity searching with more than two sets of terms.

## Acknowledgments

The authors thank Pope Leo XIV for his spiritual support, and the Maastricht University Library Systematic Literature Review Support Team for their feedback on the entire manuscript, in particular Marieke Schor and Floor Ruiter. The authors take full responsibility for the content of the manuscript.

## Funding/Support

None

## Conflict of Interest Disclosure

Both authors share responsibility for the development of the tool described in this case report. The tool has yet to undergo independent validation.

## Data Availability Statement

REMI can be downloaded from: https://github.com/MaastrichtU-Library/remi

## Author Contributions Statement

Remigius van Alebeek: Conceptualization; methodology; software (supporting); writing – original draft; writing – review and editing. Rinus Verdonschot: Methodology; software (lead); writing – original draft; writing – review and editing.

## Notes

### Competing Interest Statement

The authors have declared no competing interest.

## References

1. National Library of Medicine (US). About PubMed [Internet]. In: PubMed. Bethesda, MD: National Library of Medicine (US) [rev. 11 Mar 2025; cited 19 Oct 2026]. https://pubmed.ncbi.nlm.nih.gov/about/.

2. National Library of Medicine (US). For Publishers [Internet]. In: National Library of Medicine. Bethesda, MD: National Library of Medicine (US) [reviewed 26 Aug 2025; cited 19 Oct 2026]. https://www.nlm.nih.gov/portals/publishers.html.

3. Bramer WM, Rethlefsen ML, Kleijnen J, Franco OH. Optimal database combinations for literature searches in systematic reviews: a prospective exploratory study. Syst Rev. 2017;6(1):245. DOI: 10.1186/s13643-017-0644-y.

4. Heintz M, Hval G, Tornes RA, Byelyey N, Hafstad E, Næss GE, Bakkeli M. Optimizing the literature search: coverage of included references in systematic reviews in Medline and Embase. J Med Libr Assoc. 2023;111(1-2):599–605. DOI: 10.5195/jmla.2023.1482.

5. Lefebvre C, Glanville J, Briscoe S, Featherstone R, Littlewood A, Metzendorf MI, Noel-Storr A, Paynter R, Rader T, Thomas J, Wieland LS. Chapter 4: Searching for and selecting studies [Internet]. In: Higgins JPT, Thomas J, Chandler J, Cumpston M, Li T, Page MJ, Welch V, eds. Cochrane Handbook for Systematic Reviews of Interventions, version 6.5.1. London: Cochrane; 2025 [updated Mar 2025; cited 19 Oct 2026]. https://www.cochrane.org/authors/handbooks-and-manuals/handbook/current/chapter-04.

6. National Library of Medicine (US). PubMed User Guide [Internet]. In: PubMed. Bethesda, MD: National Library of Medicine (US) [rev. 30 Sept 2026; cited 20 Oct 2026]. https://pubmed.ncbi.nlm.nih.gov/help/.

7. PubMed Update: Proximity Search Now Available in PubMed. NLM Tech Bull. 2022 Nov–Dec;(449):e4. [Posted 29 Nov 2022; cited 19 Oct 2026]. https://www.nlm.nih.gov/pubs/techbull/nd22/nd22_pubmed_proximity_search_available.html.

8. MacDonald H, Comer C, Foster M, Labelle PR, Marsalis S, Nyhan K, Premji Z, Rogers M, Splenda R, Stansfield C, Young S. Searching for studies: a guide to information retrieval for Campbell systematic reviews. Campbell Syst Rev. 2024;20(3):e1433. DOI: 10.1002/cl2.1433.

9. White M. Pairwise PubMed Search Generator [Internet]. GitHub; 2025 [cited 20 Oct 2026]. https://github.com/marijane/pairwise-pubmed-search-generator.

10. National Center for Biotechnology Information (US). Entrez Programming Utilities Help [Internet]. Bethesda, MD: National Center for Biotechnology Information (US); 2010–[Ongoing publication; cited 19 Oct 2026]. https://www.ncbi.nlm.nih.gov/books/NBK25501/.

11. Kim S, Cho KY, Kwon J, Sim K, Seok D, Tak H, Jo J, Eom K. An inorganic-rich SEI layer by the catalyzed reduction of LiNO3 enabled by surface-abundant hydrogen bonding for stable lithium metal batteries. Small. 2023;19(26):e2207222. DOI: 10.1002/smll.202207222.

12. Goeree KS, van der Venne WBW, Visser E, Piatkowski AA, Mouës-Vink CM. Impact of donor site on fat graft survival in autologous fat transfer to the breast: a systematic review. JPRAS Open. 2026;51:217–29. Published online 17 Jun 2026. DOI: 10.1016/j.jpra.2026.06.002.

13. Lauwens L, Ribeiro MF, Zegers CML, Høyer M, Alapetite C, Blomstrand M, Calugaru V, Di Perri D, Iannalfi A, Lütgendorf-Caucig C, Paulsen F, Postma AA, Méndèz Romero A, Timmermann B, Troost EGC, van der Weide HL, Whitfield GA, Harrabi S, Lambrecht M, Eekers DBP. Systematic review of MRI alterations in the brain following proton and photon radiation therapy: Towards a uniform European Particle Therapy Network (EPTN) definition. Radiother Oncol. 2025;208:110936. Published online 11 May 2025. DOI: 10.1016/j.radonc.2025.110936.

14. Peters LHL, Debie VHJ, Vervloet M, van Schayck OCP, Muris JWM, Gidding-Slok AHM. Promoting patient empowerment in chronic care: a scoping review of key facilitating factors. Patient Educ Couns. 2026;152:109802. Published online 29 Jul 2026. DOI: 10.1016/j.pec.2026.109802.

15. Van Ruijven B, Lantinga MA, Drenth JPH, Pierik MJ, Gevers TJG, Veldhuijzen G. Quality of evidence in remote monitoring of patients with liver cirrhosis: a systematic review. Gastro Hep Adv. 2026;5(8):100997. DOI: 10.1016/j.gastha.2026.100997.

16. Rethlefsen ML, Kirtley S, Waffenschmidt S, Ayala AP, Moher D, Page MJ, Koffel JB, PRISMA-S Group. PRISMA-S: an extension to the PRISMA Statement for Reporting Literature Searches in Systematic Reviews. Syst Rev. 2021;10(1):39. DOI: 10.1186/s13643-020-01542-z.

