## Supplementary File 1 for "REMI: A Free Tool for Truncated Proximity Searching in PubMed"

**REMI 2.1 Article | Appendices**

### Appendix A: REMI 2.1 Query Example

Table S1: Queries generated by REMI for the concepts vaccin* and hesitan* (illustrative;

variant lists abbreviated)

| **Step** | **Query** |
| --- | --- |
| Round 1, sent to PubMed | ((vaccin*[tiab])) AND ((hesitan*[tiab])) |
| Round 2, query for new forms of concept 1 | ((vaccin*[tiab]) NOT ("vaccination"[tiab] OR "vaccine"[tiab] OR …)) AND ((hesitan*[tiab])) |
| Round 2, query for new forms of concept 2 | ((vaccin*[tiab])) AND ((hesitan*[tiab]) NOT ("hesitancy"[tiab] OR "hesitant"[tiab] OR …)) |
| Round 3 onward | Same structure, with each exclusion list extended by the new variants; a concept's query stops once it yields no new variants |
| Final search to be pasted into PubMed by the user | "vaccination hesitancy"[tiab:~3] OR "vaccination hesitant"[tiab:~3] OR "vaccine hesitancy"[tiab:~3] OR … |

### Appendix B: PubMed Query Rejections

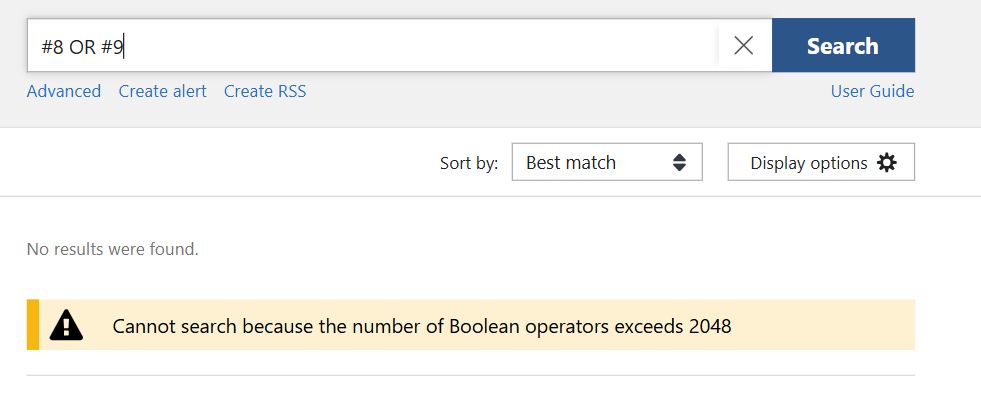

Figure S1: PubMed 2048 Boolean Limit demonstrated. This limit is also reached when attempting to combine queries through numerical references.

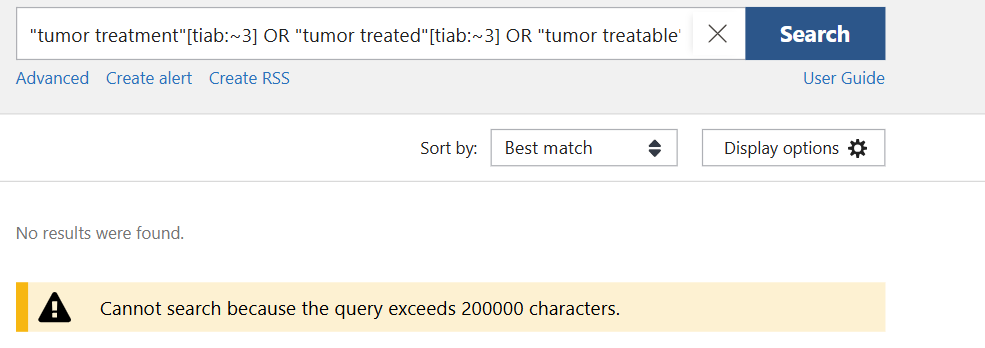

Figure S2: PubMed 200.000 Character Limit demonstrated.

### Appendix C: Keyword Search Validation Searches (15/10/2026)

#### Tumo* search:

**Truncation search:** Tumo*[tiab]
**Keyword search:** tumor[tiab] OR tumors[tiab] OR tumorous[tiab] OR tumour[tiab] OR tumourigenesis[tiab] OR tumours[tiab] OR tumori[tiab] OR tumoricidal[tiab] OR tumoroids[tiab] OR tumorsphere[tiab] OR tumourigenicity[tiab] OR tumourigenic[tiab] OR tumored[tiab] OR tumoroid[tiab] OR tumoration[tiab] OR tumorigen[tiab] OR tumorigenesis[tiab] OR tumorspheres[tiab] OR tumourectomy[tiab] OR tumorectomy[tiab] OR tumorgenesis[tiab] OR tumoral[tiab] OR tumoristatic[tiab] OR tumorbearing[tiab] OR tumorogenesis[tiab] OR tumorhead[tiab] OR tumouroids[tiab] OR tumochuan[tiab] OR tumorigens[tiab] OR tumourogenic[tiab] OR tumormarker[tiab] OR tumorigeneses[tiab] OR tumourogenesis[tiab] OR tumorigenic[tiab] OR tumorgenicity[tiab] OR tumouroid[tiab] OR tumorsuppressor[tiab] OR tumores[tiab] OR tumourous[tiab] OR tumoritropic[tiab] OR tumover[tiab] OR tumorogenic[tiab] OR tumoropathy[tiab] OR tumorigenicities[tiab] OR tumoro[tiab] OR tumorale[tiab] OR tumorlike[tiab] OR tumorboard[tiab] OR tumonolide[tiab] OR tumoren[tiab] OR tumorpromoting[tiab] OR tumous[tiab] OR tumorigenically[tiab] OR tumorigenicity[tiab] OR tumoursphere[tiab] OR tumonoic[tiab] OR tumorigenisity[tiab] OR tumoregenesis[tiab] OR tumorscan[tiab] OR tumorfusions[tiab] OR tumourogenicity[tiab] OR tumoriogenesis[tiab] OR tumorvirus[tiab] OR tumoric[tiab] OR tumourlike[tiab] OR tumorostatic[tiab] OR tumourgenic[tiab] OR tumon[tiab] OR tumoricide[tiab] OR tumorlysis[tiab] OR tumorations[tiab] OR tumorettes[tiab] OR tumorlets[tiab] OR tumoural[tiab] OR tumornecrosis[tiab] OR tumorigenecity[tiab] OR tumorigеnеsis[tiab] OR tumorresten[tiab] OR tumoración[tiab] OR tumorzentrum[tiab] OR tumorignesis[tiab] OR tumorablative[tiab] OR tumorgenecity[tiab] OR tumoregenicity[tiab] OR tumorigenetic[tiab] OR tumormarkers[tiab] OR tumorin[tiab] OR tumordokumentationssystem[tiab] OR tumorisiphum[tiab] OR tumorlocalizing[tiab] OR tumorchemosensitivity[tiab] OR tumourisation[tiab] OR tumouristatic[tiab] OR tumournecrosis[tiab] OR tumorigenesity[tiab] OR tumorbiological[tiab] OR tumolo[tiab] OR tumorus[tiab] OR tumorclones[tiab] OR tumourbiological[tiab] OR tumorresection[tiab] OR tumorinhibitory[tiab] OR tumorforming[tiab] OR tumorales[tiab] OR tumorectomie[tiab] OR tumorsubset[tiab] OR tumoic[tiab] OR tumorboards[tiab] OR tumort[tiab] OR tumorstent[tiab] OR tumore[tiab] OR tumornecrosisfactor[tiab] OR tumove[tiab] OR tumo[tiab] OR tumordicke[tiab] OR tumorization[tiab] OR tumormap[tiab] OR tumorreste[tiab] OR tumortherapie[tiab] OR tumorigensis[tiab] OR tumoralized[tiab] OR tumorigeneisis[tiab] OR tumortropic[tiab] OR tumorgrading[tiab] OR tumorigenity[tiab] OR tumova[tiab] OR tumorsyndrom[tiab] OR tumorstadium[tiab] OR tumorbedingten[tiab] OR tumourmetrics[tiab] OR tumortype[tiab] OR tumordiathesis[tiab] OR tumorlytic[tiab] OR tumorigencity[tiab] OR tumourtumour[tiab] OR tumourally[tiab] OR tumouricide[tiab] OR tumoricidial[tiab] OR tumorsstrategies[tiab] OR tumorviruses[tiab] OR tumoured[tiab] OR tumournephrectomy[tiab] OR tumout[tiab] OR tumorkranken[tiab] OR tumouritropic[tiab] OR tumorals[tiab] OR tumornecrotic[tiab] OR tumorrest[tiab] OR tumorbiology[tiab] OR tumoribus[tiab] OR tumoralis[tiab] OR tumorfree[tiab] OR tumorotropic[tiab] OR tumorcell[tiab] OR tumourspecific[tiab] OR tumourettes[tiab] OR tumorhost[tiab] OR tumorsurgery[tiab] OR tumorography[tiab] OR tumourbearing[tiab] OR tumorinhibiting[tiab] OR tumorlet[tiab] OR tumodi[tiab] OR tumorosomes[tiab] OR tumorimmune[tiab] OR tumornecrosisfactoralpha[tiab] OR tumornutrition[tiab] OR tumorally[tiab] OR tumospec[tiab] OR tumorgrowth[tiab] OR tumoros[tiab] OR tumourbed[tiab] OR tumorsagenet[tiab] OR tumorbedingte[tiab] OR tumorscope[tiab] OR tumo505[tiab] OR tumoriform[tiab] OR tumorral[tiab] OR tumoris[tiab] OR tumotezuoqi[tiab] OR tumorsversion[tiab] OR tumorgenetischen[tiab] OR tumorgraft[tiab] OR tumorsand[tiab] OR tumot[tiab] OR tumorcidal[tiab] OR tumorassociated[tiab] OR tumorspecific[tiab] OR tumorously[tiab] OR tumorcells[tiab] OR tumosynthesis[tiab] OR tumorzytogenetik[tiab] OR tumorgenetik[tiab] OR tumorentstehung[tiab] OR tumorok[tiab] OR tumorsize[tiab] OR tumordetektion[tiab] OR tumorigenicity2[tiab] OR tumorscape[tiab] OR tumorscapes[tiab] OR tumor1[tiab] OR tumor2[tiab] OR tumoour[tiab] OR tumorized[tiab] OR tumourspheroids[tiab] OR tumourspheres[tiab] OR tumorogenicity[tiab] OR tumorimmunology[tiab] OR tumorectomies[tiab] OR tumoritr[tiab] OR tumorb1[tiab] OR tumouron[tiab] OR tumouricidal[tiab] OR tumorsyndromen[tiab] OR tumorzellbiologie[tiab] OR tumourgenesis[tiab] OR tumorexzision[tiab] OR tumorsuppressive[tiab] OR tumogenesis[tiab] OR tumorzellreste[tiab] OR tumourlets[tiab] OR tumorgewebe[tiab] OR tumorentitäten[tiab] OR tumorgenic[tiab] OR tumorangiogenesis[tiab] OR tumorassoziierte[tiab] OR tumorerkrankungen[tiab] OR tumortömegét[tiab] OR tumorosphere[tiab] OR tumorzellen[tiab] OR tumourcharacteristics[tiab] OR tumorigenicitiy[tiab] OR tumoursand[tiab] OR tumorkines[tiab] OR tumorkine[tiab] OR tumorial[tiab] OR tumororthopaedics[tiab] OR tumorrelated[tiab] OR tumorstages[tiab] OR tumorolytic[tiab] OR tumorcode[tiab] OR tumorprosthesis[tiab] OR tumorfibula[tiab] OR tumornode[tiab] OR tumorifrons[tiab] OR tumorexzisionen[tiab] OR tumors1[tiab] OR tumorapa[tiab] OR tumorprotein[tiab] OR tumorsthat[tiab] OR tumorinvasion[tiab] OR tumoroperationen[tiab] OR tumorvorsorge[tiab] OR tumorähnliche[tiab] OR tumopr[tiab] OR tumornekrosefaktor[tiab] OR tumorgeschwulst[tiab] OR tumorresektion[tiab] OR tumorbasistherapie[tiab] OR tumourassociated[tiab] OR tumorcelllines[tiab] OR tumorentfernung[tiab] OR tumorresponsewas[tiab] OR tumorogenisity[tiab] OR tumorzellgrößen[tiab] OR tumoururgent[tiab] OR tumourhypertrophic[tiab] OR tumornodes[tiab] OR tumorigentic[tiab] OR tumorswere[tiab] OR tumorwachstum[tiab] OR tumortargeting[tiab] OR tumoricides[tiab] OR tumorogensis[tiab] OR tumorimmunologische[tiab] OR tumoteyou[tiab] OR tumorispheres[tiab] OR tumorigenenesis[tiab] OR tumoir[tiab] OR tumoregensis[tiab] OR tumouroccurrences[tiab] OR tumoring[tiab] OR tumorigeneis[tiab] OR tumography[tiab] OR tumorinduced[tiab] OR tumorellenes[tiab] OR tumorlocation[tiab] OR tumourand[tiab] OR tumorepidemiológia[tiab] OR tumorigeicity[tiab] OR tumoralpotent[tiab] OR tumordforegoing[tiab] OR tumorspheric[tiab] OR tumorinhibitor[tiab] OR tumorigeneity[tiab] OR tumorvirgae[tiab] OR tumorigenensis[tiab] OR tumornecross[tiab] OR tumorogenecity[tiab] OR tumoursto[tiab] OR tumourof[tiab] OR tumorderived[tiab] OR tumorexstirpation[tiab] OR tumorigenes[tiab] OR tumorospheres[tiab] OR tumouric[tiab] OR tumoropathic[tiab] OR tumorigenzcity[tiab] OR tumorsthatare[tiab] OR tumorigenisis[tiab] OR tumorilor[tiab] OR tumorangiognesis[tiab] OR tumorigeness[tiab] OR tumorbiologie[tiab] OR tumour1[tiab] OR tumouts[tiab] OR tumorablations[tiab] OR tumorablation[tiab] OR tumoricidals[tiab] OR tumorexpressing[tiab] OR tumouration[tiab] OR tumorogenesity[tiab] OR tumordisease[tiab] OR tumorcerebri[tiab] OR tumorless[tiab] OR tumortumour[tiab] OR tumoxetine[tiab] OR tumouran[tiab] OR tumory[tiab] OR tumorstromal[tiab] OR tumorsuppressors[tiab] OR tumourology[tiab] OR tumorsyndrome[tiab] OR tumorbed[tiab] OR tumoreginicity[tiab] OR tumortropism[tiab] OR tumotectomy[tiab] OR tumorto[tiab] OR tumorsranged[tiab] OR tumorical[tiab] OR tumonrs[tiab] OR tumorigram[tiab] OR tumorregression[tiab] OR tumoractivated[tiab] OR tumoursupressor[tiab] OR tumoronecroticance[tiab] OR tumorembolisation[tiab] OR tumourgrowth[tiab] OR tumouristasis[tiab] OR tumorradicality[tiab] OR tumorgraphy[tiab] OR tumonisin[tiab] OR tumorigeneic[tiab] OR tumoricid[tiab] OR tumourpositive[tiab] OR tumorectomia[tiab] OR tumorcytotoxic[tiab] OR tumoursize[tiab] OR tumoursis[tiab] OR tumourprotective[tiab] OR tumorzentren[tiab] OR tumosa[tiab] OR tumourfluoroscopy[tiab] OR tumourectomies[tiab] OR tumoritrophism[tiab] OR tumorette[tiab] OR tumorpatients[tiab] OR tumorsera[tiab] OR tumoricity[tiab] OR tumorosteolysis[tiab] OR tumortherapy[tiab] OR tumorlyzing[tiab] OR tumourchemotherapy[tiab] OR tumortoxic[tiab] OR tumourimmunology[tiab] OR tumorinfiltrated[tiab] OR tumoraspect[tiab] OR tumoruous[tiab] OR tumourmamongst[tiab] OR tumours6[tiab] OR tumourcells[tiab] OR tumorsuspected[tiab] OR tumordosis[tiab] OR tumorogeneity[tiab] OR tumorlia[tiab] OR tumoriformis[tiab] OR tumortracer[tiab] OR tumorscanning[tiab] OR tumorophilic[tiab] OR tumorgenom[tiab] OR tumormetric[tiab] OR tumorinduction[tiab] OR tumourfree[tiab] OR tumorantigens[tiab] OR tumorae[tiab] OR tumorons[tiab] OR tumorts[tiab] OR tumofascient[tiab] OR tumorr[tiab] OR tumofacient[tiab] OR tumourfilaroides[tiab] OR tumora[tiab] OR tumoraffin[tiab] OR tumourinduction[tiab] OR tumourless[tiab] OR tumoar[tiab]

#### Polic* search:

**Truncated search:** Polic*[tiab] **Keyword search:** policy[tiab] OR policosanol[tiab] OR policemen[tiab] OR policies[tiab] OR policosanols[tiab] OR policymakers[tiab] OR policeman[tiab] OR policlinics[tiab] OR police[tiab] OR policresulen[tiab] OR policy1[tiab] OR policystic[tiab] OR policyholder[tiab] OR policis[tiab] OR policaptil[tiab] OR policewomen[tiab] OR policryps[tiab] OR polichemotherapy[tiab] OR policlonal[tiab] OR policard[tiab] OR policitemia[tiab] OR policymap[tiab] OR policymaking[tiab] OR policyclic[tiab] OR policosanolplus[tiab] OR policlinique[tiab] OR policoro[tiab] OR policing[tiab] OR policimbac[tiab] OR policlinica[tiab] OR policello[tiab] OR policaris[tiab] OR policlänico[tiab] OR polic[tiab] OR polici[tiab] OR policaju[tiab] OR policardiographic[tiab] OR policoff[tiab] OR policlínica[tiab] OR policare[tiab] OR polich[tiab] OR polichlorinated[tiab] OR policondritis[tiab] OR policistronic[tiab] OR polichnitos[tiab] OR policks[tiab] OR polichromatic[tiab] OR polichni[tiab] OR policardiography[tiab] OR policatan[tiab] OR policewoman[tiab] OR policaprolactone[tiab] OR policomprimido[tiab] OR polications[tiab] OR policarbone[tiab] OR policresulene[tiab] OR polichia[tiab] OR policias[tiab] OR policlinico[tiab] OR polices[tiab] OR polichno[tiab] OR policuspidifera[tiab] OR policia[tiab] OR policyrelated[tiab] OR policeraty[tiab] OR policyfile[tiab] OR policarpa[tiab] OR polichnodes[tiab] OR policetidis[tiab] OR policondrite[tiab] OR policlonality[tiab] OR policístico[tiab] OR policythemia[tiab] OR policytemia[tiab] OR policarpov[tiab] OR polichloride[tiab] OR policistic[tiab] OR policarbophil[tiab] OR polichem[tiab] OR policyventure[tiab] OR policentrico[tiab] OR polication[tiab] OR policka[tiab] OR policattedra[tiab] OR policarbonate[tiab] OR policarboxylate[tiab] OR polichromic[tiab] OR policardiograms[tiab] OR polichotomous[tiab] OR policymaker[tiab] OR policlinic[tiab] OR policyholders[tiab] OR policypreparação[tiab] OR policed[tiab] OR policythis[tiab] OR policomorbidities[tiab] OR policarpo[tiab] OR policymonthly[tiab] OR policer[tiab] OR policythe[tiab] OR policdk[tiab] OR policronamos[tiab] OR policychain[tiab] OR policytree[tiab] OR policywise[tiab] OR policrisis[tiab] OR polical[tiab] OR policings[tiab] OR polichondritis[tiab] OR polichete[tiab] OR policlonales[tiab] OR policymodels[tiab] OR policistico[tiab] OR polichaete[tiab] OR policital[tiab] OR polick[tiab] OR policyrationing[tiab] OR policoria[tiab] OR policía[tiab] OR policarboxylic[tiab] OR policonsumo[tiab] OR policéntrica[tiab] OR policlincoumberto[tiab] OR policromática[tiab] OR policheta[tiab] OR policaprolacotne[tiab] OR policórica[tiab] OR policystronic[tiab] OR policordia[tiab] OR policlonalidade[tiab] OR policlonais[tiab] OR policymarkers[tiab] OR policalicosis[tiab] OR policlinically[tiab] OR policot[tiab] OR policarbonyl[tiab] OR policymakings[tiab] OR polichemotherapeutic[tiab] OR policisztásovárium[tiab] OR policarboksilate[tiab] OR polichaeta[tiab] OR policyagendas[tiab] OR policristalyne[tiab] OR policlinical[tiab] OR policíclicos[tiab] OR policers[tiab] OR policization[tiab] OR polichimiotherapy[tiab] OR policyarenas[tiab] OR policarboxyboltorn[tiab] OR policarpio[tiab] OR policyland[tiab] OR policaproamide[tiab] OR policyfacilitated[tiab] OR policocanol[tiab] OR policastillo[tiab] OR policyst[tiab] OR policromathohilic[tiab] OR policontusions[tiab] OR policaries[tiab] OR policey[tiab] OR policastro[tiab] OR polichaetes[tiab] OR policationic[tiab] OR policie[tiab] OR policentric[tiab] OR policínica[tiab] OR polichromatophilous[tiab] OR policyandguidance[tiab] OR policyenabling[tiab] OR polictilenglycole[tiab] OR policl[tiab] OR polichaets[tiab] OR policlithrum[tiab] OR polichiomiotherapy[tiab] OR policitor[tiab] OR policlm[tiab] OR policlonally[tiab] OR policial[tiab] OR policehaving[tiab] OR policiessince[tiab] OR polichromasia[tiab] OR polichak[tiab] OR polichlorodibenzofurans[tiab] OR polichlorodibenzo[tiab] OR policystosis[tiab] OR policiessydney[tiab] OR policyplanners[tiab] OR policínico[tiab] OR policydublin[tiab] OR policity[tiab] OR policurved[tiab] OR policarpine[tiab] OR policansky[tiab] OR polichinelle[tiab] OR polichrome[tiab] OR policus[tiab] OR polichemiotherapy[tiab] OR policephalus[tiab] OR polichemioterapy[tiab] OR polichemio[tiab] OR polichemotherapic[tiab] OR policithemic[tiab] OR policydal[tiab] OR polica[tiab] OR policlincs[tiab] OR policlänica[tiab] OR policerate[tiab]

#### Health* search:

**Truncation search**: Health*[tiab]
**Keyword search:** health[tiab] OR healthspan[tiab] OR healthcare[tiab] OR healthy[tiab] OR healthful[tiab] OR healthiness[tiab] OR healthsouth[tiab] OR healthcore[tiab] OR healthfulness[tiab] OR healthtrust[tiab] OR healthiest[tiab] OR healthstyles[tiab] OR healthgrades[tiab] OR healthly[tiab] OR healthsystems[tiab] OR healthgrid[tiab] OR healthdyne[tiab] OR healthily[tiab] OR healthverity[tiab] OR healtheast[tiab] OR healthone[tiab] OR healthlines[tiab] OR healthline[tiab] OR health2006[tiab] OR healthplan[tiab] OR healthgrids[tiab] OR healthtech[tiab] OR healthkit[tiab] OR healthsource[tiab] OR healthscape[tiab] OR healthways[tiab] OR healthpoint[tiab] OR healthstats[tiab] OR healthtronics[tiab] OR healthconnect[tiab] OR healthware[tiab] OR healthscan[tiab] OR healthism[tiab] OR healthpatch[tiab] OR healthdot[tiab] OR healthrota[tiab] OR healthright[tiab] OR healthpass[tiab] OR healthstar[tiab] OR healthid[tiab] OR healthmpowerment[tiab] OR healthimpact[tiab] OR healthvest[tiab] OR healthnuts[tiab] OR healthwatch[tiab] OR healthnet[tiab] OR healthcheck[tiab] OR health2000[tiab] OR healthtracker[tiab] OR healthtrack[tiab] OR healthworks[tiab] OR healthgate[tiab] OR healthsystem[tiab] OR healthium[tiab] OR healthsports[tiab] OR healthyhearts[tiab] OR healthboard[tiab] OR healthtexas[tiab] OR healthcom[tiab] OR healthstart[tiab] OR healthplix[tiab] OR healthbench[tiab] OR healthtree[tiab] OR healthinformaticslab[tiab] OR healthecco[tiab] OR healthorganization[tiab] OR health2008[tiab] OR healthcoin[tiab] OR health2[tiab] OR healthydiet[tiab] OR healthquest[tiab] OR healthquiz[tiab] OR healthcorp[tiab] OR healthdata[tiab] OR healthprompt[tiab] OR healthberry[tiab] OR healthycow24[tiab] OR health1[tiab] OR healthinfonet[tiab] OR healthlnk[tiab] OR healthspace[tiab] OR healthplus[tiab] OR healthchoice[tiab] OR healtheon[tiab] OR healthco[tiab] OR healthlawyer[tiab] OR healthwest[tiab] OR healthier[tiab] OR healthc[tiab] OR healthspans[tiab] OR healthmail[tiab] OR healthrelated[tiab] OR healthforce[tiab] OR healthycontrols[tiab] OR healthwise[tiab] OR healthmindr[tiab] OR healthsense[tiab] OR healthylact[tiab] OR healthinsurance[tiab] OR healthyair[tiab] OR healthcareer[tiab] OR healthmeasures[tiab] OR healthymoms[tiab] OR healtheathlete[tiab] OR healthstyle[tiab] OR healthstreet[tiab] OR healthcarers[tiab] OR healthfood[tiab] OR healthprofessional[tiab] OR healthtalkonline[tiab] OR healthscope[tiab] OR healthplans[tiab] OR healthport[tiab] OR healthspeak[tiab] OR healthairegister[tiab] OR healthpathways[tiab] OR healthlink[tiab] OR healthymind[tiab] OR healthpartners[tiab] OR healthgpt[tiab] OR health31[tiab] OR healthcareassociated[tiab] OR health4life[tiab] OR healthjump[tiab] OR healths[tiab] OR healthyplant[tiab] OR healthcall[tiab] OR healthydonors[tiab] OR healthenhancing[tiab] OR healthcareprofessionals[tiab] OR healthentia[tiab] OR healthtwist[tiab] OR healthproblem[tiab] OR healtherx[tiab] OR healthypeople[tiab] OR healthand[tiab] OR healthgivers[tiab] OR healthspring[tiab] OR healthcentre[tiab] OR health49[tiab] OR healthmpowers[tiab] OR healthpact[tiab] OR healthrisk[tiab] OR healthsciences[tiab] OR healthkeeping[tiab] OR healthieru[tiab] OR healthpayer[tiab] OR healthcares[tiab] OR healthviews[tiab] OR healthyhay[tiab] OR healthtm[tiab] OR healthnetwork[tiab] OR healthphysics[tiab] OR healthbridge[tiab] OR healthicisation[tiab] OR healthcarer[tiab] OR healthwear[tiab] OR healthe[tiab] OR healthicization[tiab] OR healthies[tiab] OR healthfoods[tiab] OR healthnews[tiab] OR healthshare[tiab] OR healthcontrol[tiab] OR healthcareopathy[tiab] OR healthless[tiab] OR healthward[tiab] OR healthseekers[tiab] OR healthineers[tiab] OR healthyroad[tiab] OR healthinequities[tiab] OR healthstat[tiab] OR health4all[tiab] OR healthcase[tiab] OR healthcarepolicies[tiab] OR healthaid[tiab] OR health2sync[tiab] OR healthytravel[tiab] OR healthcommunication[tiab] OR healthcarequality[tiab] OR healthypregnancy[tiab] OR healthprofessionsed[tiab] OR health5[tiab] OR healthconscious[tiab] OR healthkart[tiab] OR healthliners[tiab] OR healthdisparities[tiab] OR healthmark[tiab] OR healthmap[tiab] OR healthya[tiab] OR healthchallenge[tiab] OR healthocide[tiab] OR healthyfirst[tiab] OR healthcomm[tiab] OR healthyaging[tiab] OR healthx[tiab] OR healthgrade[tiab] OR healthworkers[tiab] OR healthcanadact[tiab] OR healthylaife[tiab] OR healthreact[tiab] OR healthdirect[tiab] OR healththis[tiab] OR healthreplyresting[tiab] OR healthcarecosts[tiab] OR healthkam[tiab] OR healthieir[tiab] OR healthp50[tiab] OR health1ul1tr002373[tiab] OR healthbeneficial[tiab] OR healthcareaccess[tiab] OR healthequality[tiab] OR healthcareworkers[tiab] OR healthich[tiab] OR healthmyne[tiab] OR healthml[tiab] OR healthyspan[tiab] OR healthvcf[tiab] OR healthpolicy[tiab] OR healthcenters[tiab] OR healthpal[tiab] OR healthineer[tiab] OR healthpromoting[tiab] OR healthymale[tiab] OR healthwashing[tiab] OR healthtalk[tiab] OR healthimproving[tiab] OR healthfully[tiab] OR healthbot[tiab] OR healthstroke[tiab] OR healthfacts[tiab] OR healthgrain[tiab] OR healthrisks[tiab] OR healthrecord[tiab] OR healthequity[tiab] OR healthlab[tiab] OR healthchain[tiab] OR healthforanimals[tiab] OR healthsearchqa[tiab] OR healthsight[tiab] OR healthorganizational[tiab] OR healthcarestablishment[tiab] OR healthissue[tiab] OR healthforall[tiab] OR health4cll[tiab] OR healthpsychologyresearch[tiab] OR healthymargin[tiab] OR healthylactating[tiab] OR healthystart[tiab] OR health14[tiab] OR healthy2go[tiab] OR healthproblemwith[tiab] OR healthcarehas[tiab] OR healthable[tiab] OR healthunlocked[tiab] OR healthhas[tiab] OR healthinternet[tiab] OR health3[tiab] OR healthcarea[tiab] OR healthist[tiab] OR health4sport[tiab] OR healthscience[tiab] OR healthy16[tiab] OR healthpulse[tiab] OR healthlth[tiab] OR healthboards[tiab] OR healthleaders[tiab] OR healthorganisation[tiab] OR healthysun[tiab] OR healthliterature[tiab] OR healthbeliefs[tiab] OR healththreatening[tiab] OR healthstatistics[tiab] OR healthendorsing[tiab] OR healthbenefits[tiab] OR healthyrabbits[tiab] OR healthquality[tiab] OR healthysika[tiab] OR healthinflammatory[tiab] OR healthsaving[tiab] OR healthebikes[tiab] OR healthebike[tiab] OR healthprofessionals[tiab] OR healthyøadipose[tiab] OR healthywomen[tiab] OR healthyfood[tiab] OR healthconcern[tiab] OR healthservice[tiab] OR healthsoaf[tiab] OR healthmapper[tiab] OR healthcar[tiab] OR healthmanagement[tiab] OR healthspot[tiab] OR healthywater[tiab] OR health4men[tiab] OR healthyworkinglives[tiab] OR healthybrains[tiab] OR healthresources[tiab] OR healthy40[tiab] OR healthprovider[tiab] OR healthopolis[tiab] OR healthties[tiab] OR healthseq[tiab] OR healthrelevant[tiab] OR healthmanifesto[tiab] OR healththe[tiab] OR healthptprothrombin[tiab] OR healthsys[tiab] OR healthography[tiab] OR healthreach[tiab] OR healthcentric[tiab] OR healthynurse[tiab] OR healthhy[tiab] OR healthroster[tiab] OR healthtrans[tiab] OR healtheway[tiab] OR healthwas[tiab] OR healthcybermap[tiab] OR healthgroup[tiab] OR healthyct[tiab] OR healthdcare[tiab] OR healthstream[tiab] OR healthy18[tiab] OR healthinstitute[tiab] OR healthiron[tiab] OR healthwell[tiab] OR healthipedia[tiab] OR healthquery[tiab] OR healthresort[tiab] OR healthedited[tiab] OR healthdesign[tiab] OR healthlight[tiab] OR healthvault[tiab] OR healthyclients[tiab] OR healthmetrics[tiab] OR healthforceontario[tiab] OR healthmatics[tiab] OR healthchem[tiab] OR healthcareby[tiab] OR healthsurvey[tiab] OR healthfinder[tiab] OR healtheconomic[tiab] OR healthia[tiab] OR healthscreen[tiab] OR healthicizing[tiab] OR healthytissue[tiab] OR healthandage[tiab] OR healthicised[tiab] OR healthseeking[tiab] OR healthcarepapers[tiab] OR healthpolitical[tiab] OR healthmedia[tiab] OR healthfield[tiab] OR healthas[tiab] OR healthresearchfunding[tiab] OR healthcentres[tiab] OR healthtest[tiab] OR healthitia[tiab] OR healthworker[tiab] OR healthierus[tiab] OR healtheries[tiab] OR healthsentry[tiab] OR healthetech[tiab] OR healthyyoung[tiab] OR healthact[tiab] OR healthnexis[tiab] OR healthyhuman[tiab] OR healthcarefuture[tiab] OR healthallies[tiab] OR healthvision[tiab] OR healthcentral[tiab] OR healthwave[tiab] OR healthsci[tiab] OR healthcarereportcards[tiab] OR healthcareinterventions[tiab] OR healthweb[tiab] OR healthchoices[tiab] OR healthlinks[tiab] OR healtharium[tiab] OR healthscapes[tiab] OR healthmaster[tiab] OR healthedco[tiab] OR healthbook[tiab] OR healthfocus[tiab] OR healthfarm[tiab] OR healthpark[tiab] OR healthcenter[tiab] OR healthchek[tiab] OR healthletter[tiab] OR healthwin[tiab] OR healthamerica[tiab] OR healthsportcenter[tiab] OR healthcraft[tiab] OR healthyland[tiab] OR healthstate[tiab] OR healthland[tiab] OR healthcaremanship[tiab] OR healtha[tiab] OR healthmaintenance[tiab]

#### Semaglut* search:

**Truncation search:** Semaglut*[tiab]
**Keyword search:** semaglutide[tiab] OR semaglutides[tiab] OR semaglutie[tiab]

### Appendix D: Proximity Search Validation Searches (19/10/2026)

#### Search 1 (Lauwens et al.): https://pubmed.ncbi.nlm.nih.gov/40360047/

##### Search 1: Original MEDLINE Search: (9937 hits)

(exp "Radiotherapy"/ **OR** "Radiotherapy".fs. **OR** ("Radiotherapy" **OR** "chemoradiotherapy" **OR** "radiochemotherapy" **OR** (("Proton" **OR** "Photon" **OR** "Radiation" **OR** "particle") adj3 ("therap*"))).ti,ab,kf.) **AND**

(exp "Meningeal Neoplasms"/ **OR** exp "Nasopharyngeal Neoplasms"/ **OR** exp "Skull Base Neoplasms"/ **OR** exp "Glioma"/ **OR** exp "Germinoma"/ **OR** exp "Pituitary Neoplasms"/ **OR** exp "Brain Neoplasms"/ **OR** exp "Paranasal Sinus Neoplasms"/ **OR** exp "Craniopharyngioma"/ **OR** exp "Pinealoma"/ **OR** exp"Chordoma"/ **OR** exp "Chondrosarcoma"/ **OR** exp "Meningioma"/ **OR** ("meningioma*" **OR** "Glioma*" **OR** "Glial Cell*" **OR** "Astrocytoma*" **OR** "Astroglioma*" **OR** "Oligoastrocytoma*" **OR** "Xanthoastrocytoma*" **OR** "DIPG" **OR** "Ependymoma*" **OR** "Medulloblastoma*" **OR** "Oligodendroglioma*" **OR** "Oligodendroblastoma*" **OR** "craniopharyngioma*" **OR** "Germinoma*" **OR** "chondrosarcoma*" **OR** "chordoma*" **OR** "meningeoma*" **OR** "oligodendrocytoma*" **OR** "pinealoma*" **OR** (("Atypical Teratoid Rhabdoid" **OR** "pituitary" **OR** "Brain" **OR** "Skull Base" **OR** "Nasopharynx" **OR** "Nasopharyngeal" **OR** "meningeal" **OR** "sinus" **OR** "intracranial" **OR** "cerebral" **OR** "subtentorial" **OR** "infratentorial" **OR** "supratentorial" **OR** "pituitary gland" **OR** "hypophyseal" **OR** "hypophysis" **OR** "cranial base" **OR** "craniobasal" **OR** "germinal" **OR** "craniopharyngeal" **OR** "glial" **OR** "ependymal") adj3 ("cancer*" **OR** "neoplasm*" **OR** "tumor*" **OR** "tumour*" **OR** "carcinoma*" **OR** "sarcoma*"))).ti,ab,kf.) **AND**

(exp "Necrosis"/ **OR** exp "Radiation Injuries"/ **OR** exp "Abnormalities, Radiation-Induced"/ **OR** "Diagnostic imaging".fs. **OR** ("necros*" **OR** "radionecros*" **OR** "radio-necros*" **OR** "pseudoprogress*" **OR** "image change*" **OR** "imaging change*" **OR** "radiographic change*" **OR** "radiological change*" **OR** "radiographic change*" **OR** "Radiation Injur*" **OR** "Radiation Induced Abnormalit*" **OR** "radiation induced contrast enhancement*" **OR** "brain stem injur*" **OR** "brainstem injur*" **OR** "RIBI" **OR** "CEBL" **OR** "RICE" **OR** (("contrast-enhanc*" **OR** "radiation induc*") adj3 ("lesion*" **OR** "leasion*"))).ti,ab,kf.)

**NOT** ((exp "animal"/ NOT exp "human"/) **OR** ("mouse" **OR** "mice" **OR** "rat" **OR** "rats" **OR** "rodent*" **OR** "pig" **OR** "pigs" **OR** "cattle" **OR** "cow" **OR** "cows" **OR** "horse*" **OR** "monkey*" **OR** "dog" **OR** "dogs" **OR** "cat" **OR** "cats" **OR** "fish" **OR** "mammal*" **OR** "murine*" **OR** "ewe" **OR** "ewes" **OR** "sheep" **OR** "zebrafish").ti) **NOT**(("review" **OR** "preprints" **OR** "conference abstract" **OR** "letter" **OR** "editorials" **OR** "note" **OR** "short survey").pt **OR** ("letter" **OR** "reply" **OR** "response" **OR** "editor*" **OR** "preface" **OR** "opinion" **OR** "meta-analysis" **OR** "meta-synthesis" **OR** "review").ti) **NOT**(exp *brain metastasis/ **OR** "metastas*".ti)

##### Search 1: REMI 2 Proximity Search convert to Medline (9945 hits):

(exp "Radiotherapy"/ **OR** "Radiotherapy".fs. **OR** ("Radiotherapy" **OR** "chemoradiotherapy" **OR** "radiochemotherapy" **OR** (((proton ADJ3 therapy) OR (proton ADJ3 therapeutic) OR (proton ADJ3 therapies) OR (proton ADJ3 therapeutical) OR (proton ADJ3 therapeutics) OR (proton ADJ3 therapsids) OR (proton ADJ3 therapsid) OR (proton ADJ3 therapeutically) OR (proton ADJ3 therapists) OR (proton ADJ3 therapsida) OR (proton ADJ3 theraplan) OR (proton ADJ3 theraphy) OR (proton ADJ3 therapanacea) OR (proton ADJ3 therapis) OR (proton ADJ3 therapep) OR (proton ADJ3 theraphosidae) OR (proton ADJ3 therapeuticals) OR (proton ADJ3 therapax) OR (proton ADJ3 theraphosa) OR (proton ADJ3 theraputic) OR (proton ADJ3 therapist) OR (proton ADJ3 therapyis) OR (proton ADJ3 therapancea) OR (proton ADJ3 therapty) OR (proton ADJ3 therapeutant) OR (proton ADJ3 therapyand) OR (proton ADJ3 theraphosoidina) OR (proton ADJ3 therapyimaging) OR (proton ADJ3 therapeuticpreventive) OR (proton ADJ3 therapying) OR (proton ADJ3 therapueutic) OR (proton ADJ3 therapeut) OR (proton ADJ3 therapeutists) OR (proton ADJ3 theraphosid) OR (proton ADJ3 therapentical) OR (proton ADJ3 therapie) OR (proton ADJ3 theraplane) OR (proton ADJ3 therapeutische) OR (photon ADJ3 therapy) OR (photon ADJ3 therapeutic) OR (photon ADJ3 therapies) OR (photon ADJ3 therapeutical) OR (photon ADJ3 therapeutics) OR (photon ADJ3 therapsids) OR (photon ADJ3 therapsid) OR (photon ADJ3 therapeutically) OR (photon ADJ3 therapists) OR (photon ADJ3 therapsida) OR (photon ADJ3 theraplan) OR (photon ADJ3 theraphy) OR (photon ADJ3 therapanacea) OR (photon ADJ3 therapis) OR (photon ADJ3 therapep) OR (photon ADJ3 theraphosidae) OR (photon ADJ3 therapeuticals) OR (photon ADJ3 therapax) OR (photon ADJ3 theraphosa) OR (photon ADJ3 theraputic) OR (photon ADJ3 therapist) OR (photon ADJ3 therapyis) OR (photon ADJ3 therapancea) OR (photon ADJ3 therapty) OR (photon ADJ3 therapeutant) OR (photon ADJ3 therapyand) OR (photon ADJ3 theraphosoidina) OR (photon ADJ3 therapyimaging) OR (photon ADJ3 therapeuticpreventive) OR (photon ADJ3 therapying) OR (photon ADJ3 therapueutic) OR (photon ADJ3 therapeut) OR (photon ADJ3 therapeutists) OR (photon ADJ3 theraphosid) OR (photon ADJ3 therapentical) OR (photon ADJ3 therapie) OR (photon ADJ3 theraplane) OR (photon ADJ3 therapeutische) OR (radiation ADJ3 therapy) OR (radiation ADJ3 therapeutic) OR (radiation ADJ3 therapies) OR (radiation ADJ3 therapeutical) OR (radiation ADJ3 therapeutics) OR (radiation ADJ3 therapsids) OR (radiation ADJ3 therapsid) OR (radiation ADJ3 therapeutically) OR (radiation ADJ3 therapists) OR (radiation ADJ3 therapsida) OR (radiation ADJ3 theraplan) OR (radiation ADJ3 theraphy) OR (radiation ADJ3 therapanacea) OR (radiation ADJ3 therapis) OR (radiation ADJ3 therapep) OR (radiation ADJ3 theraphosidae) OR (radiation ADJ3 therapeuticals) OR (radiation ADJ3 therapax) OR (radiation ADJ3 theraphosa) OR (radiation ADJ3 theraputic) OR (radiation ADJ3 therapist) OR (radiation ADJ3 therapyis) OR (radiation ADJ3 therapancea) OR (radiation ADJ3 therapty) OR (radiation ADJ3 therapeutant) OR (radiation ADJ3 therapyand) OR (radiation ADJ3 theraphosoidina) OR (radiation ADJ3 therapyimaging) OR (radiation ADJ3 therapeuticpreventive) OR (radiation ADJ3 therapying) OR (radiation ADJ3 therapueutic) OR (radiation ADJ3 therapeut) OR (radiation ADJ3 therapeutists) OR (radiation ADJ3 theraphosid) OR (radiation ADJ3 therapentical) OR (radiation ADJ3 therapie) OR (radiation ADJ3 theraplane) OR (radiation ADJ3 therapeutische) OR (particle ADJ3 therapy) OR (particle ADJ3 therapeutic) OR (particle ADJ3 therapies) OR (particle ADJ3 therapeutical) OR (particle ADJ3 therapeutics) OR (particle ADJ3 therapsids) OR (particle ADJ3 therapsid) OR (particle ADJ3 therapeutically) OR (particle ADJ3 therapists) OR (particle ADJ3 therapsida) OR (particle ADJ3 theraplan) OR (particle ADJ3 theraphy) OR (particle ADJ3 therapanacea) OR (particle ADJ3 therapis) OR (particle ADJ3 therapep) OR (particle ADJ3 theraphosidae) OR (particle ADJ3 therapeuticals) OR (particle ADJ3 therapax) OR (particle ADJ3 theraphosa) OR (particle ADJ3 theraputic) OR (particle ADJ3 therapist) OR (particle ADJ3 therapyis) OR (particle ADJ3 therapancea) OR (particle ADJ3 therapty) OR (particle ADJ3 therapeutant) OR (particle ADJ3 therapyand) OR (particle ADJ3 theraphosoidina) OR (particle ADJ3 therapyimaging) OR (particle ADJ3 therapeuticpreventive) OR (particle ADJ3 therapying) OR (particle ADJ3 therapueutic) OR (particle ADJ3 therapeut) OR (particle ADJ3 therapeutists) OR (particle ADJ3 theraphosid) OR (particle ADJ3 therapentical) OR (particle ADJ3 therapie) OR (particle ADJ3 theraplane) OR (particle ADJ3 therapeutische)))).ti,ab,kf.) **AND**

(exp "Meningeal Neoplasms"/ **OR** exp "Nasopharyngeal Neoplasms"/ **OR** exp "Skull Base Neoplasms"/ **OR** exp "Glioma"/ **OR** exp "Germinoma"/ **OR** exp "Pituitary Neoplasms"/ **OR** exp "Brain Neoplasms"/ **OR** exp "Paranasal Sinus Neoplasms"/ **OR** exp "Craniopharyngioma"/ **OR** exp "Pinealoma"/ **OR** exp"Chordoma"/ **OR** exp "Chondrosarcoma"/ **OR** exp "Meningioma"/ **OR** ("meningioma*" **OR** "Glioma*" **OR** "Glial Cell*" **OR** "Astrocytoma*" **OR** "Astroglioma*" **OR** "Oligoastrocytoma*" **OR** "Xanthoastrocytoma*" **OR** "DIPG" **OR** "Ependymoma*" **OR** "Medulloblastoma*" **OR** "Oligodendroglioma*" **OR** "Oligodendroblastoma*" **OR** "craniopharyngioma*" **OR** "Germinoma*" **OR** "chondrosarcoma*" **OR** "chordoma*" **OR** "meningeoma*" **OR** "oligodendrocytoma*" **OR** "pinealoma*" **OR** (((atypical ADJ3 teratoid ADJ3 rhabdoid ADJ3 cancer) OR (atypical ADJ3 teratoid ADJ3 rhabdoid ADJ3 cancerous) OR (atypical ADJ3 teratoid ADJ3 rhabdoid ADJ3 cancers) OR (atypical ADJ3 teratoid ADJ3 rhabdoid ADJ3 cancerogenesis) OR (atypical ADJ3 teratoid ADJ3 rhabdoid ADJ3 canceration) OR (atypical ADJ3 teratoid ADJ3 rhabdoid ADJ3 cancerogenic) OR (atypical ADJ3 teratoid ADJ3 rhabdoid ADJ3 cancerocerebral) OR (atypical ADJ3 teratoid ADJ3 rhabdoid ADJ3 neoplasms) OR (atypical ADJ3 teratoid ADJ3 rhabdoid ADJ3 neoplasm) OR (atypical ADJ3 teratoid ADJ3 rhabdoid ADJ3 neoplasma) OR (atypical ADJ3 teratoid ADJ3 rhabdoid ADJ3 neoplasmic) OR (atypical ADJ3 teratoid ADJ3 rhabdoid ADJ3 neoplasmatic) OR (atypical ADJ3 teratoid ADJ3 rhabdoid ADJ3 neoplasmas) OR (atypical ADJ3 teratoid ADJ3 rhabdoid ADJ3 tumor) OR (atypical ADJ3 teratoid ADJ3 rhabdoid ADJ3 tumors) OR (atypical ADJ3 teratoid ADJ3 rhabdoid ADJ3 tumorlike) OR (atypical ADJ3 teratoid ADJ3 rhabdoid ADJ3 tumoricidal) OR (atypical ADJ3 teratoid ADJ3 rhabdoid ADJ3 tumorigenesis) OR (atypical ADJ3 teratoid ADJ3 rhabdoid ADJ3 tumorectomy) OR (atypical ADJ3 teratoid ADJ3 rhabdoid ADJ3 tumorigenic) OR (atypical ADJ3 teratoid ADJ3 rhabdoid ADJ3 tumorgenicity) OR (atypical ADJ3 teratoid ADJ3 rhabdoid ADJ3 tumorigenicity) OR (atypical ADJ3 teratoid ADJ3 rhabdoid ADJ3 tumoral) OR (atypical ADJ3 teratoid ADJ3 rhabdoid ADJ3 tumoropathy) OR (atypical ADJ3 teratoid ADJ3 rhabdoid ADJ3 tumorogenesis) OR (atypical ADJ3 teratoid ADJ3 rhabdoid ADJ3 tumorgenesis) OR (atypical ADJ3 teratoid ADJ3 rhabdoid ADJ3 tumori) OR (atypical ADJ3 teratoid ADJ3 rhabdoid ADJ3 tumorous) OR (atypical ADJ3 teratoid ADJ3 rhabdoid ADJ3 tumoroid) OR (atypical ADJ3 teratoid ADJ3 rhabdoid ADJ3 tumour) OR (atypical ADJ3 teratoid ADJ3 rhabdoid ADJ3 tumours) OR (atypical ADJ3 teratoid ADJ3 rhabdoid ADJ3 tumourigenicity) OR (atypical ADJ3 teratoid ADJ3 rhabdoid ADJ3 tumourous) OR (atypical ADJ3 teratoid ADJ3 rhabdoid ADJ3 tumourigenesis) OR (atypical ADJ3 teratoid ADJ3 rhabdoid ADJ3 tumoural) OR (atypical ADJ3 teratoid ADJ3 rhabdoid ADJ3 carcinoma) OR (atypical ADJ3 teratoid ADJ3 rhabdoid ADJ3 carcinomas) OR (atypical ADJ3 teratoid ADJ3 rhabdoid ADJ3 carcinomatosis) OR (atypical ADJ3 teratoid ADJ3 rhabdoid ADJ3 carcinomatous) OR (atypical ADJ3 teratoid ADJ3 rhabdoid ADJ3 sarcomas) OR (atypical ADJ3 teratoid ADJ3 rhabdoid ADJ3 sarcomatous) OR (atypical ADJ3 teratoid ADJ3 rhabdoid ADJ3 sarcoma) OR (atypical ADJ3 teratoid ADJ3 rhabdoid ADJ3 sarcomatosis) OR (pituitary ADJ3 cancer) OR (pituitary ADJ3 cancerous) OR (pituitary ADJ3 cancers) OR (pituitary ADJ3 cancerogenesis) OR (pituitary ADJ3 canceration) OR (pituitary ADJ3 cancerogenic) OR (pituitary ADJ3 cancerocerebral) OR (pituitary ADJ3 neoplasms) OR (pituitary ADJ3 neoplasm) OR (pituitary ADJ3 neoplasma) OR (pituitary ADJ3 neoplasmic) OR (pituitary ADJ3 neoplasmatic) OR (pituitary ADJ3 neoplasmas) OR (pituitary ADJ3 tumor) OR (pituitary ADJ3 tumors) OR (pituitary ADJ3 tumorlike) OR (pituitary ADJ3 tumoricidal) OR (pituitary ADJ3 tumorigenesis) OR (pituitary ADJ3 tumorectomy) OR (pituitary ADJ3 tumorigenic) OR (pituitary ADJ3 tumorgenicity) OR (pituitary ADJ3 tumorigenicity) OR (pituitary ADJ3 tumoral) OR (pituitary ADJ3 tumoropathy) OR (pituitary ADJ3 tumorogenesis) OR (pituitary ADJ3 tumorgenesis) OR (pituitary ADJ3 tumori) OR (pituitary ADJ3 tumorous) OR (pituitary ADJ3 tumoroid) OR (pituitary ADJ3 tumour) OR (pituitary ADJ3 tumours) OR (pituitary ADJ3 tumourigenicity) OR (pituitary ADJ3 tumourous) OR (pituitary ADJ3 tumourigenesis) OR (pituitary ADJ3 tumoural) OR (pituitary ADJ3 carcinoma) OR (pituitary ADJ3 carcinomas) OR (pituitary ADJ3 carcinomatosis) OR (pituitary ADJ3 carcinomatous) OR (pituitary ADJ3 sarcomas) OR (pituitary ADJ3 sarcomatous) OR (pituitary ADJ3 sarcoma) OR (pituitary ADJ3 sarcomatosis) OR (brain ADJ3 cancer) OR (brain ADJ3 cancerous) OR (brain ADJ3 cancers) OR (brain ADJ3 cancerogenesis) OR (brain ADJ3 canceration) OR (brain ADJ3 cancerogenic) OR (brain ADJ3 cancerocerebral) OR (brain ADJ3 neoplasms) OR (brain ADJ3 neoplasm) OR (brain ADJ3 neoplasma) OR (brain ADJ3 neoplasmic) OR (brain ADJ3 neoplasmatic) OR (brain ADJ3 neoplasmas) OR (brain ADJ3 tumor) OR (brain ADJ3 tumors) OR (brain ADJ3 tumorlike) OR (brain ADJ3 tumoricidal) OR (brain ADJ3 tumorigenesis) OR (brain ADJ3 tumorectomy) OR (brain ADJ3 tumorigenic) OR (brain ADJ3 tumorgenicity) OR (brain ADJ3 tumorigenicity) OR (brain ADJ3 tumoral) OR (brain ADJ3 tumoropathy) OR (brain ADJ3 tumorogenesis) OR (brain ADJ3 tumorgenesis) OR (brain ADJ3 tumori) OR (brain ADJ3 tumorous) OR (brain ADJ3 tumoroid) OR (brain ADJ3 tumour) OR (brain ADJ3 tumours) OR (brain ADJ3 tumourigenicity) OR (brain ADJ3 tumourous) OR (brain ADJ3 tumourigenesis) OR (brain ADJ3 tumoural) OR (brain ADJ3 carcinoma) OR (brain ADJ3 carcinomas) OR (brain ADJ3 carcinomatosis) OR (brain ADJ3 carcinomatous) OR (brain ADJ3 sarcomas) OR (brain ADJ3 sarcomatous) OR (brain ADJ3 sarcoma) OR (brain ADJ3 sarcomatosis) OR (skull ADJ3 base ADJ3 cancer) OR (skull ADJ3 base ADJ3 cancerous) OR (skull ADJ3 base ADJ3 cancers) OR (skull ADJ3 base ADJ3 cancerogenesis) OR (skull ADJ3 base ADJ3 canceration) OR (skull ADJ3 base ADJ3 cancerogenic) OR (skull ADJ3 base ADJ3 cancerocerebral) OR (skull ADJ3 base ADJ3 neoplasms) OR (skull ADJ3 base ADJ3 neoplasm) OR (skull ADJ3 base ADJ3 neoplasma) OR (skull ADJ3 base ADJ3 neoplasmic) OR (skull ADJ3 base ADJ3 neoplasmatic) OR (skull ADJ3 base ADJ3 neoplasmas) OR (skull ADJ3 base ADJ3 tumor) OR (skull ADJ3 base ADJ3 tumors) OR (skull ADJ3 base ADJ3 tumorlike) OR (skull ADJ3 base ADJ3 tumoricidal) OR (skull ADJ3 base ADJ3 tumorigenesis) OR (skull ADJ3 base ADJ3 tumorectomy) OR (skull ADJ3 base ADJ3 tumorigenic) OR (skull ADJ3 base ADJ3 tumorgenicity) OR (skull ADJ3 base ADJ3 tumorigenicity) OR (skull ADJ3 base ADJ3 tumoral) OR (skull ADJ3 base ADJ3 tumoropathy) OR (skull ADJ3 base ADJ3 tumorogenesis) OR (skull ADJ3 base ADJ3 tumorgenesis) OR (skull ADJ3 base ADJ3 tumori) OR (skull ADJ3 base ADJ3 tumorous) OR (skull ADJ3 base ADJ3 tumoroid) OR (skull ADJ3 base ADJ3 tumour) OR (skull ADJ3 base ADJ3 tumours) OR (skull ADJ3 base ADJ3 tumourigenicity) OR (skull ADJ3 base ADJ3 tumourous) OR (skull ADJ3 base ADJ3 tumourigenesis) OR (skull ADJ3 base ADJ3 tumoural) OR (skull ADJ3 base ADJ3 carcinoma) OR (skull ADJ3 base ADJ3 carcinomas) OR (skull ADJ3 base ADJ3 carcinomatosis) OR (skull ADJ3 base ADJ3 carcinomatous) OR (skull ADJ3 base ADJ3 sarcomas) OR (skull ADJ3 base ADJ3 sarcomatous) OR (skull ADJ3 base ADJ3 sarcoma) OR (skull ADJ3 base ADJ3 sarcomatosis) OR (nasopharynx ADJ3 cancer) OR (nasopharynx ADJ3 cancerous) OR (nasopharynx ADJ3 cancers) OR (nasopharynx ADJ3 cancerogenesis) OR (nasopharynx ADJ3 canceration) OR (nasopharynx ADJ3 cancerogenic) OR (nasopharynx ADJ3 cancerocerebral) OR (nasopharynx ADJ3 neoplasms) OR (nasopharynx ADJ3 neoplasm) OR (nasopharynx ADJ3 neoplasma) OR (nasopharynx ADJ3 neoplasmic) OR (nasopharynx ADJ3 neoplasmatic) OR (nasopharynx ADJ3 neoplasmas) OR (nasopharynx ADJ3 tumor) OR (nasopharynx ADJ3 tumors) OR (nasopharynx ADJ3 tumorlike) OR (nasopharynx ADJ3 tumoricidal) OR (nasopharynx ADJ3 tumorigenesis) OR (nasopharynx ADJ3 tumorectomy) OR (nasopharynx ADJ3 tumorigenic) OR (nasopharynx ADJ3 tumorgenicity) OR (nasopharynx ADJ3 tumorigenicity) OR (nasopharynx ADJ3 tumoral) OR (nasopharynx ADJ3 tumoropathy) OR (nasopharynx ADJ3 tumorogenesis) OR (nasopharynx ADJ3 tumorgenesis) OR (nasopharynx ADJ3 tumori) OR (nasopharynx ADJ3 tumorous) OR (nasopharynx ADJ3 tumoroid) OR (nasopharynx ADJ3 tumour) OR (nasopharynx ADJ3 tumours) OR (nasopharynx ADJ3 tumourigenicity) OR (nasopharynx ADJ3 tumourous) OR (nasopharynx ADJ3 tumourigenesis) OR (nasopharynx ADJ3 tumoural) OR (nasopharynx ADJ3 carcinoma) OR (nasopharynx ADJ3 carcinomas) OR (nasopharynx ADJ3 carcinomatosis) OR (nasopharynx ADJ3 carcinomatous) OR (nasopharynx ADJ3 sarcomas) OR (nasopharynx ADJ3 sarcomatous) OR (nasopharynx ADJ3 sarcoma) OR (nasopharynx ADJ3 sarcomatosis) OR (nasopharyngeal ADJ3 cancer) OR (nasopharyngeal ADJ3 cancerous) OR (nasopharyngeal ADJ3 cancers) OR (nasopharyngeal ADJ3 cancerogenesis) OR (nasopharyngeal ADJ3 canceration) OR (nasopharyngeal ADJ3 cancerogenic) OR (nasopharyngeal ADJ3 cancerocerebral) OR (nasopharyngeal ADJ3 neoplasms) OR (nasopharyngeal ADJ3 neoplasm) OR (nasopharyngeal ADJ3 neoplasma) OR (nasopharyngeal ADJ3 neoplasmic) OR (nasopharyngeal ADJ3 neoplasmatic) OR (nasopharyngeal ADJ3 neoplasmas) OR (nasopharyngeal ADJ3 tumor) OR (nasopharyngeal ADJ3 tumors) OR (nasopharyngeal ADJ3 tumorlike) OR (nasopharyngeal ADJ3 tumoricidal) OR (nasopharyngeal ADJ3 tumorigenesis) OR (nasopharyngeal ADJ3 tumorectomy) OR (nasopharyngeal ADJ3 tumorigenic) OR (nasopharyngeal ADJ3 tumorgenicity) OR (nasopharyngeal ADJ3 tumorigenicity) OR (nasopharyngeal ADJ3 tumoral) OR (nasopharyngeal ADJ3 tumoropathy) OR (nasopharyngeal ADJ3 tumorogenesis) OR (nasopharyngeal ADJ3 tumorgenesis) OR (nasopharyngeal ADJ3 tumori) OR (nasopharyngeal ADJ3 tumorous) OR (nasopharyngeal ADJ3 tumoroid) OR (nasopharyngeal ADJ3 tumour) OR (nasopharyngeal ADJ3 tumours) OR (nasopharyngeal ADJ3 tumourigenicity) OR (nasopharyngeal ADJ3 tumourous) OR (nasopharyngeal ADJ3 tumourigenesis) OR (nasopharyngeal ADJ3 tumoural) OR (nasopharyngeal ADJ3 carcinoma) OR (nasopharyngeal ADJ3 carcinomas) OR (nasopharyngeal ADJ3 carcinomatosis) OR (nasopharyngeal ADJ3 carcinomatous) OR (nasopharyngeal ADJ3 sarcomas) OR (nasopharyngeal ADJ3 sarcomatous) OR (nasopharyngeal ADJ3 sarcoma) OR (nasopharyngeal ADJ3 sarcomatosis) OR (meningeal ADJ3 cancer) OR (meningeal ADJ3 cancerous) OR (meningeal ADJ3 cancers) OR (meningeal ADJ3 cancerogenesis) OR (meningeal ADJ3 canceration) OR (meningeal ADJ3 cancerogenic) OR (meningeal ADJ3 cancerocerebral) OR (meningeal ADJ3 neoplasms) OR (meningeal ADJ3 neoplasm) OR (meningeal ADJ3 neoplasma) OR (meningeal ADJ3 neoplasmic) OR (meningeal ADJ3 neoplasmatic) OR (meningeal ADJ3 neoplasmas) OR (meningeal ADJ3 tumor) OR (meningeal ADJ3 tumors) OR (meningeal ADJ3 tumorlike) OR (meningeal ADJ3 tumoricidal) OR (meningeal ADJ3 tumorigenesis) OR (meningeal ADJ3 tumorectomy) OR (meningeal ADJ3 tumorigenic) OR (meningeal ADJ3 tumorgenicity) OR (meningeal ADJ3 tumorigenicity) OR (meningeal ADJ3 tumoral) OR (meningeal ADJ3 tumoropathy) OR (meningeal ADJ3 tumorogenesis) OR (meningeal ADJ3 tumorgenesis) OR (meningeal ADJ3 tumori) OR (meningeal ADJ3 tumorous) OR (meningeal ADJ3 tumoroid) OR (meningeal ADJ3 tumour) OR (meningeal ADJ3 tumours) OR (meningeal ADJ3 tumourigenicity) OR (meningeal ADJ3 tumourous) OR (meningeal ADJ3 tumourigenesis) OR (meningeal ADJ3 tumoural) OR (meningeal ADJ3 carcinoma) OR (meningeal ADJ3 carcinomas) OR (meningeal ADJ3 carcinomatosis) OR (meningeal ADJ3 carcinomatous) OR (meningeal ADJ3 sarcomas) OR (meningeal ADJ3 sarcomatous) OR (meningeal ADJ3 sarcoma) OR (meningeal ADJ3 sarcomatosis) OR (sinus ADJ3 cancer) OR (sinus ADJ3 cancerous) OR (sinus ADJ3 cancers) OR (sinus ADJ3 cancerogenesis) OR (sinus ADJ3 canceration) OR (sinus ADJ3 cancerogenic) OR (sinus ADJ3 cancerocerebral) OR (sinus ADJ3 neoplasms) OR (sinus ADJ3 neoplasm) OR (sinus ADJ3 neoplasma) OR (sinus ADJ3 neoplasmic) OR (sinus ADJ3 neoplasmatic) OR (sinus ADJ3 neoplasmas) OR (sinus ADJ3 tumor) OR (sinus ADJ3 tumors) OR (sinus ADJ3 tumorlike) OR (sinus ADJ3 tumoricidal) OR (sinus ADJ3 tumorigenesis) OR (sinus ADJ3 tumorectomy) OR (sinus ADJ3 tumorigenic) OR (sinus ADJ3 tumorgenicity) OR (sinus ADJ3 tumorigenicity) OR (sinus ADJ3 tumoral) OR (sinus ADJ3 tumoropathy) OR (sinus ADJ3 tumorogenesis) OR (sinus ADJ3 tumorgenesis) OR (sinus ADJ3 tumori) OR (sinus ADJ3 tumorous) OR (sinus ADJ3 tumoroid) OR (sinus ADJ3 tumour) OR (sinus ADJ3 tumours) OR (sinus ADJ3 tumourigenicity) OR (sinus ADJ3 tumourous) OR (sinus ADJ3 tumourigenesis) OR (sinus ADJ3 tumoural) OR (sinus ADJ3 carcinoma) OR (sinus ADJ3 carcinomas) OR (sinus ADJ3 carcinomatosis) OR (sinus ADJ3 carcinomatous) OR (sinus ADJ3 sarcomas) OR (sinus ADJ3 sarcomatous) OR (sinus ADJ3 sarcoma) OR (sinus ADJ3 sarcomatosis) OR (intracranial ADJ3 cancer) OR (intracranial ADJ3 cancerous) OR (intracranial ADJ3 cancers) OR (intracranial ADJ3 cancerogenesis) OR (intracranial ADJ3 canceration) OR (intracranial ADJ3 cancerogenic) OR (intracranial ADJ3 cancerocerebral) OR (intracranial ADJ3 neoplasms) OR (intracranial ADJ3 neoplasm) OR (intracranial ADJ3 neoplasma) OR (intracranial ADJ3 neoplasmic) OR (intracranial ADJ3 neoplasmatic) OR (intracranial ADJ3 neoplasmas) OR (intracranial ADJ3 tumor) OR (intracranial ADJ3 tumors) OR (intracranial ADJ3 tumorlike) OR (intracranial ADJ3 tumoricidal) OR (intracranial ADJ3 tumorigenesis) OR (intracranial ADJ3 tumorectomy) OR (intracranial ADJ3 tumorigenic) OR (intracranial ADJ3 tumorgenicity) OR (intracranial ADJ3 tumorigenicity) OR (intracranial ADJ3 tumoral) OR (intracranial ADJ3 tumoropathy) OR (intracranial ADJ3 tumorogenesis) OR (intracranial ADJ3 tumorgenesis) OR (intracranial ADJ3 tumori) OR (intracranial ADJ3 tumorous) OR (intracranial ADJ3 tumoroid) OR (intracranial ADJ3 tumour) OR (intracranial ADJ3 tumours) OR (intracranial ADJ3 tumourigenicity) OR (intracranial ADJ3 tumourous) OR (intracranial ADJ3 tumourigenesis) OR (intracranial ADJ3 tumoural) OR (intracranial ADJ3 carcinoma) OR (intracranial ADJ3 carcinomas) OR (intracranial ADJ3 carcinomatosis) OR (intracranial ADJ3 carcinomatous) OR (intracranial ADJ3 sarcomas) OR (intracranial ADJ3 sarcomatous) OR (intracranial ADJ3 sarcoma) OR (intracranial ADJ3 sarcomatosis) OR (cerebral ADJ3 cancer) OR (cerebral ADJ3 cancerous) OR (cerebral ADJ3 cancers) OR (cerebral ADJ3 cancerogenesis) OR (cerebral ADJ3 canceration) OR (cerebral ADJ3 cancerogenic) OR (cerebral ADJ3 cancerocerebral) OR (cerebral ADJ3 neoplasms) OR (cerebral ADJ3 neoplasm) OR (cerebral ADJ3 neoplasma) OR (cerebral ADJ3 neoplasmic) OR (cerebral ADJ3 neoplasmatic) OR (cerebral ADJ3 neoplasmas) OR (cerebral ADJ3 tumor) OR (cerebral ADJ3 tumors) OR (cerebral ADJ3 tumorlike) OR (cerebral ADJ3 tumoricidal) OR (cerebral ADJ3 tumorigenesis) OR (cerebral ADJ3 tumorectomy) OR (cerebral ADJ3 tumorigenic) OR (cerebral ADJ3 tumorgenicity) OR (cerebral ADJ3 tumorigenicity) OR (cerebral ADJ3 tumoral) OR (cerebral ADJ3 tumoropathy) OR (cerebral ADJ3 tumorogenesis) OR (cerebral ADJ3 tumorgenesis) OR (cerebral ADJ3 tumori) OR (cerebral ADJ3 tumorous) OR (cerebral ADJ3 tumoroid) OR (cerebral ADJ3 tumour) OR (cerebral ADJ3 tumours) OR (cerebral ADJ3 tumourigenicity) OR (cerebral ADJ3 tumourous) OR (cerebral ADJ3 tumourigenesis) OR (cerebral ADJ3 tumoural) OR (cerebral ADJ3 carcinoma) OR (cerebral ADJ3 carcinomas) OR (cerebral ADJ3 carcinomatosis) OR (cerebral ADJ3 carcinomatous) OR (cerebral ADJ3 sarcomas) OR (cerebral ADJ3 sarcomatous) OR (cerebral ADJ3 sarcoma) OR (cerebral ADJ3 sarcomatosis) OR (subtentorial ADJ3 cancer) OR (subtentorial ADJ3 cancerous) OR (subtentorial ADJ3 cancers) OR (subtentorial ADJ3 cancerogenesis) OR (subtentorial ADJ3 canceration) OR (subtentorial ADJ3 cancerogenic) OR (subtentorial ADJ3 cancerocerebral) OR (subtentorial ADJ3 neoplasms) OR (subtentorial ADJ3 neoplasm) OR (subtentorial ADJ3 neoplasma) OR (subtentorial ADJ3 neoplasmic) OR (subtentorial ADJ3 neoplasmatic) OR (subtentorial ADJ3 neoplasmas) OR (subtentorial ADJ3 tumor) OR (subtentorial ADJ3 tumors) OR (subtentorial ADJ3 tumorlike) OR (subtentorial ADJ3 tumoricidal) OR (subtentorial ADJ3 tumorigenesis) OR (subtentorial ADJ3 tumorectomy) OR (subtentorial ADJ3 tumorigenic) OR (subtentorial ADJ3 tumorgenicity) OR (subtentorial ADJ3 tumorigenicity) OR (subtentorial ADJ3 tumoral) OR (subtentorial ADJ3 tumoropathy) OR (subtentorial ADJ3 tumorogenesis) OR (subtentorial ADJ3 tumorgenesis) OR (subtentorial ADJ3 tumori) OR (subtentorial ADJ3 tumorous) OR (subtentorial ADJ3 tumoroid) OR (subtentorial ADJ3 tumour) OR (subtentorial ADJ3 tumours) OR (subtentorial ADJ3 tumourigenicity) OR (subtentorial ADJ3 tumourous) OR (subtentorial ADJ3 tumourigenesis) OR (subtentorial ADJ3 tumoural) OR (subtentorial ADJ3 carcinoma) OR (subtentorial ADJ3 carcinomas) OR (subtentorial ADJ3 carcinomatosis) OR (subtentorial ADJ3 carcinomatous) OR (subtentorial ADJ3 sarcomas) OR (subtentorial ADJ3 sarcomatous) OR (subtentorial ADJ3 sarcoma) OR (subtentorial ADJ3 sarcomatosis) OR (infratentorial ADJ3 cancer) OR (infratentorial ADJ3 cancerous) OR (infratentorial ADJ3 cancers) OR (infratentorial ADJ3 cancerogenesis) OR (infratentorial ADJ3 canceration) OR (infratentorial ADJ3 cancerogenic) OR (infratentorial ADJ3 cancerocerebral) OR (infratentorial ADJ3 neoplasms) OR (infratentorial ADJ3 neoplasm) OR (infratentorial ADJ3 neoplasma) OR (infratentorial ADJ3 neoplasmic) OR (infratentorial ADJ3 neoplasmatic) OR (infratentorial ADJ3 neoplasmas) OR (infratentorial ADJ3 tumor) OR (infratentorial ADJ3 tumors) OR (infratentorial ADJ3 tumorlike) OR (infratentorial ADJ3 tumoricidal) OR (infratentorial ADJ3 tumorigenesis) OR (infratentorial ADJ3 tumorectomy) OR (infratentorial ADJ3 tumorigenic) OR (infratentorial ADJ3 tumorgenicity) OR (infratentorial ADJ3 tumorigenicity) OR (infratentorial ADJ3 tumoral) OR (infratentorial ADJ3 tumoropathy) OR (infratentorial ADJ3 tumorogenesis) OR (infratentorial ADJ3 tumorgenesis) OR (infratentorial ADJ3 tumori) OR (infratentorial ADJ3 tumorous) OR (infratentorial ADJ3 tumoroid) OR (infratentorial ADJ3 tumour) OR (infratentorial ADJ3 tumours) OR (infratentorial ADJ3 tumourigenicity) OR (infratentorial ADJ3 tumourous) OR (infratentorial ADJ3 tumourigenesis) OR (infratentorial ADJ3 tumoural) OR (infratentorial ADJ3 carcinoma) OR (infratentorial ADJ3 carcinomas) OR (infratentorial ADJ3 carcinomatosis) OR (infratentorial ADJ3 carcinomatous) OR (infratentorial ADJ3 sarcomas) OR (infratentorial ADJ3 sarcomatous) OR (infratentorial ADJ3 sarcoma) OR (infratentorial ADJ3 sarcomatosis) OR (supratentorial ADJ3 cancer) OR (supratentorial ADJ3 cancerous) OR (supratentorial ADJ3 cancers) OR (supratentorial ADJ3 cancerogenesis) OR (supratentorial ADJ3 canceration) OR (supratentorial ADJ3 cancerogenic) OR (supratentorial ADJ3 cancerocerebral) OR (supratentorial ADJ3 neoplasms) OR (supratentorial ADJ3 neoplasm) OR (supratentorial ADJ3 neoplasma) OR (supratentorial ADJ3 neoplasmic) OR (supratentorial ADJ3 neoplasmatic) OR (supratentorial ADJ3 neoplasmas) OR (supratentorial ADJ3 tumor) OR (supratentorial ADJ3 tumors) OR (supratentorial ADJ3 tumorlike) OR (supratentorial ADJ3 tumoricidal) OR (supratentorial ADJ3 tumorigenesis) OR (supratentorial ADJ3 tumorectomy) OR (supratentorial ADJ3 tumorigenic) OR (supratentorial ADJ3 tumorgenicity) OR (supratentorial ADJ3 tumorigenicity) OR (supratentorial ADJ3 tumoral) OR (supratentorial ADJ3 tumoropathy) OR (supratentorial ADJ3 tumorogenesis) OR (supratentorial ADJ3 tumorgenesis) OR (supratentorial ADJ3 tumori) OR (supratentorial ADJ3 tumorous) OR (supratentorial ADJ3 tumoroid) OR (supratentorial ADJ3 tumour) OR (supratentorial ADJ3 tumours) OR (supratentorial ADJ3 tumourigenicity) OR (supratentorial ADJ3 tumourous) OR (supratentorial ADJ3 tumourigenesis) OR (supratentorial ADJ3 tumoural) OR (supratentorial ADJ3 carcinoma) OR (supratentorial ADJ3 carcinomas) OR (supratentorial ADJ3 carcinomatosis) OR (supratentorial ADJ3 carcinomatous) OR (supratentorial ADJ3 sarcomas) OR (supratentorial ADJ3 sarcomatous) OR (supratentorial ADJ3 sarcoma) OR (supratentorial ADJ3 sarcomatosis) OR (pituitary ADJ3 gland ADJ3 cancer) OR (pituitary ADJ3 gland ADJ3 cancerous) OR (pituitary ADJ3 gland ADJ3 cancers) OR (pituitary ADJ3 gland ADJ3 cancerogenesis) OR (pituitary ADJ3 gland ADJ3 canceration) OR (pituitary ADJ3 gland ADJ3 cancerogenic) OR (pituitary ADJ3 gland ADJ3 cancerocerebral) OR (pituitary ADJ3 gland ADJ3 neoplasms) OR (pituitary ADJ3 gland ADJ3 neoplasm) OR (pituitary ADJ3 gland ADJ3 neoplasma) OR (pituitary ADJ3 gland ADJ3 neoplasmic) OR (pituitary ADJ3 gland ADJ3 neoplasmatic) OR (pituitary ADJ3 gland ADJ3 neoplasmas) OR (pituitary ADJ3 gland ADJ3 tumor) OR (pituitary ADJ3 gland ADJ3 tumors) OR (pituitary ADJ3 gland ADJ3 tumorlike) OR (pituitary ADJ3 gland ADJ3 tumoricidal) OR (pituitary ADJ3 gland ADJ3 tumorigenesis) OR (pituitary ADJ3 gland ADJ3 tumorectomy) OR (pituitary ADJ3 gland ADJ3 tumorigenic) OR (pituitary ADJ3 gland ADJ3 tumorgenicity) OR (pituitary ADJ3 gland ADJ3 tumorigenicity) OR (pituitary ADJ3 gland ADJ3 tumoral) OR (pituitary ADJ3 gland ADJ3 tumoropathy) OR (pituitary ADJ3 gland ADJ3 tumorogenesis) OR (pituitary ADJ3 gland ADJ3 tumorgenesis) OR (pituitary ADJ3 gland ADJ3 tumori) OR (pituitary ADJ3 gland ADJ3 tumorous) OR (pituitary ADJ3 gland ADJ3 tumoroid) OR (pituitary ADJ3 gland ADJ3 tumour) OR (pituitary ADJ3 gland ADJ3 tumours) OR (pituitary ADJ3 gland ADJ3 tumourigenicity) OR (pituitary ADJ3 gland ADJ3 tumourous) OR (pituitary ADJ3 gland ADJ3 tumourigenesis) OR (pituitary ADJ3 gland ADJ3 tumoural) OR (pituitary ADJ3 gland ADJ3 carcinoma) OR (pituitary ADJ3 gland ADJ3 carcinomas) OR (pituitary ADJ3 gland ADJ3 carcinomatosis) OR (pituitary ADJ3 gland ADJ3 carcinomatous) OR (pituitary ADJ3 gland ADJ3 sarcomas) OR (pituitary ADJ3 gland ADJ3 sarcomatous) OR (pituitary ADJ3 gland ADJ3 sarcoma) OR (pituitary ADJ3 gland ADJ3 sarcomatosis) OR (hypophyseal ADJ3 cancer) OR (hypophyseal ADJ3 cancerous) OR (hypophyseal ADJ3 cancers) OR (hypophyseal ADJ3 cancerogenesis) OR (hypophyseal ADJ3 canceration) OR (hypophyseal ADJ3 cancerogenic) OR (hypophyseal ADJ3 cancerocerebral) OR (hypophyseal ADJ3 neoplasms) OR (hypophyseal ADJ3 neoplasm) OR (hypophyseal ADJ3 neoplasma) OR (hypophyseal ADJ3 neoplasmic) OR (hypophyseal ADJ3 neoplasmatic) OR (hypophyseal ADJ3 neoplasmas) OR (hypophyseal ADJ3 tumor) OR (hypophyseal ADJ3 tumors) OR (hypophyseal ADJ3 tumorlike) OR (hypophyseal ADJ3 tumoricidal) OR (hypophyseal ADJ3 tumorigenesis) OR (hypophyseal ADJ3 tumorectomy) OR (hypophyseal ADJ3 tumorigenic) OR (hypophyseal ADJ3 tumorgenicity) OR (hypophyseal ADJ3 tumorigenicity) OR (hypophyseal ADJ3 tumoral) OR (hypophyseal ADJ3 tumoropathy) OR (hypophyseal ADJ3 tumorogenesis) OR (hypophyseal ADJ3 tumorgenesis) OR (hypophyseal ADJ3 tumori) OR (hypophyseal ADJ3 tumorous) OR (hypophyseal ADJ3 tumoroid) OR (hypophyseal ADJ3 tumour) OR (hypophyseal ADJ3 tumours) OR (hypophyseal ADJ3 tumourigenicity) OR (hypophyseal ADJ3 tumourous) OR (hypophyseal ADJ3 tumourigenesis) OR (hypophyseal ADJ3 tumoural) OR (hypophyseal ADJ3 carcinoma) OR (hypophyseal ADJ3 carcinomas) OR (hypophyseal ADJ3 carcinomatosis) OR (hypophyseal ADJ3 carcinomatous) OR (hypophyseal ADJ3 sarcomas) OR (hypophyseal ADJ3 sarcomatous) OR (hypophyseal ADJ3 sarcoma) OR (hypophyseal ADJ3 sarcomatosis) OR (hypophysis ADJ3 cancer) OR (hypophysis ADJ3 cancerous) OR (hypophysis ADJ3 cancers) OR (hypophysis ADJ3 cancerogenesis) OR (hypophysis ADJ3 canceration) OR (hypophysis ADJ3 cancerogenic) OR (hypophysis ADJ3 cancerocerebral) OR (hypophysis ADJ3 neoplasms) OR (hypophysis ADJ3 neoplasm) OR (hypophysis ADJ3 neoplasma) OR (hypophysis ADJ3 neoplasmic) OR (hypophysis ADJ3 neoplasmatic) OR (hypophysis ADJ3 neoplasmas) OR (hypophysis ADJ3 tumor) OR (hypophysis ADJ3 tumors) OR (hypophysis ADJ3 tumorlike) OR (hypophysis ADJ3 tumoricidal) OR (hypophysis ADJ3 tumorigenesis) OR (hypophysis ADJ3 tumorectomy) OR (hypophysis ADJ3 tumorigenic) OR (hypophysis ADJ3 tumorgenicity) OR (hypophysis ADJ3 tumorigenicity) OR (hypophysis ADJ3 tumoral) OR (hypophysis ADJ3 tumoropathy) OR (hypophysis ADJ3 tumorogenesis) OR (hypophysis ADJ3 tumorgenesis) OR (hypophysis ADJ3 tumori) OR (hypophysis ADJ3 tumorous) OR (hypophysis ADJ3 tumoroid) OR (hypophysis ADJ3 tumour) OR (hypophysis ADJ3 tumours) OR (hypophysis ADJ3 tumourigenicity) OR (hypophysis ADJ3 tumourous) OR (hypophysis ADJ3 tumourigenesis) OR (hypophysis ADJ3 tumoural) OR (hypophysis ADJ3 carcinoma) OR (hypophysis ADJ3 carcinomas) OR (hypophysis ADJ3 carcinomatosis) OR (hypophysis ADJ3 carcinomatous) OR (hypophysis ADJ3 sarcomas) OR (hypophysis ADJ3 sarcomatous) OR (hypophysis ADJ3 sarcoma) OR (hypophysis ADJ3 sarcomatosis) OR (cranial ADJ3 base ADJ3 cancer) OR (cranial ADJ3 base ADJ3 cancerous) OR (cranial ADJ3 base ADJ3 cancers) OR (cranial ADJ3 base ADJ3 cancerogenesis) OR (cranial ADJ3 base ADJ3 canceration) OR (cranial ADJ3 base ADJ3 cancerogenic) OR (cranial ADJ3 base ADJ3 cancerocerebral) OR (cranial ADJ3 base ADJ3 neoplasms) OR (cranial ADJ3 base ADJ3 neoplasm) OR (cranial ADJ3 base ADJ3 neoplasma) OR (cranial ADJ3 base ADJ3 neoplasmic) OR (cranial ADJ3 base ADJ3 neoplasmatic) OR (cranial ADJ3 base ADJ3 neoplasmas) OR (cranial ADJ3 base ADJ3 tumor) OR (cranial ADJ3 base ADJ3 tumors) OR (cranial ADJ3 base ADJ3 tumorlike) OR (cranial ADJ3 base ADJ3 tumoricidal) OR (cranial ADJ3 base ADJ3 tumorigenesis) OR (cranial ADJ3 base ADJ3 tumorectomy) OR (cranial ADJ3 base ADJ3 tumorigenic) OR (cranial ADJ3 base ADJ3 tumorgenicity) OR (cranial ADJ3 base ADJ3 tumorigenicity) OR (cranial ADJ3 base ADJ3 tumoral) OR (cranial ADJ3 base ADJ3 tumoropathy) OR (cranial ADJ3 base ADJ3 tumorogenesis) OR (cranial ADJ3 base ADJ3 tumorgenesis) OR (cranial ADJ3 base ADJ3 tumori) OR (cranial ADJ3 base ADJ3 tumorous) OR (cranial ADJ3 base ADJ3 tumoroid) OR (cranial ADJ3 base ADJ3 tumour) OR (cranial ADJ3 base ADJ3 tumours) OR (cranial ADJ3 base ADJ3 tumourigenicity) OR (cranial ADJ3 base ADJ3 tumourous) OR (cranial ADJ3 base ADJ3 tumourigenesis) OR (cranial ADJ3 base ADJ3 tumoural) OR (cranial ADJ3 base ADJ3 carcinoma) OR (cranial ADJ3 base ADJ3 carcinomas) OR (cranial ADJ3 base ADJ3 carcinomatosis) OR (cranial ADJ3 base ADJ3 carcinomatous) OR (cranial ADJ3 base ADJ3 sarcomas) OR (cranial ADJ3 base ADJ3 sarcomatous) OR (cranial ADJ3 base ADJ3 sarcoma) OR (cranial ADJ3 base ADJ3 sarcomatosis) OR (craniobasal ADJ3 cancer) OR (craniobasal ADJ3 cancerous) OR (craniobasal ADJ3 cancers) OR (craniobasal ADJ3 cancerogenesis) OR (craniobasal ADJ3 canceration) OR (craniobasal ADJ3 cancerogenic) OR (craniobasal ADJ3 cancerocerebral) OR (craniobasal ADJ3 neoplasms) OR (craniobasal ADJ3 neoplasm) OR (craniobasal ADJ3 neoplasma) OR (craniobasal ADJ3 neoplasmic) OR (craniobasal ADJ3 neoplasmatic) OR (craniobasal ADJ3 neoplasmas) OR (craniobasal ADJ3 tumor) OR (craniobasal ADJ3 tumors) OR (craniobasal ADJ3 tumorlike) OR (craniobasal ADJ3 tumoricidal) OR (craniobasal ADJ3 tumorigenesis) OR (craniobasal ADJ3 tumorectomy) OR (craniobasal ADJ3 tumorigenic) OR (craniobasal ADJ3 tumorgenicity) OR (craniobasal ADJ3 tumorigenicity) OR (craniobasal ADJ3 tumoral) OR (craniobasal ADJ3 tumoropathy) OR (craniobasal ADJ3 tumorogenesis) OR (craniobasal ADJ3 tumorgenesis) OR (craniobasal ADJ3 tumori) OR (craniobasal ADJ3 tumorous) OR (craniobasal ADJ3 tumoroid) OR (craniobasal ADJ3 tumour) OR (craniobasal ADJ3 tumours) OR (craniobasal ADJ3 tumourigenicity) OR (craniobasal ADJ3 tumourous) OR (craniobasal ADJ3 tumourigenesis) OR (craniobasal ADJ3 tumoural) OR (craniobasal ADJ3 carcinoma) OR (craniobasal ADJ3 carcinomas) OR (craniobasal ADJ3 carcinomatosis) OR (craniobasal ADJ3 carcinomatous) OR (craniobasal ADJ3 sarcomas) OR (craniobasal ADJ3 sarcomatous) OR (craniobasal ADJ3 sarcoma) OR (craniobasal ADJ3 sarcomatosis) OR (germinal ADJ3 cancer) OR (germinal ADJ3 cancerous) OR (germinal ADJ3 cancers) OR (germinal ADJ3 cancerogenesis) OR (germinal ADJ3 canceration) OR (germinal ADJ3 cancerogenic) OR (germinal ADJ3 cancerocerebral) OR (germinal ADJ3 neoplasms) OR (germinal ADJ3 neoplasm) OR (germinal ADJ3 neoplasma) OR (germinal ADJ3 neoplasmic) OR (germinal ADJ3 neoplasmatic) OR (germinal ADJ3 neoplasmas) OR (germinal ADJ3 tumor) OR (germinal ADJ3 tumors) OR (germinal ADJ3 tumorlike) OR (germinal ADJ3 tumoricidal) OR (germinal ADJ3 tumorigenesis) OR (germinal ADJ3 tumorectomy) OR (germinal ADJ3 tumorigenic) OR (germinal ADJ3 tumorgenicity) OR (germinal ADJ3 tumorigenicity) OR (germinal ADJ3 tumoral) OR (germinal ADJ3 tumoropathy) OR (germinal ADJ3 tumorogenesis) OR (germinal ADJ3 tumorgenesis) OR (germinal ADJ3 tumori) OR (germinal ADJ3 tumorous) OR (germinal ADJ3 tumoroid) OR (germinal ADJ3 tumour) OR (germinal ADJ3 tumours) OR (germinal ADJ3 tumourigenicity) OR (germinal ADJ3 tumourous) OR (germinal ADJ3 tumourigenesis) OR (germinal ADJ3 tumoural) OR (germinal ADJ3 carcinoma) OR (germinal ADJ3 carcinomas) OR (germinal ADJ3 carcinomatosis) OR (germinal ADJ3 carcinomatous) OR (germinal ADJ3 sarcomas) OR (germinal ADJ3 sarcomatous) OR (germinal ADJ3 sarcoma) OR (germinal ADJ3 sarcomatosis) OR (craniopharyngeal ADJ3 cancer) OR (craniopharyngeal ADJ3 cancerous) OR (craniopharyngeal ADJ3 cancers) OR (craniopharyngeal ADJ3 cancerogenesis) OR (craniopharyngeal ADJ3 canceration) OR (craniopharyngeal ADJ3 cancerogenic) OR (craniopharyngeal ADJ3 cancerocerebral) OR (craniopharyngeal ADJ3 neoplasms) OR (craniopharyngeal ADJ3 neoplasm) OR (craniopharyngeal ADJ3 neoplasma) OR (craniopharyngeal ADJ3 neoplasmic) OR (craniopharyngeal ADJ3 neoplasmatic) OR (craniopharyngeal ADJ3 neoplasmas) OR (craniopharyngeal ADJ3 tumor) OR (craniopharyngeal ADJ3 tumors) OR (craniopharyngeal ADJ3 tumorlike) OR (craniopharyngeal ADJ3 tumoricidal) OR (craniopharyngeal ADJ3 tumorigenesis) OR (craniopharyngeal ADJ3 tumorectomy) OR (craniopharyngeal ADJ3 tumorigenic) OR (craniopharyngeal ADJ3 tumorgenicity) OR (craniopharyngeal ADJ3 tumorigenicity) OR (craniopharyngeal ADJ3 tumoral) OR (craniopharyngeal ADJ3 tumoropathy) OR (craniopharyngeal ADJ3 tumorogenesis) OR (craniopharyngeal ADJ3 tumorgenesis) OR (craniopharyngeal ADJ3 tumori) OR (craniopharyngeal ADJ3 tumorous) OR (craniopharyngeal ADJ3 tumoroid) OR (craniopharyngeal ADJ3 tumour) OR (craniopharyngeal ADJ3 tumours) OR (craniopharyngeal ADJ3 tumourigenicity) OR (craniopharyngeal ADJ3 tumourous) OR (craniopharyngeal ADJ3 tumourigenesis) OR (craniopharyngeal ADJ3 tumoural) OR (craniopharyngeal ADJ3 carcinoma) OR (craniopharyngeal ADJ3 carcinomas) OR (craniopharyngeal ADJ3 carcinomatosis) OR (craniopharyngeal ADJ3 carcinomatous) OR (craniopharyngeal ADJ3 sarcomas) OR (craniopharyngeal ADJ3 sarcomatous) OR (craniopharyngeal ADJ3 sarcoma) OR (craniopharyngeal ADJ3 sarcomatosis) OR (glial ADJ3 cancer) OR (glial ADJ3 cancerous) OR (glial ADJ3 cancers) OR (glial ADJ3 cancerogenesis) OR (glial ADJ3 canceration) OR (glial ADJ3 cancerogenic) OR (glial ADJ3 cancerocerebral) OR (glial ADJ3 neoplasms) OR (glial ADJ3 neoplasm) OR (glial ADJ3 neoplasma) OR (glial ADJ3 neoplasmic) OR (glial ADJ3 neoplasmatic) OR (glial ADJ3 neoplasmas) OR (glial ADJ3 tumor) OR (glial ADJ3 tumors) OR (glial ADJ3 tumorlike) OR (glial ADJ3 tumoricidal) OR (glial ADJ3 tumorigenesis) OR (glial ADJ3 tumorectomy) OR (glial ADJ3 tumorigenic) OR (glial ADJ3 tumorgenicity) OR (glial ADJ3 tumorigenicity) OR (glial ADJ3 tumoral) OR (glial ADJ3 tumoropathy) OR (glial ADJ3 tumorogenesis) OR (glial ADJ3 tumorgenesis) OR (glial ADJ3 tumori) OR (glial ADJ3 tumorous) OR (glial ADJ3 tumoroid) OR (glial ADJ3 tumour) OR (glial ADJ3 tumours) OR (glial ADJ3 tumourigenicity) OR (glial ADJ3 tumourous) OR (glial ADJ3 tumourigenesis) OR (glial ADJ3 tumoural) OR (glial ADJ3 carcinoma) OR (glial ADJ3 carcinomas) OR (glial ADJ3 carcinomatosis) OR (glial ADJ3 carcinomatous) OR (glial ADJ3 sarcomas) OR (glial ADJ3 sarcomatous) OR (glial ADJ3 sarcoma) OR (glial ADJ3 sarcomatosis) OR (ependymal ADJ3 cancer) OR (ependymal ADJ3 cancerous) OR (ependymal ADJ3 cancers) OR (ependymal ADJ3 cancerogenesis) OR (ependymal ADJ3 canceration) OR (ependymal ADJ3 cancerogenic) OR (ependymal ADJ3 cancerocerebral) OR (ependymal ADJ3 neoplasms) OR (ependymal ADJ3 neoplasm) OR (ependymal ADJ3 neoplasma) OR (ependymal ADJ3 neoplasmic) OR (ependymal ADJ3 neoplasmatic) OR (ependymal ADJ3 neoplasmas) OR (ependymal ADJ3 tumor) OR (ependymal ADJ3 tumors) OR (ependymal ADJ3 tumorlike) OR (ependymal ADJ3 tumoricidal) OR (ependymal ADJ3 tumorigenesis) OR (ependymal ADJ3 tumorectomy) OR (ependymal ADJ3 tumorigenic) OR (ependymal ADJ3 tumorgenicity) OR (ependymal ADJ3 tumorigenicity) OR (ependymal ADJ3 tumoral) OR (ependymal ADJ3 tumoropathy) OR (ependymal ADJ3 tumorogenesis) OR (ependymal ADJ3 tumorgenesis) OR (ependymal ADJ3 tumori) OR (ependymal ADJ3 tumorous) OR (ependymal ADJ3 tumoroid) OR (ependymal ADJ3 tumour) OR (ependymal ADJ3 tumours) OR (ependymal ADJ3 tumourigenicity) OR (ependymal ADJ3 tumourous) OR (ependymal ADJ3 tumourigenesis) OR (ependymal ADJ3 tumoural) OR (ependymal ADJ3 carcinoma) OR (ependymal ADJ3 carcinomas) OR (ependymal ADJ3 carcinomatosis) OR (ependymal ADJ3 carcinomatous) OR (ependymal ADJ3 sarcomas) OR (ependymal ADJ3 sarcomatous) OR (ependymal ADJ3 sarcoma) OR (ependymal ADJ3 sarcomatosis)))).ti,ab,kf.) **AND**

(exp "Necrosis"/ **OR** exp "Radiation Injuries"/ **OR** exp "Abnormalities, Radiation-Induced"/ **OR** "Diagnostic imaging".fs. **OR** ("necros*" **OR** "radionecros*" **OR** "radio-necros*" **OR** "pseudoprogress*" **OR** "image change*" **OR** "imaging change*" **OR** "radiographic change*" **OR** "radiological change*" **OR** "radiographic change*" **OR** "Radiation Injur*" **OR** "Radiation Induced Abnormalit*" **OR** "radiation induced contrast enhancement*" **OR** "brain stem injur*" **OR** "brainstem injur*" **OR** "RIBI" **OR** "CEBL" **OR** "RICE" **OR** (((contrast-enhanced ADJ3 lesions) OR (contrast-enhanced ADJ3 lesion) OR (contrast-enhanced ADJ3 lesioned) OR (contrast-enhanced ADJ3 lesional) OR (contrast-enhanced ADJ3 lesionectomy) OR (contrast-enhanced ADJ3 lesionography) OR (contrast-enhanced ADJ3 lesionsuvmax) OR (contrast-enhanced ADJ3 lesionsuvpeak) OR (contrast-enhanced ADJ3 lesioning) OR (contrast-enhanced ADJ3 lesiones) OR (contrast-enhanced ADJ3 lesionary) OR (contrast-enhanced ADJ3 lesionsaccular) OR (contrast-enhanced ADJ3 leasions) OR (contrast-enhancement ADJ3 lesions) OR (contrast-enhancement ADJ3 lesion) OR (contrast-enhancement ADJ3 lesioned) OR (contrast-enhancement ADJ3 lesional) OR (contrast-enhancement ADJ3 lesionectomy) OR (contrast-enhancement ADJ3 lesionography) OR (contrast-enhancement ADJ3 lesionsuvmax) OR (contrast-enhancement ADJ3 lesionsuvpeak) OR (contrast-enhancement ADJ3 lesioning) OR (contrast-enhancement ADJ3 lesiones) OR (contrast-enhancement ADJ3 lesionary) OR (contrast-enhancement ADJ3 lesionsaccular) OR (contrast-enhancement ADJ3 leasions) OR (contrast-enhancing ADJ3 lesions) OR (contrast-enhancing ADJ3 lesion) OR (contrast-enhancing ADJ3 lesioned) OR (contrast-enhancing ADJ3 lesional) OR (contrast-enhancing ADJ3 lesionectomy) OR (contrast-enhancing ADJ3 lesionography) OR (contrast-enhancing ADJ3 lesionsuvmax) OR (contrast-enhancing ADJ3 lesionsuvpeak) OR (contrast-enhancing ADJ3 lesioning) OR (contrast-enhancing ADJ3 lesiones) OR (contrast-enhancing ADJ3 lesionary) OR (contrast-enhancing ADJ3 lesionsaccular) OR (contrast-enhancing ADJ3 leasions) OR (contrast-enhancer ADJ3 lesions) OR (contrast-enhancer ADJ3 lesion) OR (contrast-enhancer ADJ3 lesioned) OR (contrast-enhancer ADJ3 lesional) OR (contrast-enhancer ADJ3 lesionectomy) OR (contrast-enhancer ADJ3 lesionography) OR (contrast-enhancer ADJ3 lesionsuvmax) OR (contrast-enhancer ADJ3 lesionsuvpeak) OR (contrast-enhancer ADJ3 lesioning) OR (contrast-enhancer ADJ3 lesiones) OR (contrast-enhancer ADJ3 lesionary) OR (contrast-enhancer ADJ3 lesionsaccular) OR (contrast-enhancer ADJ3 leasions) OR (contrast-enhancers ADJ3 lesions) OR (contrast-enhancers ADJ3 lesion) OR (contrast-enhancers ADJ3 lesioned) OR (contrast-enhancers ADJ3 lesional) OR (contrast-enhancers ADJ3 lesionectomy) OR (contrast-enhancers ADJ3 lesionography) OR (contrast-enhancers ADJ3 lesionsuvmax) OR (contrast-enhancers ADJ3 lesionsuvpeak) OR (contrast-enhancers ADJ3 lesioning) OR (contrast-enhancers ADJ3 lesiones) OR (contrast-enhancers ADJ3 lesionary) OR (contrast-enhancers ADJ3 lesionsaccular) OR (contrast-enhancers ADJ3 leasions) OR (contrast-enhances ADJ3 lesions) OR (contrast-enhances ADJ3 lesion) OR (contrast-enhances ADJ3 lesioned) OR (contrast-enhances ADJ3 lesional) OR (contrast-enhances ADJ3 lesionectomy) OR (contrast-enhances ADJ3 lesionography) OR (contrast-enhances ADJ3 lesionsuvmax) OR (contrast-enhances ADJ3 lesionsuvpeak) OR (contrast-enhances ADJ3 lesioning) OR (contrast-enhances ADJ3 lesiones) OR (contrast-enhances ADJ3 lesionary) OR (contrast-enhances ADJ3 lesionsaccular) OR (contrast-enhances ADJ3 leasions) OR (contrast-enhancements ADJ3 lesions) OR (contrast-enhancements ADJ3 lesion) OR (contrast-enhancements ADJ3 lesioned) OR (contrast-enhancements ADJ3 lesional) OR (contrast-enhancements ADJ3 lesionectomy) OR (contrast-enhancements ADJ3 lesionography) OR (contrast-enhancements ADJ3 lesionsuvmax) OR (contrast-enhancements ADJ3 lesionsuvpeak) OR (contrast-enhancements ADJ3 lesioning) OR (contrast-enhancements ADJ3 lesiones) OR (contrast-enhancements ADJ3 lesionary) OR (contrast-enhancements ADJ3 lesionsaccular) OR (contrast-enhancements ADJ3 leasions) OR (contrast-enhance ADJ3 lesions) OR (contrast-enhance ADJ3 lesion) OR (contrast-enhance ADJ3 lesioned) OR (contrast-enhance ADJ3 lesional) OR (contrast-enhance ADJ3 lesionectomy) OR (contrast-enhance ADJ3 lesionography) OR (contrast-enhance ADJ3 lesionsuvmax) OR (contrast-enhance ADJ3 lesionsuvpeak) OR (contrast-enhance ADJ3 lesioning) OR (contrast-enhance ADJ3 lesiones) OR (contrast-enhance ADJ3 lesionary) OR (contrast-enhance ADJ3 lesionsaccular) OR (contrast-enhance ADJ3 leasions) OR (radiation-induced ADJ3 lesions) OR (radiation-induced ADJ3 lesion) OR (radiation-induced ADJ3 lesioned) OR (radiation-induced ADJ3 lesional) OR (radiation-induced ADJ3 lesionectomy) OR (radiation-induced ADJ3 lesionography) OR (radiation-induced ADJ3 lesionsuvmax) OR (radiation-induced ADJ3 lesionsuvpeak) OR (radiation-induced ADJ3 lesioning) OR (radiation-induced ADJ3 lesiones) OR (radiation-induced ADJ3 lesionary) OR (radiation-induced ADJ3 lesionsaccular) OR (radiation-induced ADJ3 leasions) OR (radiation-induces ADJ3 lesions) OR (radiation-induces ADJ3 lesion) OR (radiation-induces ADJ3 lesioned) OR (radiation-induces ADJ3 lesional) OR (radiation-induces ADJ3 lesionectomy) OR (radiation-induces ADJ3 lesionography) OR (radiation-induces ADJ3 lesionsuvmax) OR (radiation-induces ADJ3 lesionsuvpeak) OR (radiation-induces ADJ3 lesioning) OR (radiation-induces ADJ3 lesiones) OR (radiation-induces ADJ3 lesionary) OR (radiation-induces ADJ3 lesionsaccular) OR (radiation-induces ADJ3 leasions) OR (radiation-induce ADJ3 lesions) OR (radiation-induce ADJ3 lesion) OR (radiation-induce ADJ3 lesioned) OR (radiation-induce ADJ3 lesional) OR (radiation-induce ADJ3 lesionectomy) OR (radiation-induce ADJ3 lesionography) OR (radiation-induce ADJ3 lesionsuvmax) OR (radiation-induce ADJ3 lesionsuvpeak) OR (radiation-induce ADJ3 lesioning) OR (radiation-induce ADJ3 lesiones) OR (radiation-induce ADJ3 lesionary) OR (radiation-induce ADJ3 lesionsaccular) OR (radiation-induce ADJ3 leasions) OR (radiation-inducible ADJ3 lesions) OR (radiation-inducible ADJ3 lesion) OR (radiation-inducible ADJ3 lesioned) OR (radiation-inducible ADJ3 lesional) OR (radiation-inducible ADJ3 lesionectomy) OR (radiation-inducible ADJ3 lesionography) OR (radiation-inducible ADJ3 lesionsuvmax) OR (radiation-inducible ADJ3 lesionsuvpeak) OR (radiation-inducible ADJ3 lesioning) OR (radiation-inducible ADJ3 lesiones) OR (radiation-inducible ADJ3 lesionary) OR (radiation-inducible ADJ3 lesionsaccular) OR (radiation-inducible ADJ3 leasions) OR (radiation-induction ADJ3 lesions) OR (radiation-induction ADJ3 lesion) OR (radiation-induction ADJ3 lesioned) OR (radiation-induction ADJ3 lesional) OR (radiation-induction ADJ3 lesionectomy) OR (radiation-induction ADJ3 lesionography) OR (radiation-induction ADJ3 lesionsuvmax) OR (radiation-induction ADJ3 lesionsuvpeak) OR (radiation-induction ADJ3 lesioning) OR (radiation-induction ADJ3 lesiones) OR (radiation-induction ADJ3 lesionary) OR (radiation-induction ADJ3 lesionsaccular) OR (radiation-induction ADJ3 leasions) OR (radiation-inducing ADJ3 lesions) OR (radiation-inducing ADJ3 lesion) OR (radiation-inducing ADJ3 lesioned) OR (radiation-inducing ADJ3 lesional) OR (radiation-inducing ADJ3 lesionectomy) OR (radiation-inducing ADJ3 lesionography) OR (radiation-inducing ADJ3 lesionsuvmax) OR (radiation-inducing ADJ3 lesionsuvpeak) OR (radiation-inducing ADJ3 lesioning) OR (radiation-inducing ADJ3 lesiones) OR (radiation-inducing ADJ3 lesionary) OR (radiation-inducing ADJ3 lesionsaccular) OR (radiation-inducing ADJ3 leasions)))).ti,ab,kf.)

**NOT** ((exp "animal"/ NOT exp "human"/) **OR** ("mouse" **OR** "mice" **OR** "rat" **OR** "rats" **OR** "rodent*" **OR** "pig" **OR** "pigs" **OR** "cattle" **OR** "cow" **OR** "cows" **OR** "horse*" **OR** "monkey*" **OR** "dog" **OR** "dogs" **OR** "cat" **OR** "cats" **OR** "fish" **OR** "mammal*" **OR** "murine*" **OR** "ewe" **OR** "ewes" **OR** "sheep" **OR** "zebrafish").ti) **NOT** (("review" **OR** "preprints" **OR** "conference abstract" **OR** "letter" **OR** "editorials" **OR** "note" **OR** "short survey").pt **OR** ("letter" **OR** "reply" **OR** "response" **OR** "editor*" **OR** "preface" **OR** "opinion" **OR** "meta-analysis" **OR** "meta-synthesis" **OR** "review").ti) **NOT** (exp *brain metastasis/ **OR** "metastas*".ti)

**Terms rejected:**

Cancer: cancerological, cancerogenity, cancerogenus, cancerogenous, cancerology, cancerides, cancerostatic, cancerogens, cancerigenic, cancerization, cancerdata, cancerlit
Neoplasm: Neoplasmleft
Tumor: tumorectomy, tumornecrosis, tumormarker, tuomrlet, tumorlets, tumorsphere, tumorigeneses, tumornecrosisfactor, tumorgenic, tumorgenecity, tumoropathic, tumorogenicity, tumores, tumorigenecity, tumorigens, tumorogenic, tumorsize, tumoricide, tumore, tumorforming, tumorettes
Tumour: tumourettes, tumourbed, tumourigenic, tumourectomy, tumoursuppressor, tumourogenicity, tumourogenesis, tumourology, tumourfluoroscopy
Carcinoma: Carcinomatosa, carcinomata, carcinomatousis, carcinomatosisapropos, carcinomatis
Sarcoma: Sarcomaina, sarcomatoid, sarcomatus, sarcomata

#### Search 2 (van Ruijven et al.) https://pubmed.ncbi.nlm.nih.gov/42293777/

##### Search 2: Original MEDLINE Search: (3110 hits)

(Exp "Computers, Handheld"/ or exp "Digital Health"/ or exp "Mobile applications"/ or exp "Monitoring, ambulatory"/ or exp "Point-of-Care Systems"/ or exp "Rapid Diagnostic Tests"/ or exp "Rapid On-site Evaluation"/ or exp "Telemedicine"/ or "Telemetry"/ or "Videoconferencing"/ or (App or Apps or Desktop or (Digital ADJ3 (care* or consult* or diagnos* or health* or medicine or monitor* or rehab* or therap* or visit*)) or Ehealth* or E-health* or “E-counseling” or “E-Therap*” or (Electronic* ADJ3 (app* or health* or medicine*)) or Emedicine or “E-medicine” or EncephalApp or "Distan* Counsel*" or ((Handheld OR "Hand Held") ADJ3 (computer* or device*)) or “Health app*” or (Home ADJ3 (medicine or monitor*)) or iPad or Laptop or (Medical ADJ3 (app*)) or Mhealth or “M-health*” or (Mobile ADJ3 (app* or computer or device* or health* or medicine* or monitor*)) or "Outpatient Monitor*" or "Patient* Monitor*" or ("Point-of-care" ADJ3 (test* or diagnos*)) or "Rapid Diagnos*" or "Rapid on-site diagnos*" or "Rapid on-site evaluation" or (Remote ADJ3 (care* or consul* or diagnos* or health* or manag* or medicine* or monitor* or rehab*)) or Smartphone* or “smart-phone*” or Smartwatch or Tablet or Telecare* or Teleconsul* or Telediagnos* or Telehealth* or Telemedicine* or Telemonitor* or Telemetr* or “Tele-metr*” or Telerehab* or Telerefer* or Teletherap* or Televisit* or (Tele ADJ3 (care* or consult* or diagnos* or health* or manag* or medicine* or monitor* or rehab* or refer* or therap* or visit*)) or Thealth* or “T health*” or Videoconsult* or “Video Consult*” or Videoconf* or “Video-conf*” or (Virtual ADJ3 (care* or clinic* or consult* or health* or medicine* or monitor* or rehab* or therap* or visit*)) or Watch).ti,ab,kf.) AND (Exp "Hepatic Insufficiency"/ or exp "Hepatopulmonary syndrome"/ or exp "Hepatorenal syndrome"/ or exp "Hypertension, portal"/ or exp "Liver cirrhosis"/ or exp Ascites/ or (Ascites or Cirrh* or ((Chronic*) ADJ3 (liver OR hepatic*)) or CLD or Encephalopath* or (("end stage" or "late stage") ADJ3 (liver* or hepatic*)) or ESLD or (esophag* ADJ3 (varix* or varic*)) or (oesophag* ADJ3 (varix* or varic*)) or (gastric* ADJ3 (varix* or varic*)) or ((liver or hepatic*) ADJ3 (fibrosis or fail* or insufficien* or dysfunction*)) or "hepatopulmonary syndrome*" or "hepato-pulmonary syndrome*" or "hepatorenal syndrome*" or "hepato-renal syndrome*" or "portal hypertension" or (varic* ADJ3 (hemorrhag* or bleed*)) or (varix* ADJ3 (hemorrhag* or bleed*))).ti,ab,kf.)

##### Search 2: REMI 2 Proximity Search convert to Medline (2790 hits):

(Exp "Computers, Handheld"/ or exp "Digital Health"/ or exp "Mobile applications"/ or exp "Monitoring, ambulatory"/ or exp "Point-of-Care Systems"/ or exp "Rapid Diagnostic Tests"/ or exp "Rapid On-site Evaluation"/ or exp "Telemedicine"/ or "Telemetry"/ or "Videoconferencing"/ or (App or Apps or Desktop or ((digital ADJ3 care) OR (digital ADJ3 caregivers) OR (digital ADJ3 caregiver) OR (digital ADJ3 carestream) OR (digital ADJ3 carers) OR (digital ADJ3 career) OR (digital ADJ3 careful) OR (digital ADJ3 caregiving) OR (digital ADJ3 careggi) OR (digital ADJ3 careless) OR (digital ADJ3 carefully) OR (digital ADJ3 carer) OR (digital ADJ3 careers) OR (digital ADJ3 cared) OR (digital ADJ3 caretta) OR (digital ADJ3 carey) OR (digital ADJ3 carebara) OR (digital ADJ3 cares) OR (digital ADJ3 carefulness) OR (digital ADJ3 caressing) OR (digital ADJ3 caretakers) OR (digital ADJ3 carettacaretta) OR (digital ADJ3 consultation) OR (digital ADJ3 consultations) OR (digital ADJ3 consulting) OR (digital ADJ3 consulted) OR (digital ADJ3 consultants) OR (digital ADJ3 consultancy) OR (digital ADJ3 consultant) OR (digital ADJ3 consult) OR (digital ADJ3 consultative) OR (digital ADJ3 consultando) OR (digital ADJ3 consults) OR (digital ADJ3 diagnostic) OR (digital ADJ3 diagnosis) OR (digital ADJ3 diagnostics) OR (digital ADJ3 diagnosed) OR (digital ADJ3 diagnoses) OR (digital ADJ3 diagnosing) OR (digital ADJ3 diagnosticity) OR (digital ADJ3 diagnosable) OR (digital ADJ3 diagnostically) OR (digital ADJ3 diagnose) OR (digital ADJ3 diagnostical) OR (digital ADJ3 health) OR (digital ADJ3 healthcare) OR (digital ADJ3 healthy) OR (digital ADJ3 healthier) OR (digital ADJ3 healthineers) OR (digital ADJ3 healthymoms4mentalhealth) OR (digital ADJ3 healthkit) OR (digital ADJ3 healthism) OR (digital ADJ3 health4life) OR (digital ADJ3 healthcommunication) OR (digital ADJ3 healthily) OR (digital ADJ3 healthgrade) OR (digital ADJ3 medicine) OR (digital ADJ3 monitoring) OR (digital ADJ3 monitor) OR (digital ADJ3 monitored) OR (digital ADJ3 monitors) OR (digital ADJ3 monitorable) OR (digital ADJ3 monitorization) OR (digital ADJ3 rehabilitation) OR (digital ADJ3 rehabilitations) OR (digital ADJ3 rehabilitating) OR (digital ADJ3 rehabilitative) OR (digital ADJ3 rehabilitated) OR (digital ADJ3 rehabilitate) OR (digital ADJ3 rehab) OR (digital ADJ3 therapy) OR (digital ADJ3 therapeutic) OR (digital ADJ3 therapies) OR (digital ADJ3 therapeutics) OR (digital ADJ3 therapist) OR (digital ADJ3 therapists) OR (digital ADJ3 therapeutically) OR (digital ADJ3 therapsida) OR (digital ADJ3 theraphosidae) OR (digital ADJ3 theraphosid) OR (digital ADJ3 therapsid) OR (digital ADJ3 therapeutical) OR (digital ADJ3 visits) OR (digital ADJ3 visited) OR (digital ADJ3 visit) OR (digital ADJ3 visitation) OR (digital ADJ3 visiting) OR (digital ADJ3 visitor) OR (digital ADJ3 visitors)) or Ehealth* or E-health* or E-counseling or E-Therap* or ((electronic ADJ3 app) OR (electronic ADJ3 apps) OR (electronic ADJ3 applications) OR (electronic ADJ3 application) OR (electronic ADJ3 appliance) OR (electronic ADJ3 applicators) OR (electronic ADJ3 applicative) OR (electronic ADJ3 applicational) OR (electronic ADJ3 applet) OR (electronic ADJ3 applicationswe) OR (electronic ADJ3 applicaitions) OR (electronic ADJ3 applicationspeople) OR (electronic ADJ3 applcations) OR (electronic ADJ3 applications1) OR (electronic ADJ3 appliction) OR (electronic ADJ3 applicationof) OR (electronic ADJ3 health) OR (electronic ADJ3 healthcare) OR (electronic ADJ3 healthy) OR (electronic ADJ3 healthstar) OR (electronic ADJ3 healthful) OR (electronic ADJ3 healthsystem) OR (electronic ADJ3 healthier) OR (electronic ADJ3 healthiest) OR (electronic ADJ3 healthiness) OR (electronic ADJ3 healthlink) OR (electronic ADJ3 healthspan) OR (electronic ADJ3 healthsource) OR (electronic ADJ3 healthemove) OR (electronic ADJ3 healthinfonet) OR (electronic ADJ3 healthlnk) OR (electronic ADJ3 healthfully) OR (electronic ADJ3 healthcheck) OR (electronic ADJ3 healthwatch) OR (electronic ADJ3 healthfulness) OR (electronic ADJ3 healthrelated) OR (electronic ADJ3 healths) OR (electronic ADJ3 healthstyle) OR (electronic ADJ3 healthsystems) OR (electronic ADJ3 healthconnect) OR (electronic ADJ3 healthmatics) OR (electronic ADJ3 healthly) OR (electronic ADJ3 medicine) OR (electronic ADJ3 medicines) OR (electronic ADJ3 medicineinsight) OR (electronic ADJ3 medicineonline) OR (electronic ADJ3 medicinewise) OR (electronical ADJ3 app) OR (electronical ADJ3 apps) OR (electronical ADJ3 applications) OR (electronical ADJ3 application) OR (electronical ADJ3 appliance) OR (electronical ADJ3 applicators) OR (electronical ADJ3 applicative) OR (electronical ADJ3 applicational) OR (electronical ADJ3 applet) OR (electronical ADJ3 applicationswe) OR (electronical ADJ3 applicaitions) OR (electronical ADJ3 applicationspeople) OR (electronical ADJ3 applcations) OR (electronical ADJ3 applications1) OR (electronical ADJ3 appliction) OR (electronical ADJ3 applicationof) OR (electronical ADJ3 health) OR (electronical ADJ3 healthcare) OR (electronical ADJ3 healthy) OR (electronical ADJ3 healthstar) OR (electronical ADJ3 healthful) OR (electronical ADJ3 healthsystem) OR (electronical ADJ3 healthier) OR (electronical ADJ3 healthiest) OR (electronical ADJ3 healthiness) OR (electronical ADJ3 healthlink) OR (electronical ADJ3 healthspan) OR (electronical ADJ3 healthsource) OR (electronical ADJ3 healthemove) OR (electronical ADJ3 healthinfonet) OR (electronical ADJ3 healthlnk) OR (electronical ADJ3 healthfully) OR (electronical ADJ3 healthcheck) OR (electronical ADJ3 healthwatch) OR (electronical ADJ3 healthfulness) OR (electronical ADJ3 healthrelated) OR (electronical ADJ3 healths) OR (electronical ADJ3 healthstyle) OR (electronical ADJ3 healthsystems) OR (electronical ADJ3 healthconnect) OR (electronical ADJ3 healthmatics) OR (electronical ADJ3 healthly) OR (electronical ADJ3 medicine) OR (electronical ADJ3 medicines) OR (electronical ADJ3 medicineinsight) OR (electronical ADJ3 medicineonline) OR (electronical ADJ3 medicinewise) OR (electronics ADJ3 app) OR (electronics ADJ3 apps) OR (electronics ADJ3 applications) OR (electronics ADJ3 application) OR (electronics ADJ3 appliance) OR (electronics ADJ3 applicators) OR (electronics ADJ3 applicative) OR (electronics ADJ3 applicational) OR (electronics ADJ3 applet) OR (electronics ADJ3 applicationswe) OR (electronics ADJ3 applicaitions) OR (electronics ADJ3 applicationspeople) OR (electronics ADJ3 applcations) OR (electronics ADJ3 applications1) OR (electronics ADJ3 appliction) OR (electronics ADJ3 applicationof) OR (electronics ADJ3 health) OR (electronics ADJ3 healthcare) OR (electronics ADJ3 healthy) OR (electronics ADJ3 healthstar) OR (electronics ADJ3 healthful) OR (electronics ADJ3 healthsystem) OR (electronics ADJ3 healthier) OR (electronics ADJ3 healthiest) OR (electronics ADJ3 healthiness) OR (electronics ADJ3 healthlink) OR (electronics ADJ3 healthspan) OR (electronics ADJ3 healthsource) OR (electronics ADJ3 healthemove) OR (electronics ADJ3 healthinfonet) OR (electronics ADJ3 healthlnk) OR (electronics ADJ3 healthfully) OR (electronics ADJ3 healthcheck) OR (electronics ADJ3 healthwatch) OR (electronics ADJ3 healthfulness) OR (electronics ADJ3 healthrelated) OR (electronics ADJ3 healths) OR (electronics ADJ3 healthstyle) OR (electronics ADJ3 healthsystems) OR (electronics ADJ3 healthconnect) OR (electronics ADJ3 healthmatics) OR (electronics ADJ3 healthly) OR (electronics ADJ3 medicine) OR (electronics ADJ3 medicines) OR (electronics ADJ3 medicineinsight) OR (electronics ADJ3 medicineonline) OR (electronics ADJ3 medicinewise) OR (electronically ADJ3 app) OR (electronically ADJ3 apps) OR (electronically ADJ3 applications) OR (electronically ADJ3 application) OR (electronically ADJ3 appliance) OR (electronically ADJ3 applicators) OR (electronically ADJ3 applicative) OR (electronically ADJ3 applicational) OR (electronically ADJ3 applet) OR (electronically ADJ3 applicationswe) OR (electronically ADJ3 applicaitions) OR (electronically ADJ3 applicationspeople) OR (electronically ADJ3 applcations) OR (electronically ADJ3 applications1) OR (electronically ADJ3 appliction) OR (electronically ADJ3 applicationof) OR (electronically ADJ3 health) OR (electronically ADJ3 healthcare) OR (electronically ADJ3 healthy) OR (electronically ADJ3 healthstar) OR (electronically ADJ3 healthful) OR (electronically ADJ3 healthsystem) OR (electronically ADJ3 healthier) OR (electronically ADJ3 healthiest) OR (electronically ADJ3 healthiness) OR (electronically ADJ3 healthlink) OR (electronically ADJ3 healthspan) OR (electronically ADJ3 healthsource) OR (electronically ADJ3 healthemove) OR (electronically ADJ3 healthinfonet) OR (electronically ADJ3 healthlnk) OR (electronically ADJ3 healthfully) OR (electronically ADJ3 healthcheck) OR (electronically ADJ3 healthwatch) OR (electronically ADJ3 healthfulness) OR (electronically ADJ3 healthrelated) OR (electronically ADJ3 healths) OR (electronically ADJ3 healthstyle) OR (electronically ADJ3 healthsystems) OR (electronically ADJ3 healthconnect) OR (electronically ADJ3 healthmatics) OR (electronically ADJ3 healthly) OR (electronically ADJ3 medicine) OR (electronically ADJ3 medicines) OR (electronically ADJ3 medicineinsight) OR (electronically ADJ3 medicineonline) OR (electronically ADJ3 medicinewise) OR (electronica ADJ3 app) OR (electronica ADJ3 apps) OR (electronica ADJ3 applications) OR (electronica ADJ3 application) OR (electronica ADJ3 appliance) OR (electronica ADJ3 applicators) OR (electronica ADJ3 applicative) OR (electronica ADJ3 applicational) OR (electronica ADJ3 applet) OR (electronica ADJ3 applicationswe) OR (electronica ADJ3 applicaitions) OR (electronica ADJ3 applicationspeople) OR (electronica ADJ3 applcations) OR (electronica ADJ3 applications1) OR (electronica ADJ3 appliction) OR (electronica ADJ3 applicationof) OR (electronica ADJ3 health) OR (electronica ADJ3 healthcare) OR (electronica ADJ3 healthy) OR (electronica ADJ3 healthstar) OR (electronica ADJ3 healthful) OR (electronica ADJ3 healthsystem) OR (electronica ADJ3 healthier) OR (electronica ADJ3 healthiest) OR (electronica ADJ3 healthiness) OR (electronica ADJ3 healthlink) OR (electronica ADJ3 healthspan) OR (electronica ADJ3 healthsource) OR (electronica ADJ3 healthemove) OR (electronica ADJ3 healthinfonet) OR (electronica ADJ3 healthlnk) OR (electronica ADJ3 healthfully) OR (electronica ADJ3 healthcheck) OR (electronica ADJ3 healthwatch) OR (electronica ADJ3 healthfulness) OR (electronica ADJ3 healthrelated) OR (electronica ADJ3 healths) OR (electronica ADJ3 healthstyle) OR (electronica ADJ3 healthsystems) OR (electronica ADJ3 healthconnect) OR (electronica ADJ3 healthmatics) OR (electronica ADJ3 healthly) OR (electronica ADJ3 medicine) OR (electronica ADJ3 medicines) OR (electronica ADJ3 medicineinsight) OR (electronica ADJ3 medicineonline) OR (electronica ADJ3 medicinewise)) or Emedicine or “E-medicine” or EncephalApp or "Distan* Counsel*" or ((handheld ADJ3 computer) OR (handheld ADJ3 computerized) OR (handheld ADJ3 computers) OR (handheld ADJ3 computerised) OR (handheld ADJ3 device) OR (handheld ADJ3 devices) OR (hand-held ADJ3 computer) OR (hand-held ADJ3 computerized) OR (hand-held ADJ3 computers) OR (hand-held ADJ3 computerised) OR (hand-held ADJ3 device) OR (hand-held ADJ3 devices)) or “Health app*” or ((home ADJ3 medicine) OR (home ADJ3 monitoring) OR (home ADJ3 monitor) OR (home ADJ3 monitored) OR (home ADJ3 monitors) OR (home ADJ3 monitorate) OR (home ADJ3 monitorable) OR (home ADJ3 monitory))
or iPad or Laptop or ((medical ADJ3 app) OR (medical ADJ3 apps) OR (medical ADJ3 applied) OR (medical ADJ3 application) OR (medical ADJ3 applications) OR (medical ADJ3 appliance) OR (medical ADJ3 appliances) OR (medical ADJ3 applicator) OR (medical ADJ3 applicability) OR (medical ADJ3 applicable) OR (medical ADJ3 applying) OR (medical ADJ3 apply) OR (medical ADJ3 applicants) OR (medical ADJ3 applicators) OR (medical ADJ3 applet) OR (medical ADJ3 applies) OR (medical ADJ3 applicant) OR (medical ADJ3 apples) OR (medical ADJ3 apple) OR (medical ADJ3 applesauce) OR (medical ADJ3 applanatum) OR (medical ADJ3 applanation) OR (medical ADJ3 applebaum) OR (medical ADJ3 applicationthe) OR (medical ADJ3 applaud) OR (medical ADJ3 appl) OR (medical ADJ3 applicative) OR (medical ADJ3 appleton) OR (medical ADJ3 apple1986) OR (medical ADJ3 appleby) OR (medical ADJ3 applicationthese) OR (medical ADJ3 appliedness) OR (medical ADJ3 applique) OR (medical ADJ3 applicationsin) OR (medical ADJ3 applier) OR (medical ADJ3 appliquee) OR (medical ADJ3 applicationswe) OR (medical ADJ3 applica) OR (medical ADJ3 applauded) OR (medical ADJ3 applicationsimple) OR (medical ADJ3 applciations) OR (medical ADJ3 appl1) OR (medical ADJ3 applicated) OR (medical ADJ3 applauds) OR (medical ADJ3 applause)) or Mhealth or “M-health*” or ((mobile ADJ3 app) OR (mobile ADJ3 apps) OR (mobile ADJ3 application) OR (mobile ADJ3 applications) OR (mobile ADJ3 applied) OR (mobile ADJ3 applicability) OR (mobile ADJ3 applicator) OR (mobile ADJ3 applicable) OR (mobile ADJ3 applicators) OR (mobile ADJ3 applying) OR (mobile ADJ3 apply) OR (mobile ADJ3 appliance) OR (mobile ADJ3 apple) OR (mobile ADJ3 applies) OR (mobile ADJ3 applicants) OR (mobile ADJ3 apples) OR (mobile ADJ3 appltree) OR (mobile ADJ3 applet) OR (mobile ADJ3 applicationstate) OR (mobile ADJ3 appling) OR (mobile ADJ3 applicant) OR (mobile ADJ3 appliances) OR (mobile ADJ3 applicationsresults) OR (mobile ADJ3 applicated) OR (mobile ADJ3 computer) OR (mobile ADJ3 devices) OR (mobile ADJ3 device) OR (mobile ADJ3 health) OR (mobile ADJ3 healthcare) OR (mobile ADJ3 healthy) OR (mobile ADJ3 healthier) OR (mobile ADJ3 healthineers) OR (mobile ADJ3 healthmindr) OR (mobile ADJ3 healthemove) OR (mobile ADJ3 healthy365) OR (mobile ADJ3 healthrelated) OR (mobile ADJ3 healthspan) OR (mobile ADJ3 healthily) OR (mobile ADJ3 medicine) OR (mobile ADJ3 medicines) OR (mobile ADJ3 monitoring) OR (mobile ADJ3 monitor) OR (mobile ADJ3 monitored) OR (mobile ADJ3 monitors) OR (mobile ADJ3 monitorable) OR (mobile ADJ3 monitorization)) or "Outpatient Monitor*" or "Patient* Monitor*" or ((point-of-care ADJ3 testing) OR (point-of-care ADJ3 test) OR (point-of-care ADJ3 tests) OR (point-of-care ADJ3 tested) OR (point-of-care ADJ3 testicular) OR (point-of-care ADJ3 testable) OR (point-of-care ADJ3 testudo) OR (point-of-care ADJ3 testbed) OR (point-of-care ADJ3 testers) OR (point-of-care ADJ3 testis) OR (point-of-care ADJ3 testicle) OR (point-of-care ADJ3 testkit) OR (point-of-care ADJ3 testosterone) OR (point-of-care ADJ3 teste) OR (point-of-care ADJ3 testament) OR (point-of-care ADJ3 diagnostic) OR (point-of-care ADJ3 diagnosis) OR (point-of-care ADJ3 diagnostics) OR (point-of-care ADJ3 diagnosed) OR (point-of-care ADJ3 diagnosing) OR (point-of-care ADJ3 diagnoses) OR (point-of-care ADJ3 diagnose) OR (point-of-care ADJ3 diagnostically) OR (point-of-care ADJ3 diagnosability) OR (point-of-care ADJ3 diagnosticpathology)) or "Rapid Diagnos*" or "Rapid on-site diagnos*" or "Rapid on-site evaluation" or ((remote ADJ3 care) OR (remote ADJ3 caregiver) OR (remote ADJ3 caregivers) OR (remote ADJ3 carers) OR (remote ADJ3 careers) OR (remote ADJ3 carex) OR (remote ADJ3 careful) OR (remote ADJ3 carer) OR (remote ADJ3 career) OR (remote ADJ3 caretakers) OR (remote ADJ3 caretaker) OR (remote ADJ3 caregiving) OR (remote ADJ3 carefully) OR (remote ADJ3 caretta) OR (remote ADJ3 caress) OR (remote ADJ3 carehpv) OR (remote ADJ3 cared) OR (remote ADJ3 cares) OR (remote ADJ3 carepath) OR (remote ADJ3 carettacaretta) OR (remote ADJ3 caresses) OR (remote ADJ3 carexane) OR (remote ADJ3 carea) OR (remote ADJ3 caret) OR (remote ADJ3 carefor) OR (remote ADJ3 carestiae) OR (remote ADJ3 carelessness) OR (remote ADJ3 carew) OR (remote ADJ3 caretaking) OR (remote ADJ3 consultations) OR (remote ADJ3 consultation) OR (remote ADJ3 consulting) OR (remote ADJ3 consults) OR (remote ADJ3 consult) OR (remote ADJ3 consultands) OR (remote ADJ3 consultancy) OR (remote ADJ3 consultant) OR (remote ADJ3 consulted) OR (remote ADJ3 consultants) OR (remote ADJ3 consultancies) OR (remote ADJ3 consultative) OR (remote ADJ3 diagnostic) OR (remote ADJ3 diagnosis) OR (remote ADJ3 diagnostics) OR (remote ADJ3 diagnoses) OR (remote ADJ3 diagnosed) OR (remote ADJ3 diagnostically) OR (remote ADJ3 diagnosa) OR (remote ADJ3 diagnose) OR (remote ADJ3 diagnosing) OR (remote ADJ3 diagnosticpathology) OR (remote ADJ3 diagnosticums) OR (remote ADJ3 diagnostis) OR (remote ADJ3 health) OR (remote ADJ3 healthcare) OR (remote ADJ3 healthy) OR (remote ADJ3 healthier) OR (remote ADJ3 healthiest) OR (remote ADJ3 healthsystem) OR (remote ADJ3 healthspan) OR (remote ADJ3 healthliners) OR (remote ADJ3 healthfulness) OR (remote ADJ3 healthiness) OR (remote ADJ3 healthful) OR (remote ADJ3 healthstar) OR (remote ADJ3 management) OR (remote ADJ3 managed) OR (remote ADJ3 managing) OR (remote ADJ3 manageable) OR (remote ADJ3 manage) OR (remote ADJ3 managers) OR (remote ADJ3 managerial) OR (remote ADJ3 manag) OR (remote ADJ3 manages) OR (remote ADJ3 managements) OR (remote ADJ3 manager) OR (remote ADJ3 manager5) OR (remote ADJ3 managerialism) OR (remote ADJ3 medicine) OR (remote ADJ3 medicines) OR (remote ADJ3 medicineat) OR (remote ADJ3 monitoring) OR (remote ADJ3 monitored) OR (remote ADJ3 monitor) OR (remote ADJ3 monitors) OR (remote ADJ3 monitorization) OR (remote ADJ3 monitoringmethods) OR (remote ADJ3 rehabilitation) OR (remote ADJ3 rehabilitated) OR (remote ADJ3 rehabilitate) OR (remote ADJ3 rehabilitations) OR (remote ADJ3 rehabilitating) OR (remote ADJ3 rehabilitative) OR (remote ADJ3 rehab) OR (remote ADJ3 rehabilitationsmartphone) OR (remote ADJ3 rehabilitationwe) OR (remote ADJ3 rehabilitationthe) OR (remote ADJ3 rehabilitationremote)) or Smartphone* or “smart-phone*” or Smartwatch or Tablet or Telecare* or Teleconsul* or Telediagnos* or Telehealth* or Telemedicine* or Telemonitor* or Telemetr* or “Tele-metr*” or Telerehab* or Telerefer* or Teletherap* or Televisit* or ((tele ADJ3 care) OR (tele ADJ3 caregivers) OR (tele ADJ3 caregiver) OR (tele ADJ3 caregiving) OR (tele ADJ3 career) OR (tele ADJ3 careline) OR (tele ADJ3 careful) OR (tele ADJ3 careers) OR (tele ADJ3 carers) OR (tele ADJ3 carefully) OR (tele ADJ3 consultation) OR (tele ADJ3 consultations) OR (tele ADJ3 consults) OR (tele ADJ3 consultants) OR (tele ADJ3 consulting) OR (tele ADJ3 consult) OR (tele ADJ3 consulted) OR (tele ADJ3 consultant) OR (tele ADJ3 consultative) OR (tele ADJ3 consulta) OR (tele ADJ3 consultancy) OR (tele ADJ3 diagnostic) OR (tele ADJ3 diagnosis) OR (tele ADJ3 diagnostics) OR (tele ADJ3 diagnoses) OR (tele ADJ3 diagnosed) OR (tele ADJ3 diagnosing) OR (tele ADJ3 diagnostically) OR (tele ADJ3 diagnose) OR (tele ADJ3 diagnostician) OR (tele ADJ3 diagnosticians) OR (tele ADJ3 health) OR (tele ADJ3 healthcare) OR (tele ADJ3 healthy) OR (tele ADJ3 healthier) OR (tele ADJ3 healthspan) OR (tele ADJ3 management) OR (tele ADJ3 managing) OR (tele ADJ3 manage) OR (tele ADJ3 managed) OR (tele ADJ3 managers) OR (tele ADJ3 manager) OR (tele ADJ3 managerial) OR (tele ADJ3 manageable) OR (tele ADJ3 medicine) OR (tele ADJ3 medicines) OR (tele ADJ3 monitoring) OR (tele ADJ3 monitored) OR (tele ADJ3 monitor) OR (tele ADJ3 monitors) OR (tele ADJ3 rehabilitation) OR (tele ADJ3 rehab) OR (tele ADJ3 rehabi) OR (tele ADJ3 rehabilitative) OR (tele ADJ3 rehabilomics) OR (tele ADJ3 referral) OR (tele ADJ3 referrals) OR (tele ADJ3 reference) OR (tele ADJ3 referred) OR (tele ADJ3 referring) OR (tele ADJ3 references) OR (tele ADJ3 refers) OR (tele ADJ3 referenced) OR (tele ADJ3 refer) OR (tele ADJ3 referential) OR (tele ADJ3 referencing) OR (tele ADJ3 referencia) OR (tele ADJ3 therapy) OR (tele ADJ3 therapeutic) OR (tele ADJ3 therapists) OR (tele ADJ3 therapies) OR (tele ADJ3 therapist) OR (tele ADJ3 therapeutics) OR (tele ADJ3 visits) OR (tele ADJ3 visit) OR (tele ADJ3 visitor) OR (tele ADJ3 visiting) OR (tele ADJ3 visited) OR (tele ADJ3 visitors)) or Thealth* or “T health*” or Videoconsult* or “Video Consult*” or Videoconf* or “Video-conf*” or ((virtual ADJ3 care) OR (virtual ADJ3 caregivers) OR (virtual ADJ3 careers) OR (virtual ADJ3 caregiver) OR (virtual ADJ3 career) OR (virtual ADJ3 carestream) OR (virtual ADJ3 careful) OR (virtual ADJ3 caregiving) OR (virtual ADJ3 cares) OR (virtual ADJ3 carentonensis) OR (virtual ADJ3 carex) OR (virtual ADJ3 caresses) OR (virtual ADJ3 carers) OR (virtual ADJ3 caretakers) OR (virtual ADJ3 carefully) OR (virtual ADJ3 caregivr) OR (virtual ADJ3 carey) OR (virtual ADJ3 caretta) OR (virtual ADJ3 carene) OR (virtual ADJ3 cared) OR (virtual ADJ3 carer) OR (virtual ADJ3 careless) OR (virtual ADJ3 carest) OR (virtual ADJ3 careertrack) OR (virtual ADJ3 caressed) OR (virtual ADJ3 carefree) OR (virtual ADJ3 caren) OR (virtual ADJ3 careaga) OR (virtual ADJ3 caret) OR (virtual ADJ3 caress) OR (virtual ADJ3 carenabronchial) OR (virtual ADJ3 clinical) OR (virtual ADJ3 clinically) OR (virtual ADJ3 clinic) OR (virtual ADJ3 clinicians) OR (virtual ADJ3 clinician) OR (virtual ADJ3 clinicaltrials) OR (virtual ADJ3 clinicopathologic) OR (virtual ADJ3 clinics) OR (virtual ADJ3 clinicavr) OR (virtual ADJ3 clinicopathological) OR (virtual ADJ3 clinicals) OR (virtual ADJ3 clinicopathologically) OR (virtual ADJ3 clinico) OR (virtual ADJ3 clinicaltrial) OR (virtual ADJ3 clinicoradiological) OR (virtual ADJ3 clinicalroms) OR (virtual ADJ3 clinicalct) OR (virtual ADJ3 clinicalpractice) OR (virtual ADJ3 clinicas) OR (virtual ADJ3 consultations) OR (virtual ADJ3 consultation) OR (virtual ADJ3 consultants) OR (virtual ADJ3 consultancy) OR (virtual ADJ3 consult) OR (virtual ADJ3 consultant) OR (virtual ADJ3 consults) OR (virtual ADJ3 consultative) OR (virtual ADJ3 consulting) OR (virtual ADJ3 consulted) OR (virtual ADJ3 health) OR (virtual ADJ3 healthcare) OR (virtual ADJ3 healthy) OR (virtual ADJ3 healthineers) OR (virtual ADJ3 healthier) OR (virtual ADJ3 healthmpowerment) OR (virtual ADJ3 healthspan) OR (virtual ADJ3 healthiness) OR (virtual ADJ3 healthcomm) OR (virtual ADJ3 healthineer) OR (virtual ADJ3 healthgrid) OR (virtual ADJ3 medicine) OR (virtual ADJ3 medicines) OR (virtual ADJ3 monitoring) OR (virtual ADJ3 monitor) OR (virtual ADJ3 monitored) OR (virtual ADJ3 monitors) OR (virtual ADJ3 monitorizacion) OR (virtual ADJ3 rehabilitation) OR (virtual ADJ3 rehabilitating) OR (virtual ADJ3 rehabilitated) OR (virtual ADJ3 rehabilitative) OR (virtual ADJ3 rehabilitations) OR (virtual ADJ3 rehabilitate) OR (virtual ADJ3 rehabilitationxr) OR (virtual ADJ3 rehabi) OR (virtual ADJ3 rehabdata) OR (virtual ADJ3 rehabilitation3d) OR (virtual ADJ3 rehabilitationpeople) OR (virtual ADJ3 rehabilitationthis) OR (virtual ADJ3 rehabilitationhaving) OR (virtual ADJ3 rehabilitationassessment) OR (virtual ADJ3 rehabilitationthere) OR (virtual ADJ3 therapy) OR (virtual ADJ3 therapeutic) OR (virtual ADJ3 therapeutics) OR (virtual ADJ3 therapies) OR (virtual ADJ3 therapists) OR (virtual ADJ3 therapeutical) OR (virtual ADJ3 therapist) OR (virtual ADJ3 therapeutically) OR (virtual ADJ3 therapsida) OR (virtual ADJ3 therapsid) OR (virtual ADJ3 therapsids) OR (virtual ADJ3 therapeutictarget) OR (virtual ADJ3 therapeuticals) OR (virtual ADJ3 therapeuticapplications) OR (virtual ADJ3 visits) OR (virtual ADJ3 visited) OR (virtual ADJ3 visit) OR (virtual ADJ3 visitors) OR (virtual ADJ3 visitation) OR (virtual ADJ3 visiting) OR (virtual ADJ3 visitor)) or Watch).ti,ab,kf.) AND (Exp "Hepatic Insufficiency"/ or exp "Hepatopulmonary syndrome"/ or exp "Hepatorenal syndrome"/ or exp "Hypertension, portal"/ or exp "Liver cirrhosis"/ or exp Ascites/ or (Ascites or Cirrh* or ((chronic ADJ3 liver) OR (chronic ADJ3 hepatic) OR (chronic ADJ3 hepatica) OR (chronic ADJ3 hepaticojejunostomy) OR (chronic ADJ3 hepatically) OR (chronic ADJ3 hepatical) OR (chronical ADJ3 liver) OR (chronical ADJ3 hepatic) OR (chronical ADJ3 hepatica) OR (chronical ADJ3 hepaticojejunostomy) OR (chronical ADJ3 hepatically) OR (chronical ADJ3 hepatical) OR (chronicle ADJ3 liver) OR (chronicle ADJ3 hepatic) OR (chronicle ADJ3 hepatica) OR (chronicle ADJ3 hepaticojejunostomy) OR (chronicle ADJ3 hepatically) OR (chronicle ADJ3 hepatical) OR (chronica ADJ3 liver) OR (chronica ADJ3 hepatic) OR (chronica ADJ3 hepatica) OR (chronica ADJ3 hepaticojejunostomy) OR (chronica ADJ3 hepatically) OR (chronica ADJ3 hepatical) OR (chronically ADJ3 liver) OR (chronically ADJ3 hepatic) OR (chronically ADJ3 hepatica) OR (chronically ADJ3 hepaticojejunostomy) OR (chronically ADJ3 hepatically) OR (chronically ADJ3 hepatical) OR (chronicity ADJ3 liver) OR (chronicity ADJ3 hepatic) OR (chronicity ADJ3 hepatica) OR (chronicity ADJ3 hepaticojejunostomy) OR (chronicity ADJ3 hepatically) OR (chronicity ADJ3 hepatical) OR (chronicled ADJ3 liver) OR (chronicled ADJ3 hepatic) OR (chronicled ADJ3 hepatica) OR (chronicled ADJ3 hepaticojejunostomy) OR (chronicled ADJ3 hepatically) OR (chronicled ADJ3 hepatical) OR (chronicization ADJ3 liver) OR (chronicization ADJ3 hepatic) OR (chronicization ADJ3 hepatica) OR (chronicization ADJ3 hepaticojejunostomy) OR (chronicization ADJ3 hepatically) OR (chronicization ADJ3 hepatical) OR (chronicliver ADJ3 liver) OR (chronicliver ADJ3 hepatic) OR (chronicliver ADJ3 hepatica) OR (chronicliver ADJ3 hepaticojejunostomy) OR (chronicliver ADJ3 hepatically) OR (chronicliver ADJ3 hepatical) OR (chronicisation ADJ3 liver) OR (chronicisation ADJ3 hepatic) OR (chronicisation ADJ3 hepatica) OR (chronicisation ADJ3 hepaticojejunostomy) OR (chronicisation ADJ3 hepatically) OR (chronicisation ADJ3 hepatical) OR (chronicex ADJ3 liver) OR (chronicex ADJ3 hepatic) OR (chronicex ADJ3 hepatica) OR (chronicex ADJ3 hepaticojejunostomy) OR (chronicex ADJ3 hepatically) OR (chronicex ADJ3 hepatical) OR (chronicin ADJ3 liver) OR (chronicin ADJ3 hepatic) OR (chronicin ADJ3 hepatica) OR (chronicin ADJ3 hepaticojejunostomy) OR (chronicin ADJ3 hepatically) OR (chronicin ADJ3 hepatical) OR (chronicles ADJ3 liver) OR (chronicles ADJ3 hepatic) OR (chronicles ADJ3 hepatica) OR (chronicles ADJ3 hepaticojejunostomy) OR (chronicles ADJ3 hepatically) OR (chronicles ADJ3 hepatical) OR (chronicinflammation ADJ3 liver) OR (chronicinflammation ADJ3 hepatic) OR (chronicinflammation ADJ3 hepatica) OR (chronicinflammation ADJ3 hepaticojejunostomy) OR (chronicinflammation ADJ3 hepatically) OR (chronicinflammation ADJ3 hepatical) OR (chronicliverdiseases ADJ3 liver) OR (chronicliverdiseases ADJ3 hepatic) OR (chronicliverdiseases ADJ3 hepatica) OR (chronicliverdiseases ADJ3 hepaticojejunostomy) OR (chronicliverdiseases ADJ3 hepatically) OR (chronicliverdiseases ADJ3 hepatical) OR (chronicallyinfected ADJ3 liver) OR (chronicallyinfected ADJ3 hepatic) OR (chronicallyinfected ADJ3 hepatica) OR (chronicallyinfected ADJ3 hepaticojejunostomy) OR (chronicallyinfected ADJ3 hepatically) OR (chronicallyinfected ADJ3 hepatical) OR (chronice ADJ3 liver) OR (chronice ADJ3 hepatic) OR (chronice ADJ3 hepatica) OR (chronice ADJ3 hepaticojejunostomy) OR (chronice ADJ3 hepatically) OR (chronice ADJ3 hepatical) OR (chronichepatitis ADJ3 liver) OR (chronichepatitis ADJ3 hepatic) OR (chronichepatitis ADJ3 hepatica) OR (chronichepatitis ADJ3 hepaticojejunostomy) OR (chronichepatitis ADJ3 hepatically) OR (chronichepatitis ADJ3 hepatical)) or CLD or Encephalopath* or ((end ADJ3 stage ADJ3 liver) OR (end ADJ3 stage ADJ3 livers) OR (end ADJ3 stage ADJ3 liverpool) OR (end ADJ3 stage ADJ3 liverwort) OR (end ADJ3 stage ADJ3 liverworts) OR (end ADJ3 stage ADJ3 livera) OR (end ADJ3 stage ADJ3 livermultiscan) OR (end ADJ3 stage ADJ3 hepatic) OR (end ADJ3 stage ADJ3 hepatica) OR (end ADJ3 stage ADJ3 hepaticojejunostomy) OR (end ADJ3 stage ADJ3 hepatically) OR (end ADJ3 stage ADJ3 hepaticus) OR (end ADJ3 stage ADJ3 hepaticae) OR (late ADJ3 stage ADJ3 liver) OR (late ADJ3 stage ADJ3 livers) OR (late ADJ3 stage ADJ3 liverpool) OR (late ADJ3 stage ADJ3 liverwort) OR (late ADJ3 stage ADJ3 liverworts) OR (late ADJ3 stage ADJ3 livera) OR (late ADJ3 stage ADJ3 livermultiscan) OR (late ADJ3 stage ADJ3 hepatic) OR (late ADJ3 stage ADJ3 hepatica) OR (late ADJ3 stage ADJ3 hepaticojejunostomy) OR (late ADJ3 stage ADJ3 hepatically) OR (late ADJ3 stage ADJ3 hepaticus) OR (late ADJ3 stage ADJ3 hepaticae)) or ESLD or ((esophageal ADJ3 varices) OR (esophageal ADJ3 variceal) OR (esophageal ADJ3 varicose) OR (esophageal ADJ3 varicosis) OR (esophageal ADJ3 varicocele) OR (esophageal ADJ3 varicocelectomy) OR (esophageal ADJ3 varicoid) OR (esophageal ADJ3 varicosely) OR (esophageal ADJ3 varicosities) OR (esophageal ADJ3 varication) OR (esophageal ADJ3 varicous) OR (esophageal ADJ3 varicosclerosation) OR (esophageal ADJ3 varicauda) OR (esophageal ADJ3 varicosity) OR (esophageal ADJ3 varicies) OR (esophageal ADJ3 varicial) OR (esophageal ADJ3 varicosed) OR (esophageal ADJ3 varicoses) OR (esophageal ADJ3 varicesa) OR (esophageal ADJ3 varice) OR (esophageal ADJ3 varicosclerosants) OR (esophagogastric ADJ3 varices) OR (esophagogastric ADJ3 variceal) OR (esophagogastric ADJ3 varicose) OR (esophagogastric ADJ3 varicosis) OR (esophagogastric ADJ3 varicocele) OR (esophagogastric ADJ3 varicocelectomy) OR (esophagogastric ADJ3 varicoid) OR (esophagogastric ADJ3 varicosely) OR (esophagogastric ADJ3 varicosities) OR (esophagogastric ADJ3 varication) OR (esophagogastric ADJ3 varicous) OR (esophagogastric ADJ3 varicosclerosation) OR (esophagogastric ADJ3 varicauda) OR (esophagogastric ADJ3 varicosity) OR (esophagogastric ADJ3 varicies) OR (esophagogastric ADJ3 varicial) OR (esophagogastric ADJ3 varicosed) OR (esophagogastric ADJ3 varicoses) OR (esophagogastric ADJ3 varicesa) OR (esophagogastric ADJ3 varice) OR (esophagogastric ADJ3 varicosclerosants) OR (esophagogastroduodenoscopy ADJ3 varices) OR (esophagogastroduodenoscopy ADJ3 variceal) OR (esophagogastroduodenoscopy ADJ3 varicose) OR (esophagogastroduodenoscopy ADJ3 varicosis) OR (esophagogastroduodenoscopy ADJ3 varicocele) OR (esophagogastroduodenoscopy ADJ3 varicocelectomy) OR (esophagogastroduodenoscopy ADJ3 varicoid) OR (esophagogastroduodenoscopy ADJ3 varicosely) OR (esophagogastroduodenoscopy ADJ3 varicosities) OR (esophagogastroduodenoscopy ADJ3 varication) OR (esophagogastroduodenoscopy ADJ3 varicous) OR (esophagogastroduodenoscopy ADJ3 varicosclerosation) OR (esophagogastroduodenoscopy ADJ3 varicauda) OR (esophagogastroduodenoscopy ADJ3 varicosity) OR (esophagogastroduodenoscopy ADJ3 varicies) OR (esophagogastroduodenoscopy ADJ3 varicial) OR (esophagogastroduodenoscopy ADJ3 varicosed) OR (esophagogastroduodenoscopy ADJ3 varicoses) OR (esophagogastroduodenoscopy ADJ3 varicesa) OR (esophagogastroduodenoscopy ADJ3 varice) OR (esophagogastroduodenoscopy ADJ3 varicosclerosants) OR (esophagus ADJ3 varices) OR (esophagus ADJ3 variceal) OR (esophagus ADJ3 varicose) OR (esophagus ADJ3 varicosis) OR (esophagus ADJ3 varicocele) OR (esophagus ADJ3 varicocelectomy) OR (esophagus ADJ3 varicoid) OR (esophagus ADJ3 varicosely) OR (esophagus ADJ3 varicosities) OR (esophagus ADJ3 varication) OR (esophagus ADJ3 varicous) OR (esophagus ADJ3 varicosclerosation) OR (esophagus ADJ3 varicauda) OR (esophagus ADJ3 varicosity) OR (esophagus ADJ3 varicies) OR (esophagus ADJ3 varicial) OR (esophagus ADJ3 varicosed) OR (esophagus ADJ3 varicoses) OR (esophagus ADJ3 varicesa) OR (esophagus ADJ3 varice) OR (esophagus ADJ3 varicosclerosants) OR (esophagitis ADJ3 varices) OR (esophagitis ADJ3 variceal) OR (esophagitis ADJ3 varicose) OR (esophagitis ADJ3 varicosis) OR (esophagitis ADJ3 varicocele) OR (esophagitis ADJ3 varicocelectomy) OR (esophagitis ADJ3 varicoid) OR (esophagitis ADJ3 varicosely) OR (esophagitis ADJ3 varicosities) OR (esophagitis ADJ3 varication) OR (esophagitis ADJ3 varicous) OR (esophagitis ADJ3 varicosclerosation) OR (esophagitis ADJ3 varicauda) OR (esophagitis ADJ3 varicosity) OR (esophagitis ADJ3 varicies) OR (esophagitis ADJ3 varicial) OR (esophagitis ADJ3 varicosed) OR (esophagitis ADJ3 varicoses) OR (esophagitis ADJ3 varicesa) OR (esophagitis ADJ3 varice) OR (esophagitis ADJ3 varicosclerosants) OR (esophagic ADJ3 varices) OR (esophagic ADJ3 variceal) OR (esophagic ADJ3 varicose) OR (esophagic ADJ3 varicosis) OR (esophagic ADJ3 varicocele) OR (esophagic ADJ3 varicocelectomy) OR (esophagic ADJ3 varicoid) OR (esophagic ADJ3 varicosely) OR (esophagic ADJ3 varicosities) OR (esophagic ADJ3 varication) OR (esophagic ADJ3 varicous) OR (esophagic ADJ3 varicosclerosation) OR (esophagic ADJ3 varicauda) OR (esophagic ADJ3 varicosity) OR (esophagic ADJ3 varicies) OR (esophagic ADJ3 varicial) OR (esophagic ADJ3 varicosed) OR (esophagic ADJ3 varicoses) OR (esophagic ADJ3 varicesa) OR (esophagic ADJ3 varice) OR (esophagic ADJ3 varicosclerosants) OR (esophago ADJ3 varices) OR (esophago ADJ3 variceal) OR (esophago ADJ3 varicose) OR (esophago ADJ3 varicosis) OR (esophago ADJ3 varicocele) OR (esophago ADJ3 varicocelectomy) OR (esophago ADJ3 varicoid) OR (esophago ADJ3 varicosely) OR (esophago ADJ3 varicosities) OR (esophago ADJ3 varication) OR (esophago ADJ3 varicous) OR (esophago ADJ3 varicosclerosation) OR (esophago ADJ3 varicauda) OR (esophago ADJ3 varicosity) OR (esophago ADJ3 varicies) OR (esophago ADJ3 varicial) OR (esophago ADJ3 varicosed) OR (esophago ADJ3 varicoses) OR (esophago ADJ3 varicesa) OR (esophago ADJ3 varice) OR (esophago ADJ3 varicosclerosants) OR (esophagojejunal ADJ3 varices) OR (esophagojejunal ADJ3 variceal) OR (esophagojejunal ADJ3 varicose) OR (esophagojejunal ADJ3 varicosis) OR (esophagojejunal ADJ3 varicocele) OR (esophagojejunal ADJ3 varicocelectomy) OR (esophagojejunal ADJ3 varicoid) OR (esophagojejunal ADJ3 varicosely) OR (esophagojejunal ADJ3 varicosities) OR (esophagojejunal ADJ3 varication) OR (esophagojejunal ADJ3 varicous) OR (esophagojejunal ADJ3 varicosclerosation) OR (esophagojejunal ADJ3 varicauda) OR (esophagojejunal ADJ3 varicosity) OR (esophagojejunal ADJ3 varicies) OR (esophagojejunal ADJ3 varicial) OR (esophagojejunal ADJ3 varicosed) OR (esophagojejunal ADJ3 varicoses) OR (esophagojejunal ADJ3 varicesa) OR (esophagojejunal ADJ3 varice) OR (esophagojejunal ADJ3 varicosclerosants) OR (esophagogastroduodenal ADJ3 varices) OR (esophagogastroduodenal ADJ3 variceal) OR (esophagogastroduodenal ADJ3 varicose) OR (esophagogastroduodenal ADJ3 varicosis) OR (esophagogastroduodenal ADJ3 varicocele) OR (esophagogastroduodenal ADJ3 varicocelectomy) OR (esophagogastroduodenal ADJ3 varicoid) OR (esophagogastroduodenal ADJ3 varicosely) OR (esophagogastroduodenal ADJ3 varicosities) OR (esophagogastroduodenal ADJ3 varication) OR (esophagogastroduodenal ADJ3 varicous) OR (esophagogastroduodenal ADJ3 varicosclerosation) OR (esophagogastroduodenal ADJ3 varicauda) OR (esophagogastroduodenal ADJ3 varicosity) OR (esophagogastroduodenal ADJ3 varicies) OR (esophagogastroduodenal ADJ3 varicial) OR (esophagogastroduodenal ADJ3 varicosed) OR (esophagogastroduodenal ADJ3 varicoses) OR (esophagogastroduodenal ADJ3 varicesa) OR (esophagogastroduodenal ADJ3 varice) OR (esophagogastroduodenal ADJ3 varicosclerosants) OR (esophagogastritis ADJ3 varices) OR (esophagogastritis ADJ3 variceal) OR (esophagogastritis ADJ3 varicose) OR (esophagogastritis ADJ3 varicosis) OR (esophagogastritis ADJ3 varicocele) OR (esophagogastritis ADJ3 varicocelectomy) OR (esophagogastritis ADJ3 varicoid) OR (esophagogastritis ADJ3 varicosely) OR (esophagogastritis ADJ3 varicosities) OR (esophagogastritis ADJ3 varication) OR (esophagogastritis ADJ3 varicous) OR (esophagogastritis ADJ3 varicosclerosation) OR (esophagogastritis ADJ3 varicauda) OR (esophagogastritis ADJ3 varicosity) OR (esophagogastritis ADJ3 varicies) OR (esophagogastritis ADJ3 varicial) OR (esophagogastritis ADJ3 varicosed) OR (esophagogastritis ADJ3 varicoses) OR (esophagogastritis ADJ3 varicesa) OR (esophagogastritis ADJ3 varice) OR (esophagogastritis ADJ3 varicosclerosants) OR (esophageo ADJ3 varices) OR (esophageo ADJ3 variceal) OR (esophageo ADJ3 varicose) OR (esophageo ADJ3 varicosis) OR (esophageo ADJ3 varicocele) OR (esophageo ADJ3 varicocelectomy) OR (esophageo ADJ3 varicoid) OR (esophageo ADJ3 varicosely) OR (esophageo ADJ3 varicosities) OR (esophageo ADJ3 varication) OR (esophageo ADJ3 varicous) OR (esophageo ADJ3 varicosclerosation) OR (esophageo ADJ3 varicauda) OR (esophageo ADJ3 varicosity) OR (esophageo ADJ3 varicies) OR (esophageo ADJ3 varicial) OR (esophageo ADJ3 varicosed) OR (esophageo ADJ3 varicoses) OR (esophageo ADJ3 varicesa) OR (esophageo ADJ3 varice) OR (esophageo ADJ3 varicosclerosants) OR (esophagectomy ADJ3 varices) OR (esophagectomy ADJ3 variceal) OR (esophagectomy ADJ3 varicose) OR (esophagectomy ADJ3 varicosis) OR (esophagectomy ADJ3 varicocele) OR (esophagectomy ADJ3 varicocelectomy) OR (esophagectomy ADJ3 varicoid) OR (esophagectomy ADJ3 varicosely) OR (esophagectomy ADJ3 varicosities) OR (esophagectomy ADJ3 varication) OR (esophagectomy ADJ3 varicous) OR (esophagectomy ADJ3 varicosclerosation) OR (esophagectomy ADJ3 varicauda) OR (esophagectomy ADJ3 varicosity) OR (esophagectomy ADJ3 varicies) OR (esophagectomy ADJ3 varicial) OR (esophagectomy ADJ3 varicosed) OR (esophagectomy ADJ3 varicoses) OR (esophagectomy ADJ3 varicesa) OR (esophagectomy ADJ3 varice) OR (esophagectomy ADJ3 varicosclerosants) OR (esophagology ADJ3 varices) OR (esophagology ADJ3 variceal) OR (esophagology ADJ3 varicose) OR (esophagology ADJ3 varicosis) OR (esophagology ADJ3 varicocele) OR (esophagology ADJ3 varicocelectomy) OR (esophagology ADJ3 varicoid) OR (esophagology ADJ3 varicosely) OR (esophagology ADJ3 varicosities) OR (esophagology ADJ3 varication) OR (esophagology ADJ3 varicous) OR (esophagology ADJ3 varicosclerosation) OR (esophagology ADJ3 varicauda) OR (esophagology ADJ3 varicosity) OR (esophagology ADJ3 varicies) OR (esophagology ADJ3 varicial) OR (esophagology ADJ3 varicosed) OR (esophagology ADJ3 varicoses) OR (esophagology ADJ3 varicesa) OR (esophagology ADJ3 varice) OR (esophagology ADJ3 varicosclerosants) OR (esophagastric ADJ3 varices) OR (esophagastric ADJ3 variceal) OR (esophagastric ADJ3 varicose) OR (esophagastric ADJ3 varicosis) OR (esophagastric ADJ3 varicocele) OR (esophagastric ADJ3 varicocelectomy) OR (esophagastric ADJ3 varicoid) OR (esophagastric ADJ3 varicosely) OR (esophagastric ADJ3 varicosities) OR (esophagastric ADJ3 varication) OR (esophagastric ADJ3 varicous) OR (esophagastric ADJ3 varicosclerosation) OR (esophagastric ADJ3 varicauda) OR (esophagastric ADJ3 varicosity) OR (esophagastric ADJ3 varicies) OR (esophagastric ADJ3 varicial) OR (esophagastric ADJ3 varicosed) OR (esophagastric ADJ3 varicoses) OR (esophagastric ADJ3 varicesa) OR (esophagastric ADJ3 varice) OR (esophagastric ADJ3 varicosclerosants) OR (esophagological ADJ3 varices) OR (esophagological ADJ3 variceal) OR (esophagological ADJ3 varicose) OR (esophagological ADJ3 varicosis) OR (esophagological ADJ3 varicocele) OR (esophagological ADJ3 varicocelectomy) OR (esophagological ADJ3 varicoid) OR (esophagological ADJ3 varicosely) OR (esophagological ADJ3 varicosities) OR (esophagological ADJ3 varication) OR (esophagological ADJ3 varicous) OR (esophagological ADJ3 varicosclerosation) OR (esophagological ADJ3 varicauda) OR (esophagological ADJ3 varicosity) OR (esophagological ADJ3 varicies) OR (esophagological ADJ3 varicial) OR (esophagological ADJ3 varicosed) OR (esophagological ADJ3 varicoses) OR (esophagological ADJ3 varicesa) OR (esophagological ADJ3 varice) OR (esophagological ADJ3 varicosclerosants) OR (esophagel ADJ3 varices) OR (esophagel ADJ3 variceal) OR (esophagel ADJ3 varicose) OR (esophagel ADJ3 varicosis) OR (esophagel ADJ3 varicocele) OR (esophagel ADJ3 varicocelectomy) OR (esophagel ADJ3 varicoid) OR (esophagel ADJ3 varicosely) OR (esophagel ADJ3 varicosities) OR (esophagel ADJ3 varication) OR (esophagel ADJ3 varicous) OR (esophagel ADJ3 varicosclerosation) OR (esophagel ADJ3 varicauda) OR (esophagel ADJ3 varicosity) OR (esophagel ADJ3 varicies) OR (esophagel ADJ3 varicial) OR (esophagel ADJ3 varicosed) OR (esophagel ADJ3 varicoses) OR (esophagel ADJ3 varicesa) OR (esophagel ADJ3 varice) OR (esophagel ADJ3 varicosclerosants) OR (esophagoscopy ADJ3 varices) OR (esophagoscopy ADJ3 variceal) OR (esophagoscopy ADJ3 varicose) OR (esophagoscopy ADJ3 varicosis) OR (esophagoscopy ADJ3 varicocele) OR (esophagoscopy ADJ3 varicocelectomy) OR (esophagoscopy ADJ3 varicoid) OR (esophagoscopy ADJ3 varicosely) OR (esophagoscopy ADJ3 varicosities) OR (esophagoscopy ADJ3 varication) OR (esophagoscopy ADJ3 varicous) OR (esophagoscopy ADJ3 varicosclerosation) OR (esophagoscopy ADJ3 varicauda) OR (esophagoscopy ADJ3 varicosity) OR (esophagoscopy ADJ3 varicies) OR (esophagoscopy ADJ3 varicial) OR (esophagoscopy ADJ3 varicosed) OR (esophagoscopy ADJ3 varicoses) OR (esophagoscopy ADJ3 varicesa) OR (esophagoscopy ADJ3 varice) OR (esophagoscopy ADJ3 varicosclerosants) OR (esophagocardial ADJ3 varices) OR (esophagocardial ADJ3 variceal) OR (esophagocardial ADJ3 varicose) OR (esophagocardial ADJ3 varicosis) OR (esophagocardial ADJ3 varicocele) OR (esophagocardial ADJ3 varicocelectomy) OR (esophagocardial ADJ3 varicoid) OR (esophagocardial ADJ3 varicosely) OR (esophagocardial ADJ3 varicosities) OR (esophagocardial ADJ3 varication) OR (esophagocardial ADJ3 varicous) OR (esophagocardial ADJ3 varicosclerosation) OR (esophagocardial ADJ3 varicauda) OR (esophagocardial ADJ3 varicosity) OR (esophagocardial ADJ3 varicies) OR (esophagocardial ADJ3 varicial) OR (esophagocardial ADJ3 varicosed) OR (esophagocardial ADJ3 varicoses) OR (esophagocardial ADJ3 varicesa) OR (esophagocardial ADJ3 varice) OR (esophagocardial ADJ3 varicosclerosants) OR (esophagal ADJ3 varices) OR (esophagal ADJ3 variceal) OR (esophagal ADJ3 varicose) OR (esophagal ADJ3 varicosis) OR (esophagal ADJ3 varicocele) OR (esophagal ADJ3 varicocelectomy) OR (esophagal ADJ3 varicoid) OR (esophagal ADJ3 varicosely) OR (esophagal ADJ3 varicosities) OR (esophagal ADJ3 varication) OR (esophagal ADJ3 varicous) OR (esophagal ADJ3 varicosclerosation) OR (esophagal ADJ3 varicauda) OR (esophagal ADJ3 varicosity) OR (esophagal ADJ3 varicies) OR (esophagal ADJ3 varicial) OR (esophagal ADJ3 varicosed) OR (esophagal ADJ3 varicoses) OR (esophagal ADJ3 varicesa) OR (esophagal ADJ3 varice) OR (esophagal ADJ3 varicosclerosants) OR (esophagocardia ADJ3 varices) OR (esophagocardia ADJ3 variceal) OR (esophagocardia ADJ3 varicose) OR (esophagocardia ADJ3 varicosis) OR (esophagocardia ADJ3 varicocele) OR (esophagocardia ADJ3 varicocelectomy) OR (esophagocardia ADJ3 varicoid) OR (esophagocardia ADJ3 varicosely) OR (esophagocardia ADJ3 varicosities) OR (esophagocardia ADJ3 varication) OR (esophagocardia ADJ3 varicous) OR (esophagocardia ADJ3 varicosclerosation) OR (esophagocardia ADJ3 varicauda) OR (esophagocardia ADJ3 varicosity) OR (esophagocardia ADJ3 varicies) OR (esophagocardia ADJ3 varicial) OR (esophagocardia ADJ3 varicosed) OR (esophagocardia ADJ3 varicoses) OR (esophagocardia ADJ3 varicesa) OR (esophagocardia ADJ3 varice) OR (esophagocardia ADJ3 varicosclerosants) OR (esophagopleural ADJ3 varices) OR (esophagopleural ADJ3 variceal) OR (esophagopleural ADJ3 varicose) OR (esophagopleural ADJ3 varicosis) OR (esophagopleural ADJ3 varicocele) OR (esophagopleural ADJ3 varicocelectomy) OR (esophagopleural ADJ3 varicoid) OR (esophagopleural ADJ3 varicosely) OR (esophagopleural ADJ3 varicosities) OR (esophagopleural ADJ3 varication) OR (esophagopleural ADJ3 varicous) OR (esophagopleural ADJ3 varicosclerosation) OR (esophagopleural ADJ3 varicauda) OR (esophagopleural ADJ3 varicosity) OR (esophagopleural ADJ3 varicies) OR (esophagopleural ADJ3 varicial) OR (esophagopleural ADJ3 varicosed) OR (esophagopleural ADJ3 varicoses) OR (esophagopleural ADJ3 varicesa) OR (esophagopleural ADJ3 varice) OR (esophagopleural ADJ3 varicosclerosants) OR (esophagobronchial ADJ3 varices) OR (esophagobronchial ADJ3 variceal) OR (esophagobronchial ADJ3 varicose) OR (esophagobronchial ADJ3 varicosis) OR (esophagobronchial ADJ3 varicocele) OR (esophagobronchial ADJ3 varicocelectomy) OR (esophagobronchial ADJ3 varicoid) OR (esophagobronchial ADJ3 varicosely) OR (esophagobronchial ADJ3 varicosities) OR (esophagobronchial ADJ3 varication) OR (esophagobronchial ADJ3 varicous) OR (esophagobronchial ADJ3 varicosclerosation) OR (esophagobronchial ADJ3 varicauda) OR (esophagobronchial ADJ3 varicosity) OR (esophagobronchial ADJ3 varicies) OR (esophagobronchial ADJ3 varicial) OR (esophagobronchial ADJ3 varicosed) OR (esophagobronchial ADJ3 varicoses) OR (esophagobronchial ADJ3 varicesa) OR (esophagobronchial ADJ3 varice) OR (esophagobronchial ADJ3 varicosclerosants) OR (esophagical ADJ3 varices) OR (esophagical ADJ3 variceal) OR (esophagical ADJ3 varicose) OR (esophagical ADJ3 varicosis) OR (esophagical ADJ3 varicocele) OR (esophagical ADJ3 varicocelectomy) OR (esophagical ADJ3 varicoid) OR (esophagical ADJ3 varicosely) OR (esophagical ADJ3 varicosities) OR (esophagical ADJ3 varication) OR (esophagical ADJ3 varicous) OR (esophagical ADJ3 varicosclerosation) OR (esophagical ADJ3 varicauda) OR (esophagical ADJ3 varicosity) OR (esophagical ADJ3 varicies) OR (esophagical ADJ3 varicial) OR (esophagical ADJ3 varicosed) OR (esophagical ADJ3 varicoses) OR (esophagical ADJ3 varicesa) OR (esophagical ADJ3 varice) OR (esophagical ADJ3 varicosclerosants) OR (esophagial ADJ3 varices) OR (esophagial ADJ3 variceal) OR (esophagial ADJ3 varicose) OR (esophagial ADJ3 varicosis) OR (esophagial ADJ3 varicocele) OR (esophagial ADJ3 varicocelectomy) OR (esophagial ADJ3 varicoid) OR (esophagial ADJ3 varicosely) OR (esophagial ADJ3 varicosities) OR (esophagial ADJ3 varication) OR (esophagial ADJ3 varicous) OR (esophagial ADJ3 varicosclerosation) OR (esophagial ADJ3 varicauda) OR (esophagial ADJ3 varicosity) OR (esophagial ADJ3 varicies) OR (esophagial ADJ3 varicial) OR (esophagial ADJ3 varicosed) OR (esophagial ADJ3 varicoses) OR (esophagial ADJ3 varicesa) OR (esophagial ADJ3 varice) OR (esophagial ADJ3 varicosclerosants) OR (esophagea ADJ3 varices) OR (esophagea ADJ3 variceal) OR (esophagea ADJ3 varicose) OR (esophagea ADJ3 varicosis) OR (esophagea ADJ3 varicocele) OR (esophagea ADJ3 varicocelectomy) OR (esophagea ADJ3 varicoid) OR (esophagea ADJ3 varicosely) OR (esophagea ADJ3 varicosities) OR (esophagea ADJ3 varication) OR (esophagea ADJ3 varicous) OR (esophagea ADJ3 varicosclerosation) OR (esophagea ADJ3 varicauda) OR (esophagea ADJ3 varicosity) OR (esophagea ADJ3 varicies) OR (esophagea ADJ3 varicial) OR (esophagea ADJ3 varicosed) OR (esophagea ADJ3 varicoses) OR (esophagea ADJ3 varicesa) OR (esophagea ADJ3 varice) OR (esophagea ADJ3 varicosclerosants) OR (esophageae ADJ3 varices) OR (esophageae ADJ3 variceal) OR (esophageae ADJ3 varicose) OR (esophageae ADJ3 varicosis) OR (esophageae ADJ3 varicocele) OR (esophageae ADJ3 varicocelectomy) OR (esophageae ADJ3 varicoid) OR (esophageae ADJ3 varicosely) OR (esophageae ADJ3 varicosities) OR (esophageae ADJ3 varication) OR (esophageae ADJ3 varicous) OR (esophageae ADJ3 varicosclerosation) OR (esophageae ADJ3 varicauda) OR (esophageae ADJ3 varicosity) OR (esophageae ADJ3 varicies) OR (esophageae ADJ3 varicial) OR (esophageae ADJ3 varicosed) OR (esophageae ADJ3 varicoses) OR (esophageae ADJ3 varicesa) OR (esophageae ADJ3 varice) OR (esophageae ADJ3 varicosclerosants)) or

((liver ADJ3 fibrosis) OR (liver ADJ3 failure) OR (liver ADJ3 failed) OR (liver ADJ3 fail) OR (liver ADJ3 failing) OR (liver ADJ3 failures) OR (liver ADJ3 fails) OR (liver ADJ3 insufficient) OR (liver ADJ3 insufficiently) OR (liver ADJ3 insufficiency) OR (liver ADJ3 insufficiencies) OR (liver ADJ3 dysfunction) OR (liver ADJ3 dysfunctional) OR (liver ADJ3 dysfunctions) OR (liver ADJ3 dysfunctionality) OR (liver ADJ3 dysfunctionassociated) OR (hepatic ADJ3 fibrosis) OR (hepatic ADJ3 failure) OR (hepatic ADJ3 failed) OR (hepatic ADJ3 fail) OR (hepatic ADJ3 failing) OR (hepatic ADJ3 failures) OR (hepatic ADJ3 fails) OR (hepatic ADJ3 insufficient) OR (hepatic ADJ3 insufficiently) OR (hepatic ADJ3 insufficiency) OR (hepatic ADJ3 insufficiencies) OR (hepatic ADJ3 dysfunction) OR (hepatic ADJ3 dysfunctional) OR (hepatic ADJ3 dysfunctions) OR (hepatic ADJ3 dysfunctionality) OR (hepatic ADJ3 dysfunctionassociated) OR (hepaticojejunostomy ADJ3 fibrosis) OR (hepaticojejunostomy ADJ3 failure) OR (hepaticojejunostomy ADJ3 failed) OR (hepaticojejunostomy ADJ3 fail) OR (hepaticojejunostomy ADJ3 failing) OR (hepaticojejunostomy ADJ3 failures) OR (hepaticojejunostomy ADJ3 fails) OR (hepaticojejunostomy ADJ3 insufficient) OR (hepaticojejunostomy ADJ3 insufficiently) OR (hepaticojejunostomy ADJ3 insufficiency) OR (hepaticojejunostomy ADJ3 insufficiencies) OR (hepaticojejunostomy ADJ3 dysfunction) OR (hepaticojejunostomy ADJ3 dysfunctional) OR (hepaticojejunostomy ADJ3 dysfunctions) OR (hepaticojejunostomy ADJ3 dysfunctionality) OR (hepaticojejunostomy ADJ3 dysfunctionassociated) OR (hepaticogastrostomy ADJ3 fibrosis) OR (hepaticogastrostomy ADJ3 failure) OR (hepaticogastrostomy ADJ3 failed) OR (hepaticogastrostomy ADJ3 fail) OR (hepaticogastrostomy ADJ3 failing) OR (hepaticogastrostomy ADJ3 failures) OR (hepaticogastrostomy ADJ3 fails) OR (hepaticogastrostomy ADJ3 insufficient) OR (hepaticogastrostomy ADJ3 insufficiently) OR (hepaticogastrostomy ADJ3 insufficiency) OR (hepaticogastrostomy ADJ3 insufficiencies) OR (hepaticogastrostomy ADJ3 dysfunction) OR (hepaticogastrostomy ADJ3 dysfunctional) OR (hepaticogastrostomy ADJ3 dysfunctions) OR (hepaticogastrostomy ADJ3 dysfunctionality) OR (hepaticogastrostomy ADJ3 dysfunctionassociated) OR (hepatica ADJ3 fibrosis) OR (hepatica ADJ3 failure) OR (hepatica ADJ3 failed) OR (hepatica ADJ3 fail) OR (hepatica ADJ3 failing) OR (hepatica ADJ3 failures) OR (hepatica ADJ3 fails) OR (hepatica ADJ3 insufficient) OR (hepatica ADJ3 insufficiently) OR (hepatica ADJ3 insufficiency) OR (hepatica ADJ3 insufficiencies) OR (hepatica ADJ3 dysfunction) OR (hepatica ADJ3 dysfunctional) OR (hepatica ADJ3 dysfunctions) OR (hepatica ADJ3 dysfunctionality) OR (hepatica ADJ3 dysfunctionassociated) OR (hepaticum ADJ3 fibrosis) OR (hepaticum ADJ3 failure) OR (hepaticum ADJ3 failed) OR (hepaticum ADJ3 fail) OR (hepaticum ADJ3 failing) OR (hepaticum ADJ3 failures) OR (hepaticum ADJ3 fails) OR (hepaticum ADJ3 insufficient) OR (hepaticum ADJ3 insufficiently) OR (hepaticum ADJ3 insufficiency) OR (hepaticum ADJ3 insufficiencies) OR (hepaticum ADJ3 dysfunction) OR (hepaticum ADJ3 dysfunctional) OR (hepaticum ADJ3 dysfunctions) OR (hepaticum ADJ3 dysfunctionality) OR (hepaticum ADJ3 dysfunctionassociated) OR (hepatically ADJ3 fibrosis) OR (hepatically ADJ3 failure) OR (hepatically ADJ3 failed) OR (hepatically ADJ3 fail) OR (hepatically ADJ3 failing) OR (hepatically ADJ3 failures) OR (hepatically ADJ3 fails) OR (hepatically ADJ3 insufficient) OR (hepatically ADJ3 insufficiently) OR (hepatically ADJ3 insufficiency) OR (hepatically ADJ3 insufficiencies) OR (hepatically ADJ3 dysfunction) OR (hepatically ADJ3 dysfunctional) OR (hepatically ADJ3 dysfunctions) OR (hepatically ADJ3 dysfunctionality) OR (hepatically ADJ3 dysfunctionassociated) OR (hepatics ADJ3 fibrosis) OR (hepatics ADJ3 failure) OR (hepatics ADJ3 failed) OR (hepatics ADJ3 fail) OR (hepatics ADJ3 failing) OR (hepatics ADJ3 failures) OR (hepatics ADJ3 fails) OR (hepatics ADJ3 insufficient) OR (hepatics ADJ3 insufficiently) OR (hepatics ADJ3 insufficiency) OR (hepatics ADJ3 insufficiencies) OR (hepatics ADJ3 dysfunction) OR (hepatics ADJ3 dysfunctional) OR (hepatics ADJ3 dysfunctions) OR (hepatics ADJ3 dysfunctionality) OR (hepatics ADJ3 dysfunctionassociated) OR (hepaticoduodenostomy ADJ3 fibrosis) OR (hepaticoduodenostomy ADJ3 failure) OR (hepaticoduodenostomy ADJ3 failed) OR (hepaticoduodenostomy ADJ3 fail) OR (hepaticoduodenostomy ADJ3 failing) OR (hepaticoduodenostomy ADJ3 failures) OR (hepaticoduodenostomy ADJ3 fails) OR (hepaticoduodenostomy ADJ3 insufficient) OR (hepaticoduodenostomy ADJ3 insufficiently) OR (hepaticoduodenostomy ADJ3 insufficiency) OR (hepaticoduodenostomy ADJ3 insufficiencies) OR (hepaticoduodenostomy ADJ3 dysfunction) OR (hepaticoduodenostomy ADJ3 dysfunctional) OR (hepaticoduodenostomy ADJ3 dysfunctions) OR (hepaticoduodenostomy ADJ3 dysfunctionality) OR (hepaticoduodenostomy ADJ3 dysfunctionassociated)) or "hepatopulmonary syndrome*" or "hepato-pulmonary syndrome*" or "hepatorenal syndrome*" or "hepato-renal syndrome*" or "portal hypertension" or ((variceal ADJ3 hemorrhage) OR (variceal ADJ3 hemorrhagic) OR (variceal ADJ3 hemorrhages) OR (variceal ADJ3 hemorrhaging) OR (variceal ADJ3 hemorrhagen) OR (variceal ADJ3 hemorrhaged) OR (variceal ADJ3 bleeding) OR (variceal ADJ3 bleed) OR (variceal ADJ3 bleeds) OR (variceal ADJ3 bleedings) OR (variceal ADJ3 bleeders) OR (variceal ADJ3 bleeder) OR (variceal ADJ3 bleedingin) OR (variceal ADJ3 bleedinginrelatedportal) OR (varices ADJ3 hemorrhage) OR (varices ADJ3 hemorrhagic) OR (varices ADJ3 hemorrhages) OR (varices ADJ3 hemorrhaging) OR (varices ADJ3 hemorrhagen) OR (varices ADJ3 hemorrhaged) OR (varices ADJ3 bleeding) OR (varices ADJ3 bleed) OR (varices ADJ3 bleeds) OR (varices ADJ3 bleedings) OR (varices ADJ3 bleeders) OR (varices ADJ3 bleeder) OR (varices ADJ3 bleedingin) OR (varices ADJ3 bleedinginrelatedportal) OR (varicose ADJ3 hemorrhage) OR (varicose ADJ3 hemorrhagic) OR (varicose ADJ3 hemorrhages) OR (varicose ADJ3 hemorrhaging) OR (varicose ADJ3 hemorrhagen) OR (varicose ADJ3 hemorrhaged) OR (varicose ADJ3 bleeding) OR (varicose ADJ3 bleed) OR (varicose ADJ3 bleeds) OR (varicose ADJ3 bleedings) OR (varicose ADJ3 bleeders) OR (varicose ADJ3 bleeder) OR (varicose ADJ3 bleedingin) OR (varicose ADJ3 bleedinginrelatedportal) OR (varicosis ADJ3 hemorrhage) OR (varicosis ADJ3 hemorrhagic) OR (varicosis ADJ3 hemorrhages) OR (varicosis ADJ3 hemorrhaging) OR (varicosis ADJ3 hemorrhagen) OR (varicosis ADJ3 hemorrhaged) OR (varicosis ADJ3 bleeding) OR (varicosis ADJ3 bleed) OR (varicosis ADJ3 bleeds) OR (varicosis ADJ3 bleedings) OR (varicosis ADJ3 bleeders) OR (varicosis ADJ3 bleeder) OR (varicosis ADJ3 bleedingin) OR (varicosis ADJ3 bleedinginrelatedportal) OR (varicosities ADJ3 hemorrhage) OR (varicosities ADJ3 hemorrhagic) OR (varicosities ADJ3 hemorrhages) OR (varicosities ADJ3 hemorrhaging) OR (varicosities ADJ3 hemorrhagen) OR (varicosities ADJ3 hemorrhaged) OR (varicosities ADJ3 bleeding) OR (varicosities ADJ3 bleed) OR (varicosities ADJ3 bleeds) OR (varicosities ADJ3 bleedings) OR (varicosities ADJ3 bleeders) OR (varicosities ADJ3 bleeder) OR (varicosities ADJ3 bleedingin) OR (varicosities ADJ3 bleedinginrelatedportal) OR (varicocele ADJ3 hemorrhage) OR (varicocele ADJ3 hemorrhagic) OR (varicocele ADJ3 hemorrhages) OR (varicocele ADJ3 hemorrhaging) OR (varicocele ADJ3 hemorrhagen) OR (varicocele ADJ3 hemorrhaged) OR (varicocele ADJ3 bleeding) OR (varicocele ADJ3 bleed) OR (varicocele ADJ3 bleeds) OR (varicocele ADJ3 bleedings) OR (varicocele ADJ3 bleeders) OR (varicocele ADJ3 bleeder) OR (varicocele ADJ3 bleedingin) OR (varicocele ADJ3 bleedinginrelatedportal) OR (varicocelectomy ADJ3 hemorrhage) OR (varicocelectomy ADJ3 hemorrhagic) OR (varicocelectomy ADJ3 hemorrhages) OR (varicocelectomy ADJ3 hemorrhaging) OR (varicocelectomy ADJ3 hemorrhagen) OR (varicocelectomy ADJ3 hemorrhaged) OR (varicocelectomy ADJ3 bleeding) OR (varicocelectomy ADJ3 bleed) OR (varicocelectomy ADJ3 bleeds) OR (varicocelectomy ADJ3 bleedings) OR (varicocelectomy ADJ3 bleeders) OR (varicocelectomy ADJ3 bleeder) OR (varicocelectomy ADJ3 bleedingin) OR (varicocelectomy ADJ3 bleedinginrelatedportal) OR (varice ADJ3 hemorrhage) OR (varice ADJ3 hemorrhagic) OR (varice ADJ3 hemorrhages) OR (varice ADJ3 hemorrhaging) OR (varice ADJ3 hemorrhagen) OR (varice ADJ3 hemorrhaged) OR (varice ADJ3 bleeding) OR (varice ADJ3 bleed) OR (varice ADJ3 bleeds) OR (varice ADJ3 bleedings) OR (varice ADJ3 bleeders) OR (varice ADJ3 bleeder) OR (varice ADJ3 bleedingin) OR (varice ADJ3 bleedinginrelatedportal) OR (varicosely ADJ3 hemorrhage) OR (varicosely ADJ3 hemorrhagic) OR (varicosely ADJ3 hemorrhages) OR (varicosely ADJ3 hemorrhaging) OR (varicosely ADJ3 hemorrhagen) OR (varicosely ADJ3 hemorrhaged) OR (varicosely ADJ3 bleeding) OR (varicosely ADJ3 bleed) OR (varicosely ADJ3 bleeds) OR (varicosely ADJ3 bleedings) OR (varicosely ADJ3 bleeders) OR (varicosely ADJ3 bleeder) OR (varicosely ADJ3 bleedingin) OR (varicosely ADJ3 bleedinginrelatedportal) OR (varicelliform ADJ3 hemorrhage) OR (varicelliform ADJ3 hemorrhagic) OR (varicelliform ADJ3 hemorrhages) OR (varicelliform ADJ3 hemorrhaging) OR (varicelliform ADJ3 hemorrhagen) OR (varicelliform ADJ3 hemorrhaged) OR (varicelliform ADJ3 bleeding) OR (varicelliform ADJ3 bleed) OR (varicelliform ADJ3 bleeds) OR (varicelliform ADJ3 bleedings) OR (varicelliform ADJ3 bleeders) OR (varicelliform ADJ3 bleeder) OR (varicelliform ADJ3 bleedingin) OR (varicelliform ADJ3 bleedinginrelatedportal) OR (varicous ADJ3 hemorrhage) OR (varicous ADJ3 hemorrhagic) OR (varicous ADJ3 hemorrhages) OR (varicous ADJ3 hemorrhaging) OR (varicous ADJ3 hemorrhagen) OR (varicous ADJ3 hemorrhaged) OR (varicous ADJ3 bleeding) OR (varicous ADJ3 bleed) OR (varicous ADJ3 bleeds) OR (varicous ADJ3 bleedings) OR (varicous ADJ3 bleeders) OR (varicous ADJ3 bleeder) OR (varicous ADJ3 bleedingin) OR (varicous ADJ3 bleedinginrelatedportal) OR (varicial ADJ3 hemorrhage) OR (varicial ADJ3 hemorrhagic) OR (varicial ADJ3 hemorrhages) OR (varicial ADJ3 hemorrhaging) OR (varicial ADJ3 hemorrhagen) OR (varicial ADJ3 hemorrhaged) OR (varicial ADJ3 bleeding) OR (varicial ADJ3 bleed) OR (varicial ADJ3 bleeds) OR (varicial ADJ3 bleedings) OR (varicial ADJ3 bleeders) OR (varicial ADJ3 bleeder) OR (varicial ADJ3 bleedingin) OR (varicial ADJ3 bleedinginrelatedportal) OR (varication ADJ3 hemorrhage) OR (varication ADJ3 hemorrhagic) OR (varication ADJ3 hemorrhages) OR (varication ADJ3 hemorrhaging) OR (varication ADJ3 hemorrhagen) OR (varication ADJ3 hemorrhaged) OR (varication ADJ3 bleeding) OR (varication ADJ3 bleed) OR (varication ADJ3 bleeds) OR (varication ADJ3 bleedings) OR (varication ADJ3 bleeders) OR (varication ADJ3 bleeder) OR (varication ADJ3 bleedingin) OR (varication ADJ3 bleedinginrelatedportal) OR (varicosed ADJ3 hemorrhage) OR (varicosed ADJ3 hemorrhagic) OR (varicosed ADJ3 hemorrhages) OR (varicosed ADJ3 hemorrhaging) OR (varicosed ADJ3 hemorrhagen) OR (varicosed ADJ3 hemorrhaged) OR (varicosed ADJ3 bleeding) OR (varicosed ADJ3 bleed) OR (varicosed ADJ3 bleeds) OR (varicosed ADJ3 bleedings) OR (varicosed ADJ3 bleeders) OR (varicosed ADJ3 bleeder) OR (varicosed ADJ3 bleedingin) OR (varicosed ADJ3 bleedinginrelatedportal) OR (varicoses ADJ3 hemorrhage) OR (varicoses ADJ3 hemorrhagic) OR (varicoses ADJ3 hemorrhages) OR (varicoses ADJ3 hemorrhaging) OR (varicoses ADJ3 hemorrhagen) OR (varicoses ADJ3 hemorrhaged) OR (varicoses ADJ3 bleeding) OR (varicoses ADJ3 bleed) OR (varicoses ADJ3 bleeds) OR (varicoses ADJ3 bleedings) OR (varicoses ADJ3 bleeders) OR (varicoses ADJ3 bleeder) OR (varicoses ADJ3 bleedingin) OR (varicoses ADJ3 bleedinginrelatedportal) OR (varicellar ADJ3 hemorrhage) OR (varicellar ADJ3 hemorrhagic) OR (varicellar ADJ3 hemorrhages) OR (varicellar ADJ3 hemorrhaging) OR (varicellar ADJ3 hemorrhagen) OR (varicellar ADJ3 hemorrhaged) OR (varicellar ADJ3 bleeding) OR (varicellar ADJ3 bleed) OR (varicellar ADJ3 bleeds) OR (varicellar ADJ3 bleedings) OR (varicellar ADJ3 bleeders) OR (varicellar ADJ3 bleeder) OR (varicellar ADJ3 bleedingin) OR (varicellar ADJ3 bleedinginrelatedportal) OR (varicocelectomies ADJ3 hemorrhage) OR (varicocelectomies ADJ3 hemorrhagic) OR (varicocelectomies ADJ3 hemorrhages) OR (varicocelectomies ADJ3 hemorrhaging) OR (varicocelectomies ADJ3 hemorrhagen) OR (varicocelectomies ADJ3 hemorrhaged) OR (varicocelectomies ADJ3 bleeding) OR (varicocelectomies ADJ3 bleed) OR (varicocelectomies ADJ3 bleeds) OR (varicocelectomies ADJ3 bleedings) OR (varicocelectomies ADJ3 bleeders) OR (varicocelectomies ADJ3 bleeder) OR (varicocelectomies ADJ3 bleedingin) OR (varicocelectomies ADJ3 bleedinginrelatedportal) OR (varicosclerosation ADJ3 hemorrhage) OR (varicosclerosation ADJ3 hemorrhagic) OR (varicosclerosation ADJ3 hemorrhages) OR (varicosclerosation ADJ3 hemorrhaging) OR (varicosclerosation ADJ3 hemorrhagen) OR (varicosclerosation ADJ3 hemorrhaged) OR (varicosclerosation ADJ3 bleeding) OR (varicosclerosation ADJ3 bleed) OR (varicosclerosation ADJ3 bleeds) OR (varicosclerosation ADJ3 bleedings) OR (varicosclerosation ADJ3 bleeders) OR (varicosclerosation ADJ3 bleeder) OR (varicosclerosation ADJ3 bleedingin) OR (varicosclerosation ADJ3 bleedinginrelatedportal) OR (varicoceles ADJ3 hemorrhage) OR (varicoceles ADJ3 hemorrhagic) OR (varicoceles ADJ3 hemorrhages) OR (varicoceles ADJ3 hemorrhaging) OR (varicoceles ADJ3 hemorrhagen) OR (varicoceles ADJ3 hemorrhaged) OR (varicoceles ADJ3 bleeding) OR (varicoceles ADJ3 bleed) OR (varicoceles ADJ3 bleeds) OR (varicoceles ADJ3 bleedings) OR (varicoceles ADJ3 bleeders) OR (varicoceles ADJ3 bleeder) OR (varicoceles ADJ3 bleedingin) OR (varicoceles ADJ3 bleedinginrelatedportal) OR (varicesa ADJ3 hemorrhage) OR (varicesa ADJ3 hemorrhagic) OR (varicesa ADJ3 hemorrhages) OR (varicesa ADJ3 hemorrhaging) OR (varicesa ADJ3 hemorrhagen) OR (varicesa ADJ3 hemorrhaged) OR (varicesa ADJ3 bleeding) OR (varicesa ADJ3 bleed) OR (varicesa ADJ3 bleeds) OR (varicesa ADJ3 bleedings) OR (varicesa ADJ3 bleeders) OR (varicesa ADJ3 bleeder) OR (varicesa ADJ3 bleedingin) OR (varicesa ADJ3 bleedinginrelatedportal) OR (varicosa ADJ3 hemorrhage) OR (varicosa ADJ3 hemorrhagic) OR (varicosa ADJ3 hemorrhages) OR (varicosa ADJ3 hemorrhaging) OR (varicosa ADJ3 hemorrhagen) OR (varicosa ADJ3 hemorrhaged) OR (varicosa ADJ3 bleeding) OR (varicosa ADJ3 bleed) OR (varicosa ADJ3 bleeds) OR (varicosa ADJ3 bleedings) OR (varicosa ADJ3 bleeders) OR (varicosa ADJ3 bleeder) OR (varicosa ADJ3 bleedingin) OR (varicosa ADJ3 bleedinginrelatedportal) OR (varicosity ADJ3 hemorrhage) OR (varicosity ADJ3 hemorrhagic) OR (varicosity ADJ3 hemorrhages) OR (varicosity ADJ3 hemorrhaging) OR (varicosity ADJ3 hemorrhagen) OR (varicosity ADJ3 hemorrhaged) OR (varicosity ADJ3 bleeding) OR (varicosity ADJ3 bleed) OR (varicosity ADJ3 bleeds) OR (varicosity ADJ3 bleedings) OR (varicosity ADJ3 bleeders) OR (varicosity ADJ3 bleeder) OR (varicosity ADJ3 bleedingin) OR (varicosity ADJ3 bleedinginrelatedportal) OR (variciform ADJ3 hemorrhage) OR (variciform ADJ3 hemorrhagic) OR (variciform ADJ3 hemorrhages) OR (variciform ADJ3 hemorrhaging) OR (variciform ADJ3 hemorrhagen) OR (variciform ADJ3 hemorrhaged) OR (variciform ADJ3 bleeding) OR (variciform ADJ3 bleed) OR (variciform ADJ3 bleeds) OR (variciform ADJ3 bleedings) OR (variciform ADJ3 bleeders) OR (variciform ADJ3 bleeder) OR (variciform ADJ3 bleedingin) OR (variciform ADJ3 bleedinginrelatedportal) OR (variconazole ADJ3 hemorrhage) OR (variconazole ADJ3 hemorrhagic) OR (variconazole ADJ3 hemorrhages) OR (variconazole ADJ3 hemorrhaging) OR (variconazole ADJ3 hemorrhagen) OR (variconazole ADJ3 hemorrhaged) OR (variconazole ADJ3 bleeding) OR (variconazole ADJ3 bleed) OR (variconazole ADJ3 bleeds) OR (variconazole ADJ3 bleedings) OR (variconazole ADJ3 bleeders) OR (variconazole ADJ3 bleeder) OR (variconazole ADJ3 bleedingin) OR (variconazole ADJ3 bleedinginrelatedportal) OR (varicies ADJ3 hemorrhage) OR (varicies ADJ3 hemorrhagic) OR (varicies ADJ3 hemorrhages) OR (varicies ADJ3 hemorrhaging) OR (varicies ADJ3 hemorrhagen) OR (varicies ADJ3 hemorrhaged) OR (varicies ADJ3 bleeding) OR (varicies ADJ3 bleed) OR (varicies ADJ3 bleeds) OR (varicies ADJ3 bleedings) OR (varicies ADJ3 bleeders) OR (varicies ADJ3 bleeder) OR (varicies ADJ3 bleedingin) OR (varicies ADJ3 bleedinginrelatedportal) OR (varical ADJ3 hemorrhage) OR (varical ADJ3 hemorrhagic) OR (varical ADJ3 hemorrhages) OR (varical ADJ3 hemorrhaging) OR (varical ADJ3 hemorrhagen) OR (varical ADJ3 hemorrhaged) OR (varical ADJ3 bleeding) OR (varical ADJ3 bleed) OR (varical ADJ3 bleeds) OR (varical ADJ3 bleedings) OR (varical ADJ3 bleeders) OR (varical ADJ3 bleeder) OR (varical ADJ3 bleedingin) OR (varical ADJ3 bleedinginrelatedportal) OR (varicelloid ADJ3 hemorrhage) OR (varicelloid ADJ3 hemorrhagic) OR (varicelloid ADJ3 hemorrhages) OR (varicelloid ADJ3 hemorrhaging) OR (varicelloid ADJ3 hemorrhagen) OR (varicelloid ADJ3 hemorrhaged) OR (varicelloid ADJ3 bleeding) OR (varicelloid ADJ3 bleed) OR (varicelloid ADJ3 bleeds) OR (varicelloid ADJ3 bleedings) OR (varicelloid ADJ3 bleeders) OR (varicelloid ADJ3 bleeder) OR (varicelloid ADJ3 bleedingin) OR (varicelloid ADJ3 bleedinginrelatedportal) OR (varicosum ADJ3 hemorrhage) OR (varicosum ADJ3 hemorrhagic) OR (varicosum ADJ3 hemorrhages) OR (varicosum ADJ3 hemorrhaging) OR (varicosum ADJ3 hemorrhagen) OR (varicosum ADJ3 hemorrhaged) OR (varicosum ADJ3 bleeding) OR (varicosum ADJ3 bleed) OR (varicosum ADJ3 bleeds) OR (varicosum ADJ3 bleedings) OR (varicosum ADJ3 bleeders) OR (varicosum ADJ3 bleeder) OR (varicosum ADJ3 bleedingin) OR (varicosum ADJ3 bleedinginrelatedportal) OR (varico ADJ3 hemorrhage) OR (varico ADJ3 hemorrhagic) OR (varico ADJ3 hemorrhages) OR (varico ADJ3 hemorrhaging) OR (varico ADJ3 hemorrhagen) OR (varico ADJ3 hemorrhaged) OR (varico ADJ3 bleeding) OR (varico ADJ3 bleed) OR (varico ADJ3 bleeds) OR (varico ADJ3 bleedings) OR (varico ADJ3 bleeders) OR (varico ADJ3 bleeder) OR (varico ADJ3 bleedingin) OR (varico ADJ3 bleedinginrelatedportal) OR (varicele ADJ3 hemorrhage) OR (varicele ADJ3 hemorrhagic) OR (varicele ADJ3 hemorrhages) OR (varicele ADJ3 hemorrhaging) OR (varicele ADJ3 hemorrhagen) OR (varicele ADJ3 hemorrhaged) OR (varicele ADJ3 bleeding) OR (varicele ADJ3 bleed) OR (varicele ADJ3 bleeds) OR (varicele ADJ3 bleedings) OR (varicele ADJ3 bleeders) OR (varicele ADJ3 bleeder) OR (varicele ADJ3 bleedingin) OR (varicele ADJ3 bleedinginrelatedportal) OR (varix ADJ3 hemorrhage) OR (varix ADJ3 hemorrhagic) OR (varix ADJ3 hemorrhages) OR (varix ADJ3 hemorrhaging) OR (varix ADJ3 hemorrhagen) OR (varix ADJ3 hemorrhaged) OR (varix ADJ3 bleeding) OR (varix ADJ3 bleed) OR (varix ADJ3 bleeds) OR (varix ADJ3 bleedings) OR (varix ADJ3 bleeders) OR (varix ADJ3 bleeder) OR (varix ADJ3 bleedingin) OR (varix ADJ3 bleedinginrelatedportal) OR (varixes ADJ3 hemorrhage) OR (varixes ADJ3 hemorrhagic) OR (varixes ADJ3 hemorrhages) OR (varixes ADJ3 hemorrhaging) OR (varixes ADJ3 hemorrhagen) OR (varixes ADJ3 hemorrhaged) OR (varixes ADJ3 bleeding) OR (varixes ADJ3 bleed) OR (varixes ADJ3 bleeds) OR (varixes ADJ3 bleedings) OR (varixes ADJ3 bleeders) OR (varixes ADJ3 bleeder) OR (varixes ADJ3 bleedingin) OR (varixes ADJ3 bleedinginrelatedportal))).ti,ab,kf.)

**Variant Rejected:**

Electronic*: Electronicmedical, electronicstructure, electroniclearning, electronicizing, electronicfans, electroniccase, electronicmaterials, electronicmessage

App (1^st^ occurrence) (note, since REMI can’t handle 3 letter truncation, this was converted to app or apps or appl*): Applicator, applied, applanation, appl, applicable, applying, applicability, applicant, apply, apples, applanatum, apple, applier, applies, appliances, applicants, applicabilities, appliable, applicated, applandimeric, appliedbfield, appletini, applied, appleton, appliques, applica
Health: healthineers, healthplix, healthdot, healthmail, healthcore, healthycow24, healthfacts, healthgrades, healthpathways, healthpartners, healthverity, healthcareer, healthjump, healthnuts, healtheast, healthiron, healthplus
Varic (1^st^ occurrence): Varicella, varicolored, variceropgrahy, varicula
Esophag: esophagogastrectomy, esophagoscope,  esophagogastroscopy, esophagram, esophagogastrojejeunoscopy, esophagogastroduodonoscopy, esophagogastroduod, esophagogastroduodenoscopies
Varic (2^nd^ occurrence): Varicella, varicellovirus, varicolored,
 **Included articles Ids (8/12):** 40552924, 36087864, 39037370, 35331941, 26070524, 39877863, 32876581, 28618192
Retrieved by original search: 8
Retreived by remi2.1 search: 8

#### Search 3 (Goeree et al.): https://pubmed.ncbi.nlm.nih.gov/42519403/

##### Search 3: Original MEDLINE Search: (3049 hits)

(exp Adipose Tissue/tr or exp Adipocytes/tr or ((exp "Autografts"/ or exp "tissue and organ harvesting"/ or exp "Transplantation, Autologous"/) and (exp "Adipose Tissue"/ or exp "Adipocytes"/ or (fat or fats or lipo* or lipid* or adipose).ti,ab,kf.)) or exp "lipectomy"/ or (((fat or fats or lipo* or lipid* or adipose) adj3 (trans* or injection* or graft* or autograft* or autotrans*)) or lipoinjection* or lipomodel* or lipotrans* or lipoaspirat* or liposuct* or lipectom* or lipofil* or AFT).ti,ab,kf.) and (exp "Mammaplasty"/ or exp Breast/su or (((Reconstruct* or Augment* or Oncoplasti*) adj4 Breast*) or mammoplast* or mastectom* or mastopex*).ti,ab,kf. or (exp "Transplant Donor Site"/ or ("donor site*" or "harvest site*").ti,ab,kf.))

##### Search 3: REMI 2 Proximity Search convert to Medline (3004 hits):

(exp Adipose Tissue/tr or exp Adipocytes/tr or ((exp "Autografts"/ or exp "tissue and organ harvesting"/ or exp "Transplantation, Autologous"/) and (exp "Adipose Tissue"/ or exp "Adipocytes"/ or (fat or fats or lipo* or lipid* or adipose).ti,ab,kf.)) or exp "lipectomy"/ or (((fat ADJ3 transport) OR (fat ADJ3 transplantation) OR (fat ADJ3 transfer) OR (fat ADJ3 transmission) OR (fat ADJ3 transporter) OR (fat ADJ3 transplant) OR (fat ADJ3 trans) OR (fat ADJ3 translocation) OR (fat ADJ3 translocator) OR (fat ADJ3 transfusions) OR (fat ADJ3 transfusion) OR (fat ADJ3 transferred) OR (fat ADJ3 transfused) OR (fat ADJ3 translocates) OR (fat ADJ3 transfat) OR (fat ADJ3 transbuccal) OR (fat ADJ3 transposition) OR (fat ADJ3 transportation) OR (fat ADJ3 transferable) OR (fat ADJ3 transpose) OR (fat ADJ3 transported) OR (fat ADJ3 transplanted) OR (fat ADJ3 translocalization) OR (fat ADJ3 transporting) OR (fat ADJ3 transfered) OR (fat ADJ3 transposed) OR (fat ADJ3 transplantable) OR (fat ADJ3 transplantations) OR (fat ADJ3 transfusional) OR (fat ADJ3 transfats) OR (fat ADJ3 transports) OR (fat ADJ3 transposable) OR (fat ADJ3 transfers) OR (fat ADJ3 transportable) OR (fat ADJ3 transplantology) OR (fat ADJ3 transplantability) OR (fat ADJ3 translocating) OR (fat ADJ3 transplanting) OR (fat ADJ3 transplants) OR (fat ADJ3 transferrable) OR (fat ADJ3 transfusing) OR (fat ADJ3 transferral) OR (fat ADJ3 transportability) OR (fat ADJ3 translocated) OR (fat ADJ3 transposing) OR (fat ADJ3 translocations) OR (fat ADJ3 injection) OR (fat ADJ3 injections) OR (fat ADJ3 injectionem) OR (fat ADJ3 injectional) OR (fat ADJ3 graft) OR (fat ADJ3 grafting) OR (fat ADJ3 grafts) OR (fat ADJ3 grafted) OR (fat ADJ3 graftable) OR (fat ADJ3 graftmaster) OR (fat ADJ3 graftversus) OR (fat ADJ3 grafton) OR (fat ADJ3 autograft) OR (fat ADJ3 autografts) OR (fat ADJ3 autografting) OR (fat ADJ3 autografted) OR (fat ADJ3 autotransplantation) OR (fat ADJ3 autotransporter) OR (fat ADJ3 autotransplant) OR (fat ADJ3 autotransfusion) OR (fat ADJ3 autotransporters) OR (fat ADJ3 autotransport) OR (fat ADJ3 autotransplants) OR (fat ADJ3 autotransplanting) OR (fat ADJ3 autotransplanted) OR (fats ADJ3 transport) OR (fats ADJ3 transplantation) OR (fats ADJ3 transfer) OR (fats ADJ3 transmission) OR (fats ADJ3 transporter) OR (fats ADJ3 transplant) OR (fats ADJ3 trans) OR (fats ADJ3 translocation) OR (fats ADJ3 translocator) OR (fats ADJ3 transfusions) OR (fats ADJ3 transfusion) OR (fats ADJ3 transferred) OR (fats ADJ3 transfused) OR (fats ADJ3 translocates) OR (fats ADJ3 transfat) OR (fats ADJ3 transbuccal) OR (fats ADJ3 transposition) OR (fats ADJ3 transportation) OR (fats ADJ3 transferable) OR (fats ADJ3 transpose) OR (fats ADJ3 transported) OR (fats ADJ3 transplanted) OR (fats ADJ3 translocalization) OR (fats ADJ3 transporting) OR (fats ADJ3 transfered) OR (fats ADJ3 transposed) OR (fats ADJ3 transplantable) OR (fats ADJ3 transplantations) OR (fats ADJ3 transfusional) OR (fats ADJ3 transfats) OR (fats ADJ3 transports) OR (fats ADJ3 transposable) OR (fats ADJ3 transfers) OR (fats ADJ3 transportable) OR (fats ADJ3 transplantology) OR (fats ADJ3 transplantability) OR (fats ADJ3 translocating) OR (fats ADJ3 transplanting) OR (fats ADJ3 transplants) OR (fats ADJ3 transferrable) OR (fats ADJ3 transfusing) OR (fats ADJ3 transferral) OR (fats ADJ3 transportability) OR (fats ADJ3 translocated) OR (fats ADJ3 transposing) OR (fats ADJ3 translocations) OR (fats ADJ3 injection) OR (fats ADJ3 injections) OR (fats ADJ3 injectionem) OR (fats ADJ3 injectional) OR (fats ADJ3 graft) OR (fats ADJ3 grafting) OR (fats ADJ3 grafts) OR (fats ADJ3 grafted) OR (fats ADJ3 graftable) OR (fats ADJ3 graftmaster) OR (fats ADJ3 graftversus) OR (fats ADJ3 grafton) OR (fats ADJ3 autograft) OR (fats ADJ3 autografts) OR (fats ADJ3 autografting) OR (fats ADJ3 autografted) OR (fats ADJ3 autotransplantation) OR (fats ADJ3 autotransporter) OR (fats ADJ3 autotransplant) OR (fats ADJ3 autotransfusion) OR (fats ADJ3 autotransporters) OR (fats ADJ3 autotransport) OR (fats ADJ3 autotransplants) OR (fats ADJ3 autotransplanting) OR (fats ADJ3 autotransplanted) OR (lipo ADJ3 transport) OR (lipo ADJ3 transplantation) OR (lipo ADJ3 transfer) OR (lipo ADJ3 transmission) OR (lipo ADJ3 transporter) OR (lipo ADJ3 transplant) OR (lipo ADJ3 trans) OR (lipo ADJ3 translocation) OR (lipo ADJ3 translocator) OR (lipo ADJ3 transfusions) OR (lipo ADJ3 transfusion) OR (lipo ADJ3 transferred) OR (lipo ADJ3 transfused) OR (lipo ADJ3 translocates) OR (lipo ADJ3 transfat) OR (lipo ADJ3 transbuccal) OR (lipo ADJ3 transposition) OR (lipo ADJ3 transportation) OR (lipo ADJ3 transferable) OR (lipo ADJ3 transpose) OR (lipo ADJ3 transported) OR (lipo ADJ3 transplanted) OR (lipo ADJ3 translocalization) OR (lipo ADJ3 transporting) OR (lipo ADJ3 transfered) OR (lipo ADJ3 transposed) OR (lipo ADJ3 transplantable) OR (lipo ADJ3 transplantations) OR (lipo ADJ3 transfusional) OR (lipo ADJ3 transfats) OR (lipo ADJ3 transports) OR (lipo ADJ3 transposable) OR (lipo ADJ3 transfers) OR (lipo ADJ3 transportable) OR (lipo ADJ3 transplantology) OR (lipo ADJ3 transplantability) OR (lipo ADJ3 translocating) OR (lipo ADJ3 transplanting) OR (lipo ADJ3 transplants) OR (lipo ADJ3 transferrable) OR (lipo ADJ3 transfusing) OR (lipo ADJ3 transferral) OR (lipo ADJ3 transportability) OR (lipo ADJ3 translocated) OR (lipo ADJ3 transposing) OR (lipo ADJ3 translocations) OR (lipo ADJ3 injection) OR (lipo ADJ3 injections) OR (lipo ADJ3 injectionem) OR (lipo ADJ3 injectional) OR (lipo ADJ3 graft) OR (lipo ADJ3 grafting) OR (lipo ADJ3 grafts) OR (lipo ADJ3 grafted) OR (lipo ADJ3 graftable) OR (lipo ADJ3 graftmaster) OR (lipo ADJ3 graftversus) OR (lipo ADJ3 grafton) OR (lipo ADJ3 autograft) OR (lipo ADJ3 autografts) OR (lipo ADJ3 autografting) OR (lipo ADJ3 autografted) OR (lipo ADJ3 autotransplantation) OR (lipo ADJ3 autotransporter) OR (lipo ADJ3 autotransplant) OR (lipo ADJ3 autotransfusion) OR (lipo ADJ3 autotransporters) OR (lipo ADJ3 autotransport) OR (lipo ADJ3 autotransplants) OR (lipo ADJ3 autotransplanting) OR (lipo ADJ3 autotransplanted) OR (lipoid ADJ3 transport) OR (lipoid ADJ3 transplantation) OR (lipoid ADJ3 transfer) OR (lipoid ADJ3 transmission) OR (lipoid ADJ3 transporter) OR (lipoid ADJ3 transplant) OR (lipoid ADJ3 trans) OR (lipoid ADJ3 translocation) OR (lipoid ADJ3 translocator) OR (lipoid ADJ3 transfusions) OR (lipoid ADJ3 transfusion) OR (lipoid ADJ3 transferred) OR (lipoid ADJ3 transfused) OR (lipoid ADJ3 translocates) OR (lipoid ADJ3 transfat) OR (lipoid ADJ3 transbuccal) OR (lipoid ADJ3 transposition) OR (lipoid ADJ3 transportation) OR (lipoid ADJ3 transferable) OR (lipoid ADJ3 transpose) OR (lipoid ADJ3 transported) OR (lipoid ADJ3 transplanted) OR (lipoid ADJ3 translocalization) OR (lipoid ADJ3 transporting) OR (lipoid ADJ3 transfered) OR (lipoid ADJ3 transposed) OR (lipoid ADJ3 transplantable) OR (lipoid ADJ3 transplantations) OR (lipoid ADJ3 transfusional) OR (lipoid ADJ3 transfats) OR (lipoid ADJ3 transports) OR (lipoid ADJ3 transposable) OR (lipoid ADJ3 transfers) OR (lipoid ADJ3 transportable) OR (lipoid ADJ3 transplantology) OR (lipoid ADJ3 transplantability) OR (lipoid ADJ3 translocating) OR (lipoid ADJ3 transplanting) OR (lipoid ADJ3 transplants) OR (lipoid ADJ3 transferrable) OR (lipoid ADJ3 transfusing) OR (lipoid ADJ3 transferral) OR (lipoid ADJ3 transportability) OR (lipoid ADJ3 translocated) OR (lipoid ADJ3 transposing) OR (lipoid ADJ3 translocations) OR (lipoid ADJ3 injection) OR (lipoid ADJ3 injections) OR (lipoid ADJ3 injectionem) OR (lipoid ADJ3 injectional) OR (lipoid ADJ3 graft) OR (lipoid ADJ3 grafting) OR (lipoid ADJ3 grafts) OR (lipoid ADJ3 grafted) OR (lipoid ADJ3 graftable) OR (lipoid ADJ3 graftmaster) OR (lipoid ADJ3 graftversus) OR (lipoid ADJ3 grafton) OR (lipoid ADJ3 autograft) OR (lipoid ADJ3 autografts) OR (lipoid ADJ3 autografting) OR (lipoid ADJ3 autografted) OR (lipoid ADJ3 autotransplantation) OR (lipoid ADJ3 autotransporter) OR (lipoid ADJ3 autotransplant) OR (lipoid ADJ3 autotransfusion) OR (lipoid ADJ3 autotransporters) OR (lipoid ADJ3 autotransport) OR (lipoid ADJ3 autotransplants) OR (lipoid ADJ3 autotransplanting) OR (lipoid ADJ3 autotransplanted) OR (lipoaspirate ADJ3 transport) OR (lipoaspirate ADJ3 transplantation) OR (lipoaspirate ADJ3 transfer) OR (lipoaspirate ADJ3 transmission) OR (lipoaspirate ADJ3 transporter) OR (lipoaspirate ADJ3 transplant) OR (lipoaspirate ADJ3 trans) OR (lipoaspirate ADJ3 translocation) OR (lipoaspirate ADJ3 translocator) OR (lipoaspirate ADJ3 transfusions) OR (lipoaspirate ADJ3 transfusion) OR (lipoaspirate ADJ3 transferred) OR (lipoaspirate ADJ3 transfused) OR (lipoaspirate ADJ3 translocates) OR (lipoaspirate ADJ3 transfat) OR (lipoaspirate ADJ3 transbuccal) OR (lipoaspirate ADJ3 transposition) OR (lipoaspirate ADJ3 transportation) OR (lipoaspirate ADJ3 transferable) OR (lipoaspirate ADJ3 transpose) OR (lipoaspirate ADJ3 transported) OR (lipoaspirate ADJ3 transplanted) OR (lipoaspirate ADJ3 translocalization) OR (lipoaspirate ADJ3 transporting) OR (lipoaspirate ADJ3 transfered) OR (lipoaspirate ADJ3 transposed) OR (lipoaspirate ADJ3 transplantable) OR (lipoaspirate ADJ3 transplantations) OR (lipoaspirate ADJ3 transfusional) OR (lipoaspirate ADJ3 transfats) OR (lipoaspirate ADJ3 transports) OR (lipoaspirate ADJ3 transposable) OR (lipoaspirate ADJ3 transfers) OR (lipoaspirate ADJ3 transportable) OR (lipoaspirate ADJ3 transplantology) OR (lipoaspirate ADJ3 transplantability) OR (lipoaspirate ADJ3 translocating) OR (lipoaspirate ADJ3 transplanting) OR (lipoaspirate ADJ3 transplants) OR (lipoaspirate ADJ3 transferrable) OR (lipoaspirate ADJ3 transfusing) OR (lipoaspirate ADJ3 transferral) OR (lipoaspirate ADJ3 transportability) OR (lipoaspirate ADJ3 translocated) OR (lipoaspirate ADJ3 transposing) OR (lipoaspirate ADJ3 translocations) OR (lipoaspirate ADJ3 injection) OR (lipoaspirate ADJ3 injections) OR (lipoaspirate ADJ3 injectionem) OR (lipoaspirate ADJ3 injectional) OR (lipoaspirate ADJ3 graft) OR (lipoaspirate ADJ3 grafting) OR (lipoaspirate ADJ3 grafts) OR (lipoaspirate ADJ3 grafted) OR (lipoaspirate ADJ3 graftable) OR (lipoaspirate ADJ3 graftmaster) OR (lipoaspirate ADJ3 graftversus) OR (lipoaspirate ADJ3 grafton) OR (lipoaspirate ADJ3 autograft) OR (lipoaspirate ADJ3 autografts) OR (lipoaspirate ADJ3 autografting) OR (lipoaspirate ADJ3 autografted) OR (lipoaspirate ADJ3 autotransplantation) OR (lipoaspirate ADJ3 autotransporter) OR (lipoaspirate ADJ3 autotransplant) OR (lipoaspirate ADJ3 autotransfusion) OR (lipoaspirate ADJ3 autotransporters) OR (lipoaspirate ADJ3 autotransport) OR (lipoaspirate ADJ3 autotransplants) OR (lipoaspirate ADJ3 autotransplanting) OR (lipoaspirate ADJ3 autotransplanted) OR (lipofilling ADJ3 transport) OR (lipofilling ADJ3 transplantation) OR (lipofilling ADJ3 transfer) OR (lipofilling ADJ3 transmission) OR (lipofilling ADJ3 transporter) OR (lipofilling ADJ3 transplant) OR (lipofilling ADJ3 trans) OR (lipofilling ADJ3 translocation) OR (lipofilling ADJ3 translocator) OR (lipofilling ADJ3 transfusions) OR (lipofilling ADJ3 transfusion) OR (lipofilling ADJ3 transferred) OR (lipofilling ADJ3 transfused) OR (lipofilling ADJ3 translocates) OR (lipofilling ADJ3 transfat) OR (lipofilling ADJ3 transbuccal) OR (lipofilling ADJ3 transposition) OR (lipofilling ADJ3 transportation) OR (lipofilling ADJ3 transferable) OR (lipofilling ADJ3 transpose) OR (lipofilling ADJ3 transported) OR (lipofilling ADJ3 transplanted) OR (lipofilling ADJ3 translocalization) OR (lipofilling ADJ3 transporting) OR (lipofilling ADJ3 transfered) OR (lipofilling ADJ3 transposed) OR (lipofilling ADJ3 transplantable) OR (lipofilling ADJ3 transplantations) OR (lipofilling ADJ3 transfusional) OR (lipofilling ADJ3 transfats) OR (lipofilling ADJ3 transports) OR (lipofilling ADJ3 transposable) OR (lipofilling ADJ3 transfers) OR (lipofilling ADJ3 transportable) OR (lipofilling ADJ3 transplantology) OR (lipofilling ADJ3 transplantability) OR (lipofilling ADJ3 translocating) OR (lipofilling ADJ3 transplanting) OR (lipofilling ADJ3 transplants) OR (lipofilling ADJ3 transferrable) OR (lipofilling ADJ3 transfusing) OR (lipofilling ADJ3 transferral) OR (lipofilling ADJ3 transportability) OR (lipofilling ADJ3 translocated) OR (lipofilling ADJ3 transposing) OR (lipofilling ADJ3 translocations) OR (lipofilling ADJ3 injection) OR (lipofilling ADJ3 injections) OR (lipofilling ADJ3 injectionem) OR (lipofilling ADJ3 injectional) OR (lipofilling ADJ3 graft) OR (lipofilling ADJ3 grafting) OR (lipofilling ADJ3 grafts) OR (lipofilling ADJ3 grafted) OR (lipofilling ADJ3 graftable) OR (lipofilling ADJ3 graftmaster) OR (lipofilling ADJ3 graftversus) OR (lipofilling ADJ3 grafton) OR (lipofilling ADJ3 autograft) OR (lipofilling ADJ3 autografts) OR (lipofilling ADJ3 autografting) OR (lipofilling ADJ3 autografted) OR (lipofilling ADJ3 autotransplantation) OR (lipofilling ADJ3 autotransporter) OR (lipofilling ADJ3 autotransplant) OR (lipofilling ADJ3 autotransfusion) OR (lipofilling ADJ3 autotransporters) OR (lipofilling ADJ3 autotransport) OR (lipofilling ADJ3 autotransplants) OR (lipofilling ADJ3 autotransplanting) OR (lipofilling ADJ3 autotransplanted) OR (lipoplasty ADJ3 transport) OR (lipoplasty ADJ3 transplantation) OR (lipoplasty ADJ3 transfer) OR (lipoplasty ADJ3 transmission) OR (lipoplasty ADJ3 transporter) OR (lipoplasty ADJ3 transplant) OR (lipoplasty ADJ3 trans) OR (lipoplasty ADJ3 translocation) OR (lipoplasty ADJ3 translocator) OR (lipoplasty ADJ3 transfusions) OR (lipoplasty ADJ3 transfusion) OR (lipoplasty ADJ3 transferred) OR (lipoplasty ADJ3 transfused) OR (lipoplasty ADJ3 translocates) OR (lipoplasty ADJ3 transfat) OR (lipoplasty ADJ3 transbuccal) OR (lipoplasty ADJ3 transposition) OR (lipoplasty ADJ3 transportation) OR (lipoplasty ADJ3 transferable) OR (lipoplasty ADJ3 transpose) OR (lipoplasty ADJ3 transported) OR (lipoplasty ADJ3 transplanted) OR (lipoplasty ADJ3 translocalization) OR (lipoplasty ADJ3 transporting) OR (lipoplasty ADJ3 transfered) OR (lipoplasty ADJ3 transposed) OR (lipoplasty ADJ3 transplantable) OR (lipoplasty ADJ3 transplantations) OR (lipoplasty ADJ3 transfusional) OR (lipoplasty ADJ3 transfats) OR (lipoplasty ADJ3 transports) OR (lipoplasty ADJ3 transposable) OR (lipoplasty ADJ3 transfers) OR (lipoplasty ADJ3 transportable) OR (lipoplasty ADJ3 transplantology) OR (lipoplasty ADJ3 transplantability) OR (lipoplasty ADJ3 translocating) OR (lipoplasty ADJ3 transplanting) OR (lipoplasty ADJ3 transplants) OR (lipoplasty ADJ3 transferrable) OR (lipoplasty ADJ3 transfusing) OR (lipoplasty ADJ3 transferral) OR (lipoplasty ADJ3 transportability) OR (lipoplasty ADJ3 translocated) OR (lipoplasty ADJ3 transposing) OR (lipoplasty ADJ3 translocations) OR (lipoplasty ADJ3 injection) OR (lipoplasty ADJ3 injections) OR (lipoplasty ADJ3 injectionem) OR (lipoplasty ADJ3 injectional) OR (lipoplasty ADJ3 graft) OR (lipoplasty ADJ3 grafting) OR (lipoplasty ADJ3 grafts) OR (lipoplasty ADJ3 grafted) OR (lipoplasty ADJ3 graftable) OR (lipoplasty ADJ3 graftmaster) OR (lipoplasty ADJ3 graftversus) OR (lipoplasty ADJ3 grafton) OR (lipoplasty ADJ3 autograft) OR (lipoplasty ADJ3 autografts) OR (lipoplasty ADJ3 autografting) OR (lipoplasty ADJ3 autografted) OR (lipoplasty ADJ3 autotransplantation) OR (lipoplasty ADJ3 autotransporter) OR (lipoplasty ADJ3 autotransplant) OR (lipoplasty ADJ3 autotransfusion) OR (lipoplasty ADJ3 autotransporters) OR (lipoplasty ADJ3 autotransport) OR (lipoplasty ADJ3 autotransplants) OR (lipoplasty ADJ3 autotransplanting) OR (lipoplasty ADJ3 autotransplanted) OR (lipotransfer ADJ3 transport) OR (lipotransfer ADJ3 transplantation) OR (lipotransfer ADJ3 transfer) OR (lipotransfer ADJ3 transmission) OR (lipotransfer ADJ3 transporter) OR (lipotransfer ADJ3 transplant) OR (lipotransfer ADJ3 trans) OR (lipotransfer ADJ3 translocation) OR (lipotransfer ADJ3 translocator) OR (lipotransfer ADJ3 transfusions) OR (lipotransfer ADJ3 transfusion) OR (lipotransfer ADJ3 transferred) OR (lipotransfer ADJ3 transfused) OR (lipotransfer ADJ3 translocates) OR (lipotransfer ADJ3 transfat) OR (lipotransfer ADJ3 transbuccal) OR (lipotransfer ADJ3 transposition) OR (lipotransfer ADJ3 transportation) OR (lipotransfer ADJ3 transferable) OR (lipotransfer ADJ3 transpose) OR (lipotransfer ADJ3 transported) OR (lipotransfer ADJ3 transplanted) OR (lipotransfer ADJ3 translocalization) OR (lipotransfer ADJ3 transporting) OR (lipotransfer ADJ3 transfered) OR (lipotransfer ADJ3 transposed) OR (lipotransfer ADJ3 transplantable) OR (lipotransfer ADJ3 transplantations) OR (lipotransfer ADJ3 transfusional) OR (lipotransfer ADJ3 transfats) OR (lipotransfer ADJ3 transports) OR (lipotransfer ADJ3 transposable) OR (lipotransfer ADJ3 transfers) OR (lipotransfer ADJ3 transportable) OR (lipotransfer ADJ3 transplantology) OR (lipotransfer ADJ3 transplantability) OR (lipotransfer ADJ3 translocating) OR (lipotransfer ADJ3 transplanting) OR (lipotransfer ADJ3 transplants) OR (lipotransfer ADJ3 transferrable) OR (lipotransfer ADJ3 transfusing) OR (lipotransfer ADJ3 transferral) OR (lipotransfer ADJ3 transportability) OR (lipotransfer ADJ3 translocated) OR (lipotransfer ADJ3 transposing) OR (lipotransfer ADJ3 translocations) OR (lipotransfer ADJ3 injection) OR (lipotransfer ADJ3 injections) OR (lipotransfer ADJ3 injectionem) OR (lipotransfer ADJ3 injectional) OR (lipotransfer ADJ3 graft) OR (lipotransfer ADJ3 grafting) OR (lipotransfer ADJ3 grafts) OR (lipotransfer ADJ3 grafted) OR (lipotransfer ADJ3 graftable) OR (lipotransfer ADJ3 graftmaster) OR (lipotransfer ADJ3 graftversus) OR (lipotransfer ADJ3 grafton) OR (lipotransfer ADJ3 autograft) OR (lipotransfer ADJ3 autografts) OR (lipotransfer ADJ3 autografting) OR (lipotransfer ADJ3 autografted) OR (lipotransfer ADJ3 autotransplantation) OR (lipotransfer ADJ3 autotransporter) OR (lipotransfer ADJ3 autotransplant) OR (lipotransfer ADJ3 autotransfusion) OR (lipotransfer ADJ3 autotransporters) OR (lipotransfer ADJ3 autotransport) OR (lipotransfer ADJ3 autotransplants) OR (lipotransfer ADJ3 autotransplanting) OR (lipotransfer ADJ3 autotransplanted) OR (lipomodeling ADJ3 transport) OR (lipomodeling ADJ3 transplantation) OR (lipomodeling ADJ3 transfer) OR (lipomodeling ADJ3 transmission) OR (lipomodeling ADJ3 transporter) OR (lipomodeling ADJ3 transplant) OR (lipomodeling ADJ3 trans) OR (lipomodeling ADJ3 translocation) OR (lipomodeling ADJ3 translocator) OR (lipomodeling ADJ3 transfusions) OR (lipomodeling ADJ3 transfusion) OR (lipomodeling ADJ3 transferred) OR (lipomodeling ADJ3 transfused) OR (lipomodeling ADJ3 translocates) OR (lipomodeling ADJ3 transfat) OR (lipomodeling ADJ3 transbuccal) OR (lipomodeling ADJ3 transposition) OR (lipomodeling ADJ3 transportation) OR (lipomodeling ADJ3 transferable) OR (lipomodeling ADJ3 transpose) OR (lipomodeling ADJ3 transported) OR (lipomodeling ADJ3 transplanted) OR (lipomodeling ADJ3 translocalization) OR (lipomodeling ADJ3 transporting) OR (lipomodeling ADJ3 transfered) OR (lipomodeling ADJ3 transposed) OR (lipomodeling ADJ3 transplantable) OR (lipomodeling ADJ3 transplantations) OR (lipomodeling ADJ3 transfusional) OR (lipomodeling ADJ3 transfats) OR (lipomodeling ADJ3 transports) OR (lipomodeling ADJ3 transposable) OR (lipomodeling ADJ3 transfers) OR (lipomodeling ADJ3 transportable) OR (lipomodeling ADJ3 transplantology) OR (lipomodeling ADJ3 transplantability) OR (lipomodeling ADJ3 translocating) OR (lipomodeling ADJ3 transplanting) OR (lipomodeling ADJ3 transplants) OR (lipomodeling ADJ3 transferrable) OR (lipomodeling ADJ3 transfusing) OR (lipomodeling ADJ3 transferral) OR (lipomodeling ADJ3 transportability) OR (lipomodeling ADJ3 translocated) OR (lipomodeling ADJ3 transposing) OR (lipomodeling ADJ3 translocations) OR (lipomodeling ADJ3 injection) OR (lipomodeling ADJ3 injections) OR (lipomodeling ADJ3 injectionem) OR (lipomodeling ADJ3 injectional) OR (lipomodeling ADJ3 graft) OR (lipomodeling ADJ3 grafting) OR (lipomodeling ADJ3 grafts) OR (lipomodeling ADJ3 grafted) OR (lipomodeling ADJ3 graftable) OR (lipomodeling ADJ3 graftmaster) OR (lipomodeling ADJ3 graftversus) OR (lipomodeling ADJ3 grafton) OR (lipomodeling ADJ3 autograft) OR (lipomodeling ADJ3 autografts) OR (lipomodeling ADJ3 autografting) OR (lipomodeling ADJ3 autografted) OR (lipomodeling ADJ3 autotransplantation) OR (lipomodeling ADJ3 autotransporter) OR (lipomodeling ADJ3 autotransplant) OR (lipomodeling ADJ3 autotransfusion) OR (lipomodeling ADJ3 autotransporters) OR (lipomodeling ADJ3 autotransport) OR (lipomodeling ADJ3 autotransplants) OR (lipomodeling ADJ3 autotransplanting) OR (lipomodeling ADJ3 autotransplanted) OR (lipoinjection ADJ3 transport) OR (lipoinjection ADJ3 transplantation) OR (lipoinjection ADJ3 transfer) OR (lipoinjection ADJ3 transmission) OR (lipoinjection ADJ3 transporter) OR (lipoinjection ADJ3 transplant) OR (lipoinjection ADJ3 trans) OR (lipoinjection ADJ3 translocation) OR (lipoinjection ADJ3 translocator) OR (lipoinjection ADJ3 transfusions) OR (lipoinjection ADJ3 transfusion) OR (lipoinjection ADJ3 transferred) OR (lipoinjection ADJ3 transfused) OR (lipoinjection ADJ3 translocates) OR (lipoinjection ADJ3 transfat) OR (lipoinjection ADJ3 transbuccal) OR (lipoinjection ADJ3 transposition) OR (lipoinjection ADJ3 transportation) OR (lipoinjection ADJ3 transferable) OR (lipoinjection ADJ3 transpose) OR (lipoinjection ADJ3 transported) OR (lipoinjection ADJ3 transplanted) OR (lipoinjection ADJ3 translocalization) OR (lipoinjection ADJ3 transporting) OR (lipoinjection ADJ3 transfered) OR (lipoinjection ADJ3 transposed) OR (lipoinjection ADJ3 transplantable) OR (lipoinjection ADJ3 transplantations) OR (lipoinjection ADJ3 transfusional) OR (lipoinjection ADJ3 transfats) OR (lipoinjection ADJ3 transports) OR (lipoinjection ADJ3 transposable) OR (lipoinjection ADJ3 transfers) OR (lipoinjection ADJ3 transportable) OR (lipoinjection ADJ3 transplantology) OR (lipoinjection ADJ3 transplantability) OR (lipoinjection ADJ3 translocating) OR (lipoinjection ADJ3 transplanting) OR (lipoinjection ADJ3 transplants) OR (lipoinjection ADJ3 transferrable) OR (lipoinjection ADJ3 transfusing) OR (lipoinjection ADJ3 transferral) OR (lipoinjection ADJ3 transportability) OR (lipoinjection ADJ3 translocated) OR (lipoinjection ADJ3 transposing) OR (lipoinjection ADJ3 translocations) OR (lipoinjection ADJ3 injection) OR (lipoinjection ADJ3 injections) OR (lipoinjection ADJ3 injectionem) OR (lipoinjection ADJ3 injectional) OR (lipoinjection ADJ3 graft) OR (lipoinjection ADJ3 grafting) OR (lipoinjection ADJ3 grafts) OR (lipoinjection ADJ3 grafted) OR (lipoinjection ADJ3 graftable) OR (lipoinjection ADJ3 graftmaster) OR (lipoinjection ADJ3 graftversus) OR (lipoinjection ADJ3 grafton) OR (lipoinjection ADJ3 autograft) OR (lipoinjection ADJ3 autografts) OR (lipoinjection ADJ3 autografting) OR (lipoinjection ADJ3 autografted) OR (lipoinjection ADJ3 autotransplantation) OR (lipoinjection ADJ3 autotransporter) OR (lipoinjection ADJ3 autotransplant) OR (lipoinjection ADJ3 autotransfusion) OR (lipoinjection ADJ3 autotransporters) OR (lipoinjection ADJ3 autotransport) OR (lipoinjection ADJ3 autotransplants) OR (lipoinjection ADJ3 autotransplanting) OR (lipoinjection ADJ3 autotransplanted) OR (lipoaspiration ADJ3 transport) OR (lipoaspiration ADJ3 transplantation) OR (lipoaspiration ADJ3 transfer) OR (lipoaspiration ADJ3 transmission) OR (lipoaspiration ADJ3 transporter) OR (lipoaspiration ADJ3 transplant) OR (lipoaspiration ADJ3 trans) OR (lipoaspiration ADJ3 translocation) OR (lipoaspiration ADJ3 translocator) OR (lipoaspiration ADJ3 transfusions) OR (lipoaspiration ADJ3 transfusion) OR (lipoaspiration ADJ3 transferred) OR (lipoaspiration ADJ3 transfused) OR (lipoaspiration ADJ3 translocates) OR (lipoaspiration ADJ3 transfat) OR (lipoaspiration ADJ3 transbuccal) OR (lipoaspiration ADJ3 transposition) OR (lipoaspiration ADJ3 transportation) OR (lipoaspiration ADJ3 transferable) OR (lipoaspiration ADJ3 transpose) OR (lipoaspiration ADJ3 transported) OR (lipoaspiration ADJ3 transplanted) OR (lipoaspiration ADJ3 translocalization) OR (lipoaspiration ADJ3 transporting) OR (lipoaspiration ADJ3 transfered) OR (lipoaspiration ADJ3 transposed) OR (lipoaspiration ADJ3 transplantable) OR (lipoaspiration ADJ3 transplantations) OR (lipoaspiration ADJ3 transfusional) OR (lipoaspiration ADJ3 transfats) OR (lipoaspiration ADJ3 transports) OR (lipoaspiration ADJ3 transposable) OR (lipoaspiration ADJ3 transfers) OR (lipoaspiration ADJ3 transportable) OR (lipoaspiration ADJ3 transplantology) OR (lipoaspiration ADJ3 transplantability) OR (lipoaspiration ADJ3 translocating) OR (lipoaspiration ADJ3 transplanting) OR (lipoaspiration ADJ3 transplants) OR (lipoaspiration ADJ3 transferrable) OR (lipoaspiration ADJ3 transfusing) OR (lipoaspiration ADJ3 transferral) OR (lipoaspiration ADJ3 transportability) OR (lipoaspiration ADJ3 translocated) OR (lipoaspiration ADJ3 transposing) OR (lipoaspiration ADJ3 translocations) OR (lipoaspiration ADJ3 injection) OR (lipoaspiration ADJ3 injections) OR (lipoaspiration ADJ3 injectionem) OR (lipoaspiration ADJ3 injectional) OR (lipoaspiration ADJ3 graft) OR (lipoaspiration ADJ3 grafting) OR (lipoaspiration ADJ3 grafts) OR (lipoaspiration ADJ3 grafted) OR (lipoaspiration ADJ3 graftable) OR (lipoaspiration ADJ3 graftmaster) OR (lipoaspiration ADJ3 graftversus) OR (lipoaspiration ADJ3 grafton) OR (lipoaspiration ADJ3 autograft) OR (lipoaspiration ADJ3 autografts) OR (lipoaspiration ADJ3 autografting) OR (lipoaspiration ADJ3 autografted) OR (lipoaspiration ADJ3 autotransplantation) OR (lipoaspiration ADJ3 autotransporter) OR (lipoaspiration ADJ3 autotransplant) OR (lipoaspiration ADJ3 autotransfusion) OR (lipoaspiration ADJ3 autotransporters) OR (lipoaspiration ADJ3 autotransport) OR (lipoaspiration ADJ3 autotransplants) OR (lipoaspiration ADJ3 autotransplanting) OR (lipoaspiration ADJ3 autotransplanted) OR (lipid ADJ3 transport) OR (lipid ADJ3 transplantation) OR (lipid ADJ3 transfer) OR (lipid ADJ3 transmission) OR (lipid ADJ3 transporter) OR (lipid ADJ3 transplant) OR (lipid ADJ3 trans) OR (lipid ADJ3 translocation) OR (lipid ADJ3 translocator) OR (lipid ADJ3 transfusions) OR (lipid ADJ3 transfusion) OR (lipid ADJ3 transferred) OR (lipid ADJ3 transfused) OR (lipid ADJ3 translocates) OR (lipid ADJ3 transfat) OR (lipid ADJ3 transbuccal) OR (lipid ADJ3 transposition) OR (lipid ADJ3 transportation) OR (lipid ADJ3 transferable) OR (lipid ADJ3 transpose) OR (lipid ADJ3 transported) OR (lipid ADJ3 transplanted) OR (lipid ADJ3 translocalization) OR (lipid ADJ3 transporting) OR (lipid ADJ3 transfered) OR (lipid ADJ3 transposed) OR (lipid ADJ3 transplantable) OR (lipid ADJ3 transplantations) OR (lipid ADJ3 transfusional) OR (lipid ADJ3 transfats) OR (lipid ADJ3 transports) OR (lipid ADJ3 transposable) OR (lipid ADJ3 transfers) OR (lipid ADJ3 transportable) OR (lipid ADJ3 transplantology) OR (lipid ADJ3 transplantability) OR (lipid ADJ3 translocating) OR (lipid ADJ3 transplanting) OR (lipid ADJ3 transplants) OR (lipid ADJ3 transferrable) OR (lipid ADJ3 transfusing) OR (lipid ADJ3 transferral) OR (lipid ADJ3 transportability) OR (lipid ADJ3 translocated) OR (lipid ADJ3 transposing) OR (lipid ADJ3 translocations) OR (lipid ADJ3 injection) OR (lipid ADJ3 injections) OR (lipid ADJ3 injectionem) OR (lipid ADJ3 injectional) OR (lipid ADJ3 graft) OR (lipid ADJ3 grafting) OR (lipid ADJ3 grafts) OR (lipid ADJ3 grafted) OR (lipid ADJ3 graftable) OR (lipid ADJ3 graftmaster) OR (lipid ADJ3 graftversus) OR (lipid ADJ3 grafton) OR (lipid ADJ3 autograft) OR (lipid ADJ3 autografts) OR (lipid ADJ3 autografting) OR (lipid ADJ3 autografted) OR (lipid ADJ3 autotransplantation) OR (lipid ADJ3 autotransporter) OR (lipid ADJ3 autotransplant) OR (lipid ADJ3 autotransfusion) OR (lipid ADJ3 autotransporters) OR (lipid ADJ3 autotransport) OR (lipid ADJ3 autotransplants) OR (lipid ADJ3 autotransplanting) OR (lipid ADJ3 autotransplanted) OR (lipids ADJ3 transport) OR (lipids ADJ3 transplantation) OR (lipids ADJ3 transfer) OR (lipids ADJ3 transmission) OR (lipids ADJ3 transporter) OR (lipids ADJ3 transplant) OR (lipids ADJ3 trans) OR (lipids ADJ3 translocation) OR (lipids ADJ3 translocator) OR (lipids ADJ3 transfusions) OR (lipids ADJ3 transfusion) OR (lipids ADJ3 transferred) OR (lipids ADJ3 transfused) OR (lipids ADJ3 translocates) OR (lipids ADJ3 transfat) OR (lipids ADJ3 transbuccal) OR (lipids ADJ3 transposition) OR (lipids ADJ3 transportation) OR (lipids ADJ3 transferable) OR (lipids ADJ3 transpose) OR (lipids ADJ3 transported) OR (lipids ADJ3 transplanted) OR (lipids ADJ3 translocalization) OR (lipids ADJ3 transporting) OR (lipids ADJ3 transfered) OR (lipids ADJ3 transposed) OR (lipids ADJ3 transplantable) OR (lipids ADJ3 transplantations) OR (lipids ADJ3 transfusional) OR (lipids ADJ3 transfats) OR (lipids ADJ3 transports) OR (lipids ADJ3 transposable) OR (lipids ADJ3 transfers) OR (lipids ADJ3 transportable) OR (lipids ADJ3 transplantology) OR (lipids ADJ3 transplantability) OR (lipids ADJ3 translocating) OR (lipids ADJ3 transplanting) OR (lipids ADJ3 transplants) OR (lipids ADJ3 transferrable) OR (lipids ADJ3 transfusing) OR (lipids ADJ3 transferral) OR (lipids ADJ3 transportability) OR (lipids ADJ3 translocated) OR (lipids ADJ3 transposing) OR (lipids ADJ3 translocations) OR (lipids ADJ3 injection) OR (lipids ADJ3 injections) OR (lipids ADJ3 injectionem) OR (lipids ADJ3 injectional) OR (lipids ADJ3 graft) OR (lipids ADJ3 grafting) OR (lipids ADJ3 grafts) OR (lipids ADJ3 grafted) OR (lipids ADJ3 graftable) OR (lipids ADJ3 graftmaster) OR (lipids ADJ3 graftversus) OR (lipids ADJ3 grafton) OR (lipids ADJ3 autograft) OR (lipids ADJ3 autografts) OR (lipids ADJ3 autografting) OR (lipids ADJ3 autografted) OR (lipids ADJ3 autotransplantation) OR (lipids ADJ3 autotransporter) OR (lipids ADJ3 autotransplant) OR (lipids ADJ3 autotransfusion) OR (lipids ADJ3 autotransporters) OR (lipids ADJ3 autotransport) OR (lipids ADJ3 autotransplants) OR (lipids ADJ3 autotransplanting) OR (lipids ADJ3 autotransplanted) OR (lipidic ADJ3 transport) OR (lipidic ADJ3 transplantation) OR (lipidic ADJ3 transfer) OR (lipidic ADJ3 transmission) OR (lipidic ADJ3 transporter) OR (lipidic ADJ3 transplant) OR (lipidic ADJ3 trans) OR (lipidic ADJ3 translocation) OR (lipidic ADJ3 translocator) OR (lipidic ADJ3 transfusions) OR (lipidic ADJ3 transfusion) OR (lipidic ADJ3 transferred) OR (lipidic ADJ3 transfused) OR (lipidic ADJ3 translocates) OR (lipidic ADJ3 transfat) OR (lipidic ADJ3 transbuccal) OR (lipidic ADJ3 transposition) OR (lipidic ADJ3 transportation) OR (lipidic ADJ3 transferable) OR (lipidic ADJ3 transpose) OR (lipidic ADJ3 transported) OR (lipidic ADJ3 transplanted) OR (lipidic ADJ3 translocalization) OR (lipidic ADJ3 transporting) OR (lipidic ADJ3 transfered) OR (lipidic ADJ3 transposed) OR (lipidic ADJ3 transplantable) OR (lipidic ADJ3 transplantations) OR (lipidic ADJ3 transfusional) OR (lipidic ADJ3 transfats) OR (lipidic ADJ3 transports) OR (lipidic ADJ3 transposable) OR (lipidic ADJ3 transfers) OR (lipidic ADJ3 transportable) OR (lipidic ADJ3 transplantology) OR (lipidic ADJ3 transplantability) OR (lipidic ADJ3 translocating) OR (lipidic ADJ3 transplanting) OR (lipidic ADJ3 transplants) OR (lipidic ADJ3 transferrable) OR (lipidic ADJ3 transfusing) OR (lipidic ADJ3 transferral) OR (lipidic ADJ3 transportability) OR (lipidic ADJ3 translocated) OR (lipidic ADJ3 transposing) OR (lipidic ADJ3 translocations) OR (lipidic ADJ3 injection) OR (lipidic ADJ3 injections) OR (lipidic ADJ3 injectionem) OR (lipidic ADJ3 injectional) OR (lipidic ADJ3 graft) OR (lipidic ADJ3 grafting) OR (lipidic ADJ3 grafts) OR (lipidic ADJ3 grafted) OR (lipidic ADJ3 graftable) OR (lipidic ADJ3 graftmaster) OR (lipidic ADJ3 graftversus) OR (lipidic ADJ3 grafton) OR (lipidic ADJ3 autograft) OR (lipidic ADJ3 autografts) OR (lipidic ADJ3 autografting) OR (lipidic ADJ3 autografted) OR (lipidic ADJ3 autotransplantation) OR (lipidic ADJ3 autotransporter) OR (lipidic ADJ3 autotransplant) OR (lipidic ADJ3 autotransfusion) OR (lipidic ADJ3 autotransporters) OR (lipidic ADJ3 autotransport) OR (lipidic ADJ3 autotransplants) OR (lipidic ADJ3 autotransplanting) OR (lipidic ADJ3 autotransplanted) OR (lipidemic ADJ3 transport) OR (lipidemic ADJ3 transplantation) OR (lipidemic ADJ3 transfer) OR (lipidemic ADJ3 transmission) OR (lipidemic ADJ3 transporter) OR (lipidemic ADJ3 transplant) OR (lipidemic ADJ3 trans) OR (lipidemic ADJ3 translocation) OR (lipidemic ADJ3 translocator) OR (lipidemic ADJ3 transfusions) OR (lipidemic ADJ3 transfusion) OR (lipidemic ADJ3 transferred) OR (lipidemic ADJ3 transfused) OR (lipidemic ADJ3 translocates) OR (lipidemic ADJ3 transfat) OR (lipidemic ADJ3 transbuccal) OR (lipidemic ADJ3 transposition) OR (lipidemic ADJ3 transportation) OR (lipidemic ADJ3 transferable) OR (lipidemic ADJ3 transpose) OR (lipidemic ADJ3 transported) OR (lipidemic ADJ3 transplanted) OR (lipidemic ADJ3 translocalization) OR (lipidemic ADJ3 transporting) OR (lipidemic ADJ3 transfered) OR (lipidemic ADJ3 transposed) OR (lipidemic ADJ3 transplantable) OR (lipidemic ADJ3 transplantations) OR (lipidemic ADJ3 transfusional) OR (lipidemic ADJ3 transfats) OR (lipidemic ADJ3 transports) OR (lipidemic ADJ3 transposable) OR (lipidemic ADJ3 transfers) OR (lipidemic ADJ3 transportable) OR (lipidemic ADJ3 transplantology) OR (lipidemic ADJ3 transplantability) OR (lipidemic ADJ3 translocating) OR (lipidemic ADJ3 transplanting) OR (lipidemic ADJ3 transplants) OR (lipidemic ADJ3 transferrable) OR (lipidemic ADJ3 transfusing) OR (lipidemic ADJ3 transferral) OR (lipidemic ADJ3 transportability) OR (lipidemic ADJ3 translocated) OR (lipidemic ADJ3 transposing) OR (lipidemic ADJ3 translocations) OR (lipidemic ADJ3 injection) OR (lipidemic ADJ3 injections) OR (lipidemic ADJ3 injectionem) OR (lipidemic ADJ3 injectional) OR (lipidemic ADJ3 graft) OR (lipidemic ADJ3 grafting) OR (lipidemic ADJ3 grafts) OR (lipidemic ADJ3 grafted) OR (lipidemic ADJ3 graftable) OR (lipidemic ADJ3 graftmaster) OR (lipidemic ADJ3 graftversus) OR (lipidemic ADJ3 grafton) OR (lipidemic ADJ3 autograft) OR (lipidemic ADJ3 autografts) OR (lipidemic ADJ3 autografting) OR (lipidemic ADJ3 autografted) OR (lipidemic ADJ3 autotransplantation) OR (lipidemic ADJ3 autotransporter) OR (lipidemic ADJ3 autotransplant) OR (lipidemic ADJ3 autotransfusion) OR (lipidemic ADJ3 autotransporters) OR (lipidemic ADJ3 autotransport) OR (lipidemic ADJ3 autotransplants) OR (lipidemic ADJ3 autotransplanting) OR (lipidemic ADJ3 autotransplanted) OR (adipose ADJ3 transport) OR (adipose ADJ3 transplantation) OR (adipose ADJ3 transfer) OR (adipose ADJ3 transmission) OR (adipose ADJ3 transporter) OR (adipose ADJ3 transplant) OR (adipose ADJ3 trans) OR (adipose ADJ3 translocation) OR (adipose ADJ3 translocator) OR (adipose ADJ3 transfusions) OR (adipose ADJ3 transfusion) OR (adipose ADJ3 transferred) OR (adipose ADJ3 transfused) OR (adipose ADJ3 translocates) OR (adipose ADJ3 transfat) OR (adipose ADJ3 transbuccal) OR (adipose ADJ3 transposition) OR (adipose ADJ3 transportation) OR (adipose ADJ3 transferable) OR (adipose ADJ3 transpose) OR (adipose ADJ3 transported) OR (adipose ADJ3 transplanted) OR (adipose ADJ3 translocalization) OR (adipose ADJ3 transporting) OR (adipose ADJ3 transfered) OR (adipose ADJ3 transposed) OR (adipose ADJ3 transplantable) OR (adipose ADJ3 transplantations) OR (adipose ADJ3 transfusional) OR (adipose ADJ3 transfats) OR (adipose ADJ3 transports) OR (adipose ADJ3 transposable) OR (adipose ADJ3 transfers) OR (adipose ADJ3 transportable) OR (adipose ADJ3 transplantology) OR (adipose ADJ3 transplantability) OR (adipose ADJ3 translocating) OR (adipose ADJ3 transplanting) OR (adipose ADJ3 transplants) OR (adipose ADJ3 transferrable) OR (adipose ADJ3 transfusing) OR (adipose ADJ3 transferral) OR (adipose ADJ3 transportability) OR (adipose ADJ3 translocated) OR (adipose ADJ3 transposing) OR (adipose ADJ3 translocations) OR (adipose ADJ3 injection) OR (adipose ADJ3 injections) OR (adipose ADJ3 injectionem) OR (adipose ADJ3 injectional) OR (adipose ADJ3 graft) OR (adipose ADJ3 grafting) OR (adipose ADJ3 grafts) OR (adipose ADJ3 grafted) OR (adipose ADJ3 graftable) OR (adipose ADJ3 graftmaster) OR (adipose ADJ3 graftversus) OR (adipose ADJ3 grafton) OR (adipose ADJ3 autograft) OR (adipose ADJ3 autografts) OR (adipose ADJ3 autografting) OR (adipose ADJ3 autografted) OR (adipose ADJ3 autotransplantation) OR (adipose ADJ3 autotransporter) OR (adipose ADJ3 autotransplant) OR (adipose ADJ3 autotransfusion) OR (adipose ADJ3 autotransporters) OR (adipose ADJ3 autotransport) OR (adipose ADJ3 autotransplants) OR (adipose ADJ3 autotransplanting) OR (adipose ADJ3 autotransplanted)) or lipoinjection* or lipomodel* or lipotrans* or lipoaspirat* or liposuct* or lipectom* or lipofil* or AFT).ti,ab,kf.) and (exp "Mammaplasty"/ or exp Breast/su or (((reconstruction ADJ4 breast) OR (reconstruction ADJ4 breasts) OR (reconstruction ADJ4 breastfeeding) OR (reconstruction ADJ4 breastmilk) OR (reconstruction ADJ4 breaststroke) OR (reconstruction ADJ4 breasting) OR (reconstruction ADJ4 breastcancer) OR (reconstruction ADJ4 breastsurgery) OR (reconstruction ADJ4 breasted) OR (reconstruction ADJ4 breastplate) OR (reconstruction ADJ4 breastchoice) OR (reconstruction ADJ4 breastfeed) OR (reconstruction ADJ4 breastreconstruction) OR (reconstruction ADJ4 breastconservingsurgery) OR (reconstruction ADJ4 breastics) OR (reconstruction ADJ4 breast09) OR (reconstruction ADJ4 breastfed) OR (reconstruction ADJ4 breastq) OR (reconstruction ADJ4 breasttumor) OR (reconstruction ADJ4 breastbone) OR (reconstruction ADJ4 breastlike) OR (reconstructive ADJ4 breast) OR (reconstructive ADJ4 breasts) OR (reconstructive ADJ4 breastfeeding) OR (reconstructive ADJ4 breastmilk) OR (reconstructive ADJ4 breaststroke) OR (reconstructive ADJ4 breasting) OR (reconstructive ADJ4 breastcancer) OR (reconstructive ADJ4 breastsurgery) OR (reconstructive ADJ4 breasted) OR (reconstructive ADJ4 breastplate) OR (reconstructive ADJ4 breastchoice) OR (reconstructive ADJ4 breastfeed) OR (reconstructive ADJ4 breastreconstruction) OR (reconstructive ADJ4 breastconservingsurgery) OR (reconstructive ADJ4 breastics) OR (reconstructive ADJ4 breast09) OR (reconstructive ADJ4 breastfed) OR (reconstructive ADJ4 breastq) OR (reconstructive ADJ4 breasttumor) OR (reconstructive ADJ4 breastbone) OR (reconstructive ADJ4 breastlike) OR (reconstructed ADJ4 breast) OR (reconstructed ADJ4 breasts) OR (reconstructed ADJ4 breastfeeding) OR (reconstructed ADJ4 breastmilk) OR (reconstructed ADJ4 breaststroke) OR (reconstructed ADJ4 breasting) OR (reconstructed ADJ4 breastcancer) OR (reconstructed ADJ4 breastsurgery) OR (reconstructed ADJ4 breasted) OR (reconstructed ADJ4 breastplate) OR (reconstructed ADJ4 breastchoice) OR (reconstructed ADJ4 breastfeed) OR (reconstructed ADJ4 breastreconstruction) OR (reconstructed ADJ4 breastconservingsurgery) OR (reconstructed ADJ4 breastics) OR (reconstructed ADJ4 breast09) OR (reconstructed ADJ4 breastfed) OR (reconstructed ADJ4 breastq) OR (reconstructed ADJ4 breasttumor) OR (reconstructed ADJ4 breastbone) OR (reconstructed ADJ4 breastlike) OR (reconstructions ADJ4 breast) OR (reconstructions ADJ4 breasts) OR (reconstructions ADJ4 breastfeeding) OR (reconstructions ADJ4 breastmilk) OR (reconstructions ADJ4 breaststroke) OR (reconstructions ADJ4 breasting) OR (reconstructions ADJ4 breastcancer) OR (reconstructions ADJ4 breastsurgery) OR (reconstructions ADJ4 breasted) OR (reconstructions ADJ4 breastplate) OR (reconstructions ADJ4 breastchoice) OR (reconstructions ADJ4 breastfeed) OR (reconstructions ADJ4 breastreconstruction) OR (reconstructions ADJ4 breastconservingsurgery) OR (reconstructions ADJ4 breastics) OR (reconstructions ADJ4 breast09) OR (reconstructions ADJ4 breastfed) OR (reconstructions ADJ4 breastq) OR (reconstructions ADJ4 breasttumor) OR (reconstructions ADJ4 breastbone) OR (reconstructions ADJ4 breastlike) OR (reconstructing ADJ4 breast) OR (reconstructing ADJ4 breasts) OR (reconstructing ADJ4 breastfeeding) OR (reconstructing ADJ4 breastmilk) OR (reconstructing ADJ4 breaststroke) OR (reconstructing ADJ4 breasting) OR (reconstructing ADJ4 breastcancer) OR (reconstructing ADJ4 breastsurgery) OR (reconstructing ADJ4 breasted) OR (reconstructing ADJ4 breastplate) OR (reconstructing ADJ4 breastchoice) OR (reconstructing ADJ4 breastfeed) OR (reconstructing ADJ4 breastreconstruction) OR (reconstructing ADJ4 breastconservingsurgery) OR (reconstructing ADJ4 breastics) OR (reconstructing ADJ4 breast09) OR (reconstructing ADJ4 breastfed) OR (reconstructing ADJ4 breastq) OR (reconstructing ADJ4 breasttumor) OR (reconstructing ADJ4 breastbone) OR (reconstructing ADJ4 breastlike) OR (reconstructs ADJ4 breast) OR (reconstructs ADJ4 breasts) OR (reconstructs ADJ4 breastfeeding) OR (reconstructs ADJ4 breastmilk) OR (reconstructs ADJ4 breaststroke) OR (reconstructs ADJ4 breasting) OR (reconstructs ADJ4 breastcancer) OR (reconstructs ADJ4 breastsurgery) OR (reconstructs ADJ4 breasted) OR (reconstructs ADJ4 breastplate) OR (reconstructs ADJ4 breastchoice) OR (reconstructs ADJ4 breastfeed) OR (reconstructs ADJ4 breastreconstruction) OR (reconstructs ADJ4 breastconservingsurgery) OR (reconstructs ADJ4 breastics) OR (reconstructs ADJ4 breast09) OR (reconstructs ADJ4 breastfed) OR (reconstructs ADJ4 breastq) OR (reconstructs ADJ4 breasttumor) OR (reconstructs ADJ4 breastbone) OR (reconstructs ADJ4 breastlike) OR (reconstruct ADJ4 breast) OR (reconstruct ADJ4 breasts) OR (reconstruct ADJ4 breastfeeding) OR (reconstruct ADJ4 breastmilk) OR (reconstruct ADJ4 breaststroke) OR (reconstruct ADJ4 breasting) OR (reconstruct ADJ4 breastcancer) OR (reconstruct ADJ4 breastsurgery) OR (reconstruct ADJ4 breasted) OR (reconstruct ADJ4 breastplate) OR (reconstruct ADJ4 breastchoice) OR (reconstruct ADJ4 breastfeed) OR (reconstruct ADJ4 breastreconstruction) OR (reconstruct ADJ4 breastconservingsurgery) OR (reconstruct ADJ4 breastics) OR (reconstruct ADJ4 breast09) OR (reconstruct ADJ4 breastfed) OR (reconstruct ADJ4 breastq) OR (reconstruct ADJ4 breasttumor) OR (reconstruct ADJ4 breastbone) OR (reconstruct ADJ4 breastlike) OR (reconstructable ADJ4 breast) OR (reconstructable ADJ4 breasts) OR (reconstructable ADJ4 breastfeeding) OR (reconstructable ADJ4 breastmilk) OR (reconstructable ADJ4 breaststroke) OR (reconstructable ADJ4 breasting) OR (reconstructable ADJ4 breastcancer) OR (reconstructable ADJ4 breastsurgery) OR (reconstructable ADJ4 breasted) OR (reconstructable ADJ4 breastplate) OR (reconstructable ADJ4 breastchoice) OR (reconstructable ADJ4 breastfeed) OR (reconstructable ADJ4 breastreconstruction) OR (reconstructable ADJ4 breastconservingsurgery) OR (reconstructable ADJ4 breastics) OR (reconstructable ADJ4 breast09) OR (reconstructable ADJ4 breastfed) OR (reconstructable ADJ4 breastq) OR (reconstructable ADJ4 breasttumor) OR (reconstructable ADJ4 breastbone) OR (reconstructable ADJ4 breastlike) OR (reconstructuion ADJ4 breast) OR (reconstructuion ADJ4 breasts) OR (reconstructuion ADJ4 breastfeeding) OR (reconstructuion ADJ4 breastmilk) OR (reconstructuion ADJ4 breaststroke) OR (reconstructuion ADJ4 breasting) OR (reconstructuion ADJ4 breastcancer) OR (reconstructuion ADJ4 breastsurgery) OR (reconstructuion ADJ4 breasted) OR (reconstructuion ADJ4 breastplate) OR (reconstructuion ADJ4 breastchoice) OR (reconstructuion ADJ4 breastfeed) OR (reconstructuion ADJ4 breastreconstruction) OR (reconstructuion ADJ4 breastconservingsurgery) OR (reconstructuion ADJ4 breastics) OR (reconstructuion ADJ4 breast09) OR (reconstructuion ADJ4 breastfed) OR (reconstructuion ADJ4 breastq) OR (reconstructuion ADJ4 breasttumor) OR (reconstructuion ADJ4 breastbone) OR (reconstructuion ADJ4 breastlike) OR (reconstructionwith ADJ4 breast) OR (reconstructionwith ADJ4 breasts) OR (reconstructionwith ADJ4 breastfeeding) OR (reconstructionwith ADJ4 breastmilk) OR (reconstructionwith ADJ4 breaststroke) OR (reconstructionwith ADJ4 breasting) OR (reconstructionwith ADJ4 breastcancer) OR (reconstructionwith ADJ4 breastsurgery) OR (reconstructionwith ADJ4 breasted) OR (reconstructionwith ADJ4 breastplate) OR (reconstructionwith ADJ4 breastchoice) OR (reconstructionwith ADJ4 breastfeed) OR (reconstructionwith ADJ4 breastreconstruction) OR (reconstructionwith ADJ4 breastconservingsurgery) OR (reconstructionwith ADJ4 breastics) OR (reconstructionwith ADJ4 breast09) OR (reconstructionwith ADJ4 breastfed) OR (reconstructionwith ADJ4 breastq) OR (reconstructionwith ADJ4 breasttumor) OR (reconstructionwith ADJ4 breastbone) OR (reconstructionwith ADJ4 breastlike) OR (augmentation ADJ4 breast) OR (augmentation ADJ4 breasts) OR (augmentation ADJ4 breastfeeding) OR (augmentation ADJ4 breastmilk) OR (augmentation ADJ4 breaststroke) OR (augmentation ADJ4 breasting) OR (augmentation ADJ4 breastcancer) OR (augmentation ADJ4 breastsurgery) OR (augmentation ADJ4 breasted) OR (augmentation ADJ4 breastplate) OR (augmentation ADJ4 breastchoice) OR (augmentation ADJ4 breastfeed) OR (augmentation ADJ4 breastreconstruction) OR (augmentation ADJ4 breastconservingsurgery) OR (augmentation ADJ4 breastics) OR (augmentation ADJ4 breast09) OR (augmentation ADJ4 breastfed) OR (augmentation ADJ4 breastq) OR (augmentation ADJ4 breasttumor) OR (augmentation ADJ4 breastbone) OR (augmentation ADJ4 breastlike) OR (augmented ADJ4 breast) OR (augmented ADJ4 breasts) OR (augmented ADJ4 breastfeeding) OR (augmented ADJ4 breastmilk) OR (augmented ADJ4 breaststroke) OR (augmented ADJ4 breasting) OR (augmented ADJ4 breastcancer) OR (augmented ADJ4 breastsurgery) OR (augmented ADJ4 breasted) OR (augmented ADJ4 breastplate) OR (augmented ADJ4 breastchoice) OR (augmented ADJ4 breastfeed) OR (augmented ADJ4 breastreconstruction) OR (augmented ADJ4 breastconservingsurgery) OR (augmented ADJ4 breastics) OR (augmented ADJ4 breast09) OR (augmented ADJ4 breastfed) OR (augmented ADJ4 breastq) OR (augmented ADJ4 breasttumor) OR (augmented ADJ4 breastbone) OR (augmented ADJ4 breastlike) OR (augmenting ADJ4 breast) OR (augmenting ADJ4 breasts) OR (augmenting ADJ4 breastfeeding) OR (augmenting ADJ4 breastmilk) OR (augmenting ADJ4 breaststroke) OR (augmenting ADJ4 breasting) OR (augmenting ADJ4 breastcancer) OR (augmenting ADJ4 breastsurgery) OR (augmenting ADJ4 breasted) OR (augmenting ADJ4 breastplate) OR (augmenting ADJ4 breastchoice) OR (augmenting ADJ4 breastfeed) OR (augmenting ADJ4 breastreconstruction) OR (augmenting ADJ4 breastconservingsurgery) OR (augmenting ADJ4 breastics) OR (augmenting ADJ4 breast09) OR (augmenting ADJ4 breastfed) OR (augmenting ADJ4 breastq) OR (augmenting ADJ4 breasttumor) OR (augmenting ADJ4 breastbone) OR (augmenting ADJ4 breastlike) OR (augment ADJ4 breast) OR (augment ADJ4 breasts) OR (augment ADJ4 breastfeeding) OR (augment ADJ4 breastmilk) OR (augment ADJ4 breaststroke) OR (augment ADJ4 breasting) OR (augment ADJ4 breastcancer) OR (augment ADJ4 breastsurgery) OR (augment ADJ4 breasted) OR (augment ADJ4 breastplate) OR (augment ADJ4 breastchoice) OR (augment ADJ4 breastfeed) OR (augment ADJ4 breastreconstruction) OR (augment ADJ4 breastconservingsurgery) OR (augment ADJ4 breastics) OR (augment ADJ4 breast09) OR (augment ADJ4 breastfed) OR (augment ADJ4 breastq) OR (augment ADJ4 breasttumor) OR (augment ADJ4 breastbone) OR (augment ADJ4 breastlike) OR (augments ADJ4 breast) OR (augments ADJ4 breasts) OR (augments ADJ4 breastfeeding) OR (augments ADJ4 breastmilk) OR (augments ADJ4 breaststroke) OR (augments ADJ4 breasting) OR (augments ADJ4 breastcancer) OR (augments ADJ4 breastsurgery) OR (augments ADJ4 breasted) OR (augments ADJ4 breastplate) OR (augments ADJ4 breastchoice) OR (augments ADJ4 breastfeed) OR (augments ADJ4 breastreconstruction) OR (augments ADJ4 breastconservingsurgery) OR (augments ADJ4 breastics) OR (augments ADJ4 breast09) OR (augments ADJ4 breastfed) OR (augments ADJ4 breastq) OR (augments ADJ4 breasttumor) OR (augments ADJ4 breastbone) OR (augments ADJ4 breastlike) OR (augmentin ADJ4 breast) OR (augmentin ADJ4 breasts) OR (augmentin ADJ4 breastfeeding) OR (augmentin ADJ4 breastmilk) OR (augmentin ADJ4 breaststroke) OR (augmentin ADJ4 breasting) OR (augmentin ADJ4 breastcancer) OR (augmentin ADJ4 breastsurgery) OR (augmentin ADJ4 breasted) OR (augmentin ADJ4 breastplate) OR (augmentin ADJ4 breastchoice) OR (augmentin ADJ4 breastfeed) OR (augmentin ADJ4 breastreconstruction) OR (augmentin ADJ4 breastconservingsurgery) OR (augmentin ADJ4 breastics) OR (augmentin ADJ4 breast09) OR (augmentin ADJ4 breastfed) OR (augmentin ADJ4 breastq) OR (augmentin ADJ4 breasttumor) OR (augmentin ADJ4 breastbone) OR (augmentin ADJ4 breastlike) OR (augmentations ADJ4 breast) OR (augmentations ADJ4 breasts) OR (augmentations ADJ4 breastfeeding) OR (augmentations ADJ4 breastmilk) OR (augmentations ADJ4 breaststroke) OR (augmentations ADJ4 breasting) OR (augmentations ADJ4 breastcancer) OR (augmentations ADJ4 breastsurgery) OR (augmentations ADJ4 breasted) OR (augmentations ADJ4 breastplate) OR (augmentations ADJ4 breastchoice) OR (augmentations ADJ4 breastfeed) OR (augmentations ADJ4 breastreconstruction) OR (augmentations ADJ4 breastconservingsurgery) OR (augmentations ADJ4 breastics) OR (augmentations ADJ4 breast09) OR (augmentations ADJ4 breastfed) OR (augmentations ADJ4 breastq) OR (augmentations ADJ4 breasttumor) OR (augmentations ADJ4 breastbone) OR (augmentations ADJ4 breastlike) OR (augmentative ADJ4 breast) OR (augmentative ADJ4 breasts) OR (augmentative ADJ4 breastfeeding) OR (augmentative ADJ4 breastmilk) OR (augmentative ADJ4 breaststroke) OR (augmentative ADJ4 breasting) OR (augmentative ADJ4 breastcancer) OR (augmentative ADJ4 breastsurgery) OR (augmentative ADJ4 breasted) OR (augmentative ADJ4 breastplate) OR (augmentative ADJ4 breastchoice) OR (augmentative ADJ4 breastfeed) OR (augmentative ADJ4 breastreconstruction) OR (augmentative ADJ4 breastconservingsurgery) OR (augmentative ADJ4 breastics) OR (augmentative ADJ4 breast09) OR (augmentative ADJ4 breastfed) OR (augmentative ADJ4 breastq) OR (augmentative ADJ4 breasttumor) OR (augmentative ADJ4 breastbone) OR (augmentative ADJ4 breastlike) OR (augmenter ADJ4 breast) OR (augmenter ADJ4 breasts) OR (augmenter ADJ4 breastfeeding) OR (augmenter ADJ4 breastmilk) OR (augmenter ADJ4 breaststroke) OR (augmenter ADJ4 breasting) OR (augmenter ADJ4 breastcancer) OR (augmenter ADJ4 breastsurgery) OR (augmenter ADJ4 breasted) OR (augmenter ADJ4 breastplate) OR (augmenter ADJ4 breastchoice) OR (augmenter ADJ4 breastfeed) OR (augmenter ADJ4 breastreconstruction) OR (augmenter ADJ4 breastconservingsurgery) OR (augmenter ADJ4 breastics) OR (augmenter ADJ4 breast09) OR (augmenter ADJ4 breastfed) OR (augmenter ADJ4 breastq) OR (augmenter ADJ4 breasttumor) OR (augmenter ADJ4 breastbone) OR (augmenter ADJ4 breastlike) OR (augmentors ADJ4 breast) OR (augmentors ADJ4 breasts) OR (augmentors ADJ4 breastfeeding) OR (augmentors ADJ4 breastmilk) OR (augmentors ADJ4 breaststroke) OR (augmentors ADJ4 breasting) OR (augmentors ADJ4 breastcancer) OR (augmentors ADJ4 breastsurgery) OR (augmentors ADJ4 breasted) OR (augmentors ADJ4 breastplate) OR (augmentors ADJ4 breastchoice) OR (augmentors ADJ4 breastfeed) OR (augmentors ADJ4 breastreconstruction) OR (augmentors ADJ4 breastconservingsurgery) OR (augmentors ADJ4 breastics) OR (augmentors ADJ4 breast09) OR (augmentors ADJ4 breastfed) OR (augmentors ADJ4 breastq) OR (augmentors ADJ4 breasttumor) OR (augmentors ADJ4 breastbone) OR (augmentors ADJ4 breastlike) OR (augmentor ADJ4 breast) OR (augmentor ADJ4 breasts) OR (augmentor ADJ4 breastfeeding) OR (augmentor ADJ4 breastmilk) OR (augmentor ADJ4 breaststroke) OR (augmentor ADJ4 breasting) OR (augmentor ADJ4 breastcancer) OR (augmentor ADJ4 breastsurgery) OR (augmentor ADJ4 breasted) OR (augmentor ADJ4 breastplate) OR (augmentor ADJ4 breastchoice) OR (augmentor ADJ4 breastfeed) OR (augmentor ADJ4 breastreconstruction) OR (augmentor ADJ4 breastconservingsurgery) OR (augmentor ADJ4 breastics) OR (augmentor ADJ4 breast09) OR (augmentor ADJ4 breastfed) OR (augmentor ADJ4 breastq) OR (augmentor ADJ4 breasttumor) OR (augmentor ADJ4 breastbone) OR (augmentor ADJ4 breastlike) OR (augmentatiom ADJ4 breast) OR (augmentatiom ADJ4 breasts) OR (augmentatiom ADJ4 breastfeeding) OR (augmentatiom ADJ4 breastmilk) OR (augmentatiom ADJ4 breaststroke) OR (augmentatiom ADJ4 breasting) OR (augmentatiom ADJ4 breastcancer) OR (augmentatiom ADJ4 breastsurgery) OR (augmentatiom ADJ4 breasted) OR (augmentatiom ADJ4 breastplate) OR (augmentatiom ADJ4 breastchoice) OR (augmentatiom ADJ4 breastfeed) OR (augmentatiom ADJ4 breastreconstruction) OR (augmentatiom ADJ4 breastconservingsurgery) OR (augmentatiom ADJ4 breastics) OR (augmentatiom ADJ4 breast09) OR (augmentatiom ADJ4 breastfed) OR (augmentatiom ADJ4 breastq) OR (augmentatiom ADJ4 breasttumor) OR (augmentatiom ADJ4 breastbone) OR (augmentatiom ADJ4 breastlike) OR (augmentive ADJ4 breast) OR (augmentive ADJ4 breasts) OR (augmentive ADJ4 breastfeeding) OR (augmentive ADJ4 breastmilk) OR (augmentive ADJ4 breaststroke) OR (augmentive ADJ4 breasting) OR (augmentive ADJ4 breastcancer) OR (augmentive ADJ4 breastsurgery) OR (augmentive ADJ4 breasted) OR (augmentive ADJ4 breastplate) OR (augmentive ADJ4 breastchoice) OR (augmentive ADJ4 breastfeed) OR (augmentive ADJ4 breastreconstruction) OR (augmentive ADJ4 breastconservingsurgery) OR (augmentive ADJ4 breastics) OR (augmentive ADJ4 breast09) OR (augmentive ADJ4 breastfed) OR (augmentive ADJ4 breastq) OR (augmentive ADJ4 breasttumor) OR (augmentive ADJ4 breastbone) OR (augmentive ADJ4 breastlike) OR (oncoplastic ADJ4 breast) OR (oncoplastic ADJ4 breasts) OR (oncoplastic ADJ4 breastfeeding) OR (oncoplastic ADJ4 breastmilk) OR (oncoplastic ADJ4 breaststroke) OR (oncoplastic ADJ4 breasting) OR (oncoplastic ADJ4 breastcancer) OR (oncoplastic ADJ4 breastsurgery) OR (oncoplastic ADJ4 breasted) OR (oncoplastic ADJ4 breastplate) OR (oncoplastic ADJ4 breastchoice) OR (oncoplastic ADJ4 breastfeed) OR (oncoplastic ADJ4 breastreconstruction) OR (oncoplastic ADJ4 breastconservingsurgery) OR (oncoplastic ADJ4 breastics) OR (oncoplastic ADJ4 breast09) OR (oncoplastic ADJ4 breastfed) OR (oncoplastic ADJ4 breastq) OR (oncoplastic ADJ4 breasttumor) OR (oncoplastic ADJ4 breastbone) OR (oncoplastic ADJ4 breastlike) OR (oncoplasticsurgery ADJ4 breast) OR (oncoplasticsurgery ADJ4 breasts) OR (oncoplasticsurgery ADJ4 breastfeeding) OR (oncoplasticsurgery ADJ4 breastmilk) OR (oncoplasticsurgery ADJ4 breaststroke) OR (oncoplasticsurgery ADJ4 breasting) OR (oncoplasticsurgery ADJ4 breastcancer) OR (oncoplasticsurgery ADJ4 breastsurgery) OR (oncoplasticsurgery ADJ4 breasted) OR (oncoplasticsurgery ADJ4 breastplate) OR (oncoplasticsurgery ADJ4 breastchoice) OR (oncoplasticsurgery ADJ4 breastfeed) OR (oncoplasticsurgery ADJ4 breastreconstruction) OR (oncoplasticsurgery ADJ4 breastconservingsurgery) OR (oncoplasticsurgery ADJ4 breastics) OR (oncoplasticsurgery ADJ4 breast09) OR (oncoplasticsurgery ADJ4 breastfed) OR (oncoplasticsurgery ADJ4 breastq) OR (oncoplasticsurgery ADJ4 breasttumor) OR (oncoplasticsurgery ADJ4 breastbone) OR (oncoplasticsurgery ADJ4 breastlike) OR (oncoplastique ADJ4 breast) OR (oncoplastique ADJ4 breasts) OR (oncoplastique ADJ4 breastfeeding) OR (oncoplastique ADJ4 breastmilk) OR (oncoplastique ADJ4 breaststroke) OR (oncoplastique ADJ4 breasting) OR (oncoplastique ADJ4 breastcancer) OR (oncoplastique ADJ4 breastsurgery) OR (oncoplastique ADJ4 breasted) OR (oncoplastique ADJ4 breastplate) OR (oncoplastique ADJ4 breastchoice) OR (oncoplastique ADJ4 breastfeed) OR (oncoplastique ADJ4 breastreconstruction) OR (oncoplastique ADJ4 breastconservingsurgery) OR (oncoplastique ADJ4 breastics) OR (oncoplastique ADJ4 breast09) OR (oncoplastique ADJ4 breastfed) OR (oncoplastique ADJ4 breastq) OR (oncoplastique ADJ4 breasttumor) OR (oncoplastique ADJ4 breastbone) OR (oncoplastique ADJ4 breastlike) OR (oncoplastie ADJ4 breast) OR (oncoplastie ADJ4 breasts) OR (oncoplastie ADJ4 breastfeeding) OR (oncoplastie ADJ4 breastmilk) OR (oncoplastie ADJ4 breaststroke) OR (oncoplastie ADJ4 breasting) OR (oncoplastie ADJ4 breastcancer) OR (oncoplastie ADJ4 breastsurgery) OR (oncoplastie ADJ4 breasted) OR (oncoplastie ADJ4 breastplate) OR (oncoplastie ADJ4 breastchoice) OR (oncoplastie ADJ4 breastfeed) OR (oncoplastie ADJ4 breastreconstruction) OR (oncoplastie ADJ4 breastconservingsurgery) OR (oncoplastie ADJ4 breastics) OR (oncoplastie ADJ4 breast09) OR (oncoplastie ADJ4 breastfed) OR (oncoplastie ADJ4 breastq) OR (oncoplastie ADJ4 breasttumor) OR (oncoplastie ADJ4 breastbone) OR (oncoplastie ADJ4 breastlike) OR (oncoplastics ADJ4 breast) OR (oncoplastics ADJ4 breasts) OR (oncoplastics ADJ4 breastfeeding) OR (oncoplastics ADJ4 breastmilk) OR (oncoplastics ADJ4 breaststroke) OR (oncoplastics ADJ4 breasting) OR (oncoplastics ADJ4 breastcancer) OR (oncoplastics ADJ4 breastsurgery) OR (oncoplastics ADJ4 breasted) OR (oncoplastics ADJ4 breastplate) OR (oncoplastics ADJ4 breastchoice) OR (oncoplastics ADJ4 breastfeed) OR (oncoplastics ADJ4 breastreconstruction) OR (oncoplastics ADJ4 breastconservingsurgery) OR (oncoplastics ADJ4 breastics) OR (oncoplastics ADJ4 breast09) OR (oncoplastics ADJ4 breastfed) OR (oncoplastics ADJ4 breastq) OR (oncoplastics ADJ4 breasttumor) OR (oncoplastics ADJ4 breastbone) OR (oncoplastics ADJ4 breastlike) OR (oncoplasties ADJ4 breast) OR (oncoplasties ADJ4 breasts) OR (oncoplasties ADJ4 breastfeeding) OR (oncoplasties ADJ4 breastmilk) OR (oncoplasties ADJ4 breaststroke) OR (oncoplasties ADJ4 breasting) OR (oncoplasties ADJ4 breastcancer) OR (oncoplasties ADJ4 breastsurgery) OR (oncoplasties ADJ4 breasted) OR (oncoplasties ADJ4 breastplate) OR (oncoplasties ADJ4 breastchoice) OR (oncoplasties ADJ4 breastfeed) OR (oncoplasties ADJ4 breastreconstruction) OR (oncoplasties ADJ4 breastconservingsurgery) OR (oncoplasties ADJ4 breastics) OR (oncoplasties ADJ4 breast09) OR (oncoplasties ADJ4 breastfed) OR (oncoplasties ADJ4 breastq) OR (oncoplasties ADJ4 breasttumor) OR (oncoplasties ADJ4 breastbone) OR (oncoplasties ADJ4 breastlike)) or mammoplast* or mastectom* or mastopex*).ti,ab,kf. or (exp "Transplant Donor Site"/ or ("donor site*" or "harvest site*").ti,ab,kf.))

Selected terms (it was opted to list selected terms rather than rejected, as there were over 1000 terms to choose from at trans*):
lipo: lipo, lipoid, lipoaspirate, lipofilling, lipoplasty, lipotransfer, lipomodeling, lipoinjection, lipoaspiration
lipid: lipid, lipids, lipidic, lipidemic
trans: transport, transplantation, transfer, transmission, transporter, transplant, trans, translocation, translocator, transfusions, transfusion, transferred, transfused, translocates, transfat, transbuccal, transposition, transportation, transferable, transpose, transported, transplanted, translocalization, transporting, transfered, transposed, transplantable, transplantations, transfusional, transfats, transports, transposable, transfers, transportable, transplantology, transplantability, translocating, transplanting, transplants, transferrable, transfusing, transferral, transportability, translocated, transposing, translocations,
autotrans: autotransplantation, autotransporter, autotransplant, autotransfusion, autotransporters, autotransport, autotransplants, autotransplanting, autotransplanted,

**Included articles Ids (7/7):** 30533985, 27155193, 24604787, 26962306, 39285055, 32221675, 39950221
Retrieved by original search: 6/7
Retreived by remi2.1 search: 6/7

#### Search 4 (Peters et al.): <https://pubmed.ncbi.nlm.nih.gov/42531669/>

##### Search 4: Original MEDLINE Search: (8116 hits)

##### (exp "Chronic Disease"/ or ((chronic* OR long-term OR ongoing OR continuous OR non communicable OR noncommunicable OR persistent OR enduring) ADJ3 (disease* OR condition* OR illness* OR disorder* OR sick*)).ti,ab,kf.) AND (exp "Patient participation"/ or exp "Self-management"/ or (("Self manag*" or "Self care" or “Person centered care” or ("patient*" adj3 Empower*)).ti,ab,kf. or ("Patient" adj3 (Activati* or Engag* or Involv* or Participati* or Contributi* or Collaborat*)).ti,kf.)) AND (exp "cardiovascular diseases"/ or exp "Osteoarthritis"/ or exp "Respiration disorders"/ or exp "respiratory tract diseases"/ or exp "diabetes mellitus"/ or ("cardiovascular*" or "cardiac*" or "heart" or "osteoarth*" or "respirat*” or “pulmon*" or "lung*" or "breathing" or "diabet*" OR DM OR T2DM OR T1DM).ti,ab,kf.)

##### Search 4: REMI 2 Proximity Search convert to Medline (8114 hits):

(exp "Chronic Disease"/ or (((chronic ADJ3 disease) OR (chronic ADJ3 diseases) OR (chronic ADJ3 diseased) OR (chronic ADJ3 diseasefree) OR (chronic ADJ3 disease0) OR (chronic ADJ3 diseasewas) OR (chronic ADJ3 diseasein) OR (chronic ADJ3 diseaserenin) OR (chronic ADJ3 diseaserelated) OR (chronic ADJ3 diseasemodifying) OR (chronic ADJ3 diseasemeth) OR (chronic ADJ3 diseaseavancando) OR (chronic ADJ3 diseaseprevention) OR (chronic ADJ3 disease19) OR (chronic ADJ3 diseases1) OR (chronic ADJ3 diseasehepatitis) OR (chronic ADJ3 diseaseses) OR (chronic ADJ3 diseasebefore) OR (chronic ADJ3 diseasesthe) OR (chronic ADJ3 diseasesincluding) OR (chronic ADJ3 diseasest) OR (chronic ADJ3 diseaseprogression) OR (chronic ADJ3 diseaseci) OR (chronic ADJ3 diseasedjoint) OR (chronic ADJ3 diseaseand) OR (chronic ADJ3 diseasewith) OR (chronic ADJ3 diseaseful) OR (chronic ADJ3 diseasemanagement) OR (chronic ADJ3 diseasefor) OR (chronic ADJ3 diseasepatients) OR (chronic ADJ3 diseasethat) OR (chronic ADJ3 conditions) OR (chronic ADJ3 conditionally) OR (chronic ADJ3 condition) OR (chronic ADJ3 conditioner) OR (chronic ADJ3 conditioners) OR (chronic ADJ3 conditional) OR (chronic ADJ3 conditioning) OR (chronic ADJ3 conditionality) OR (chronic ADJ3 conditioned) OR (chronic ADJ3 conditionings) OR (chronic ADJ3 conditionalities) OR (chronic ADJ3 conditionals) OR (chronic ADJ3 conditionallyhealthy) OR (chronic ADJ3 conditionalization) OR (chronic ADJ3 conditionstrial) OR (chronic ADJ3 conditionsin) OR (chronic ADJ3 conditionsns) OR (chronic ADJ3 conditionsby) OR (chronic ADJ3 conditionning) OR (chronic ADJ3 conditioningcoaches) OR (chronic ADJ3 conditionining) OR (chronic ADJ3 conditionsthe) OR (chronic ADJ3 conditioncategory) OR (chronic ADJ3 conditionsglass) OR (chronic ADJ3 conditionized) OR (chronic ADJ3 conditionstrained) OR (chronic ADJ3 conditions4) OR (chronic ADJ3 conditionsto) OR (chronic ADJ3 conditionsfor) OR (chronic ADJ3 conditionn) OR (chronic ADJ3 conditionary) OR (chronic ADJ3 conditionability) OR (chronic ADJ3 conditiong) OR (chronic ADJ3 illness) OR (chronic ADJ3 illnesses) OR (chronic ADJ3 illnesspmh) OR (chronic ADJ3 illnessnwb) OR (chronic ADJ3 illnessnew) OR (chronic ADJ3 disorder) OR (chronic ADJ3 disorders) OR (chronic ADJ3 disorderly) OR (chronic ADJ3 disordering) OR (chronic ADJ3 disordered) OR (chronic ADJ3 disorderliness) OR (chronic ADJ3 disorderswhich) OR (chronic ADJ3 disorderof) OR (chronic ADJ3 disorderd) OR (chronic ADJ3 disordercould) OR (chronic ADJ3 disorderis) OR (chronic ADJ3 disorderrs) OR (chronic ADJ3 sicker) OR (chronic ADJ3 sickest) OR (chronic ADJ3 sickle) OR (chronic ADJ3 sickkids) OR (chronic ADJ3 sicknesses) OR (chronic ADJ3 sickness) OR (chronic ADJ3 sickening) OR (chronic ADJ3 sicklefin) OR (chronic ADJ3 sickened) OR (chronic ADJ3 sickling) OR (chronic ADJ3 sickbed) OR (chronic ADJ3 sick) OR (chronic ADJ3 sickly) OR (chronic ADJ3 sicko) OR (chronic ADJ3 sickles) OR (chronic ADJ3 sickel) OR (chronic ADJ3 sickroom) OR (chronic ADJ3 sicks) OR (chronic ADJ3 sickbay) OR (chronic ADJ3 sicktime) OR (chronic ADJ3 sickie) OR (chronic ADJ3 sicknes) OR (chronic ADJ3 sickleave) OR (chronic ADJ3 sickens) OR (chronic ADJ3 sicklaneny) OR (chronic ADJ3 sicklepod) OR (chronic ADJ3 sickliness) OR (chronic ADJ3 sickenss) OR (chronic ADJ3 sicklemia) OR (chronically ADJ3 disease) OR (chronically ADJ3 diseases) OR (chronically ADJ3 diseased) OR (chronically ADJ3 diseasefree) OR (chronically ADJ3 disease0) OR (chronically ADJ3 diseasewas) OR (chronically ADJ3 diseasein) OR (chronically ADJ3 diseaserenin) OR (chronically ADJ3 diseaserelated) OR (chronically ADJ3 diseasemodifying) OR (chronically ADJ3 diseasemeth) OR (chronically ADJ3 diseaseavancando) OR (chronically ADJ3 diseaseprevention) OR (chronically ADJ3 disease19) OR (chronically ADJ3 diseases1) OR (chronically ADJ3 diseasehepatitis) OR (chronically ADJ3 diseaseses) OR (chronically ADJ3 diseasebefore) OR (chronically ADJ3 diseasesthe) OR (chronically ADJ3 diseasesincluding) OR (chronically ADJ3 diseasest) OR (chronically ADJ3 diseaseprogression) OR (chronically ADJ3 diseaseci) OR (chronically ADJ3 diseasedjoint) OR (chronically ADJ3 diseaseand) OR (chronically ADJ3 diseasewith) OR (chronically ADJ3 diseaseful) OR (chronically ADJ3 diseasemanagement) OR (chronically ADJ3 diseasefor) OR (chronically ADJ3 diseasepatients) OR (chronically ADJ3 diseasethat) OR (chronically ADJ3 conditions) OR (chronically ADJ3 conditionally) OR (chronically ADJ3 condition) OR (chronically ADJ3 conditioner) OR (chronically ADJ3 conditioners) OR (chronically ADJ3 conditional) OR (chronically ADJ3 conditioning) OR (chronically ADJ3 conditionality) OR (chronically ADJ3 conditioned) OR (chronically ADJ3 conditionings) OR (chronically ADJ3 conditionalities) OR (chronically ADJ3 conditionals) OR (chronically ADJ3 conditionallyhealthy) OR (chronically ADJ3 conditionalization) OR (chronically ADJ3 conditionstrial) OR (chronically ADJ3 conditionsin) OR (chronically ADJ3 conditionsns) OR (chronically ADJ3 conditionsby) OR (chronically ADJ3 conditionning) OR (chronically ADJ3 conditioningcoaches) OR (chronically ADJ3 conditionining) OR (chronically ADJ3 conditionsthe) OR (chronically ADJ3 conditioncategory) OR (chronically ADJ3 conditionsglass) OR (chronically ADJ3 conditionized) OR (chronically ADJ3 conditionstrained) OR (chronically ADJ3 conditions4) OR (chronically ADJ3 conditionsto) OR (chronically ADJ3 conditionsfor) OR (chronically ADJ3 conditionn) OR (chronically ADJ3 conditionary) OR (chronically ADJ3 conditionability) OR (chronically ADJ3 conditiong) OR (chronically ADJ3 illness) OR (chronically ADJ3 illnesses) OR (chronically ADJ3 illnesspmh) OR (chronically ADJ3 illnessnwb) OR (chronically ADJ3 illnessnew) OR (chronically ADJ3 disorder) OR (chronically ADJ3 disorders) OR (chronically ADJ3 disorderly) OR (chronically ADJ3 disordering) OR (chronically ADJ3 disordered) OR (chronically ADJ3 disorderliness) OR (chronically ADJ3 disorderswhich) OR (chronically ADJ3 disorderof) OR (chronically ADJ3 disorderd) OR (chronically ADJ3 disordercould) OR (chronically ADJ3 disorderis) OR (chronically ADJ3 disorderrs) OR (chronically ADJ3 sicker) OR (chronically ADJ3 sickest) OR (chronically ADJ3 sickle) OR (chronically ADJ3 sickkids) OR (chronically ADJ3 sicknesses) OR (chronically ADJ3 sickness) OR (chronically ADJ3 sickening) OR (chronically ADJ3 sicklefin) OR (chronically ADJ3 sickened) OR (chronically ADJ3 sickling) OR (chronically ADJ3 sickbed) OR (chronically ADJ3 sick) OR (chronically ADJ3 sickly) OR (chronically ADJ3 sicko) OR (chronically ADJ3 sickles) OR (chronically ADJ3 sickel) OR (chronically ADJ3 sickroom) OR (chronically ADJ3 sicks) OR (chronically ADJ3 sickbay) OR (chronically ADJ3 sicktime) OR (chronically ADJ3 sickie) OR (chronically ADJ3 sicknes) OR (chronically ADJ3 sickleave) OR (chronically ADJ3 sickens) OR (chronically ADJ3 sicklaneny) OR (chronically ADJ3 sicklepod) OR (chronically ADJ3 sickliness) OR (chronically ADJ3 sickenss) OR (chronically ADJ3 sicklemia) OR (chronicity ADJ3 disease) OR (chronicity ADJ3 diseases) OR (chronicity ADJ3 diseased) OR (chronicity ADJ3 diseasefree) OR (chronicity ADJ3 disease0) OR (chronicity ADJ3 diseasewas) OR (chronicity ADJ3 diseasein) OR (chronicity ADJ3 diseaserenin) OR (chronicity ADJ3 diseaserelated) OR (chronicity ADJ3 diseasemodifying) OR (chronicity ADJ3 diseasemeth) OR (chronicity ADJ3 diseaseavancando) OR (chronicity ADJ3 diseaseprevention) OR (chronicity ADJ3 disease19) OR (chronicity ADJ3 diseases1) OR (chronicity ADJ3 diseasehepatitis) OR (chronicity ADJ3 diseaseses) OR (chronicity ADJ3 diseasebefore) OR (chronicity ADJ3 diseasesthe) OR (chronicity ADJ3 diseasesincluding) OR (chronicity ADJ3 diseasest) OR (chronicity ADJ3 diseaseprogression) OR (chronicity ADJ3 diseaseci) OR (chronicity ADJ3 diseasedjoint) OR (chronicity ADJ3 diseaseand) OR (chronicity ADJ3 diseasewith) OR (chronicity ADJ3 diseaseful) OR (chronicity ADJ3 diseasemanagement) OR (chronicity ADJ3 diseasefor) OR (chronicity ADJ3 diseasepatients) OR (chronicity ADJ3 diseasethat) OR (chronicity ADJ3 conditions) OR (chronicity ADJ3 conditionally) OR (chronicity ADJ3 condition) OR (chronicity ADJ3 conditioner) OR (chronicity ADJ3 conditioners) OR (chronicity ADJ3 conditional) OR (chronicity ADJ3 conditioning) OR (chronicity ADJ3 conditionality) OR (chronicity ADJ3 conditioned) OR (chronicity ADJ3 conditionings) OR (chronicity ADJ3 conditionalities) OR (chronicity ADJ3 conditionals) OR (chronicity ADJ3 conditionallyhealthy) OR (chronicity ADJ3 conditionalization) OR (chronicity ADJ3 conditionstrial) OR (chronicity ADJ3 conditionsin) OR (chronicity ADJ3 conditionsns) OR (chronicity ADJ3 conditionsby) OR (chronicity ADJ3 conditionning) OR (chronicity ADJ3 conditioningcoaches) OR (chronicity ADJ3 conditionining) OR (chronicity ADJ3 conditionsthe) OR (chronicity ADJ3 conditioncategory) OR (chronicity ADJ3 conditionsglass) OR (chronicity ADJ3 conditionized) OR (chronicity ADJ3 conditionstrained) OR (chronicity ADJ3 conditions4) OR (chronicity ADJ3 conditionsto) OR (chronicity ADJ3 conditionsfor) OR (chronicity ADJ3 conditionn) OR (chronicity ADJ3 conditionary) OR (chronicity ADJ3 conditionability) OR (chronicity ADJ3 conditiong) OR (chronicity ADJ3 illness) OR (chronicity ADJ3 illnesses) OR (chronicity ADJ3 illnesspmh) OR (chronicity ADJ3 illnessnwb) OR (chronicity ADJ3 illnessnew) OR (chronicity ADJ3 disorder) OR (chronicity ADJ3 disorders) OR (chronicity ADJ3 disorderly) OR (chronicity ADJ3 disordering) OR (chronicity ADJ3 disordered) OR (chronicity ADJ3 disorderliness) OR (chronicity ADJ3 disorderswhich) OR (chronicity ADJ3 disorderof) OR (chronicity ADJ3 disorderd) OR (chronicity ADJ3 disordercould) OR (chronicity ADJ3 disorderis) OR (chronicity ADJ3 disorderrs) OR (chronicity ADJ3 sicker) OR (chronicity ADJ3 sickest) OR (chronicity ADJ3 sickle) OR (chronicity ADJ3 sickkids) OR (chronicity ADJ3 sicknesses) OR (chronicity ADJ3 sickness) OR (chronicity ADJ3 sickening) OR (chronicity ADJ3 sicklefin) OR (chronicity ADJ3 sickened) OR (chronicity ADJ3 sickling) OR (chronicity ADJ3 sickbed) OR (chronicity ADJ3 sick) OR (chronicity ADJ3 sickly) OR (chronicity ADJ3 sicko) OR (chronicity ADJ3 sickles) OR (chronicity ADJ3 sickel) OR (chronicity ADJ3 sickroom) OR (chronicity ADJ3 sicks) OR (chronicity ADJ3 sickbay) OR (chronicity ADJ3 sicktime) OR (chronicity ADJ3 sickie) OR (chronicity ADJ3 sicknes) OR (chronicity ADJ3 sickleave) OR (chronicity ADJ3 sickens) OR (chronicity ADJ3 sicklaneny) OR (chronicity ADJ3 sicklepod) OR (chronicity ADJ3 sickliness) OR (chronicity ADJ3 sickenss) OR (chronicity ADJ3 sicklemia) OR (chronicpulmonary ADJ3 disease) OR (chronicpulmonary ADJ3 diseases) OR (chronicpulmonary ADJ3 diseased) OR (chronicpulmonary ADJ3 diseasefree) OR (chronicpulmonary ADJ3 disease0) OR (chronicpulmonary ADJ3 diseasewas) OR (chronicpulmonary ADJ3 diseasein) OR (chronicpulmonary ADJ3 diseaserenin) OR (chronicpulmonary ADJ3 diseaserelated) OR (chronicpulmonary ADJ3 diseasemodifying) OR (chronicpulmonary ADJ3 diseasemeth) OR (chronicpulmonary ADJ3 diseaseavancando) OR (chronicpulmonary ADJ3 diseaseprevention) OR (chronicpulmonary ADJ3 disease19) OR (chronicpulmonary ADJ3 diseases1) OR (chronicpulmonary ADJ3 diseasehepatitis) OR (chronicpulmonary ADJ3 diseaseses) OR (chronicpulmonary ADJ3 diseasebefore) OR (chronicpulmonary ADJ3 diseasesthe) OR (chronicpulmonary ADJ3 diseasesincluding) OR (chronicpulmonary ADJ3 diseasest) OR (chronicpulmonary ADJ3 diseaseprogression) OR (chronicpulmonary ADJ3 diseaseci) OR (chronicpulmonary ADJ3 diseasedjoint) OR (chronicpulmonary ADJ3 diseaseand) OR (chronicpulmonary ADJ3 diseasewith) OR (chronicpulmonary ADJ3 diseaseful) OR (chronicpulmonary ADJ3 diseasemanagement) OR (chronicpulmonary ADJ3 diseasefor) OR (chronicpulmonary ADJ3 diseasepatients) OR (chronicpulmonary ADJ3 diseasethat) OR (chronicpulmonary ADJ3 conditions) OR (chronicpulmonary ADJ3 conditionally) OR (chronicpulmonary ADJ3 condition) OR (chronicpulmonary ADJ3 conditioner) OR (chronicpulmonary ADJ3 conditioners) OR (chronicpulmonary ADJ3 conditional) OR (chronicpulmonary ADJ3 conditioning) OR (chronicpulmonary ADJ3 conditionality) OR (chronicpulmonary ADJ3 conditioned) OR (chronicpulmonary ADJ3 conditionings) OR (chronicpulmonary ADJ3 conditionalities) OR (chronicpulmonary ADJ3 conditionals) OR (chronicpulmonary ADJ3 conditionallyhealthy) OR (chronicpulmonary ADJ3 conditionalization) OR (chronicpulmonary ADJ3 conditionstrial) OR (chronicpulmonary ADJ3 conditionsin) OR (chronicpulmonary ADJ3 conditionsns) OR (chronicpulmonary ADJ3 conditionsby) OR (chronicpulmonary ADJ3 conditionning) OR (chronicpulmonary ADJ3 conditioningcoaches) OR (chronicpulmonary ADJ3 conditionining) OR (chronicpulmonary ADJ3 conditionsthe) OR (chronicpulmonary ADJ3 conditioncategory) OR (chronicpulmonary ADJ3 conditionsglass) OR (chronicpulmonary ADJ3 conditionized) OR (chronicpulmonary ADJ3 conditionstrained) OR (chronicpulmonary ADJ3 conditions4) OR (chronicpulmonary ADJ3 conditionsto) OR (chronicpulmonary ADJ3 conditionsfor) OR (chronicpulmonary ADJ3 conditionn) OR (chronicpulmonary ADJ3 conditionary) OR (chronicpulmonary ADJ3 conditionability) OR (chronicpulmonary ADJ3 conditiong) OR (chronicpulmonary ADJ3 illness) OR (chronicpulmonary ADJ3 illnesses) OR (chronicpulmonary ADJ3 illnesspmh) OR (chronicpulmonary ADJ3 illnessnwb) OR (chronicpulmonary ADJ3 illnessnew) OR (chronicpulmonary ADJ3 disorder) OR (chronicpulmonary ADJ3 disorders) OR (chronicpulmonary ADJ3 disorderly) OR (chronicpulmonary ADJ3 disordering) OR (chronicpulmonary ADJ3 disordered) OR (chronicpulmonary ADJ3 disorderliness) OR (chronicpulmonary ADJ3 disorderswhich) OR (chronicpulmonary ADJ3 disorderof) OR (chronicpulmonary ADJ3 disorderd) OR (chronicpulmonary ADJ3 disordercould) OR (chronicpulmonary ADJ3 disorderis) OR (chronicpulmonary ADJ3 disorderrs) OR (chronicpulmonary ADJ3 sicker) OR (chronicpulmonary ADJ3 sickest) OR (chronicpulmonary ADJ3 sickle) OR (chronicpulmonary ADJ3 sickkids) OR (chronicpulmonary ADJ3 sicknesses) OR (chronicpulmonary ADJ3 sickness) OR (chronicpulmonary ADJ3 sickening) OR (chronicpulmonary ADJ3 sicklefin) OR (chronicpulmonary ADJ3 sickened) OR (chronicpulmonary ADJ3 sickling) OR (chronicpulmonary ADJ3 sickbed) OR (chronicpulmonary ADJ3 sick) OR (chronicpulmonary ADJ3 sickly) OR (chronicpulmonary ADJ3 sicko) OR (chronicpulmonary ADJ3 sickles) OR (chronicpulmonary ADJ3 sickel) OR (chronicpulmonary ADJ3 sickroom) OR (chronicpulmonary ADJ3 sicks) OR (chronicpulmonary ADJ3 sickbay) OR (chronicpulmonary ADJ3 sicktime) OR (chronicpulmonary ADJ3 sickie) OR (chronicpulmonary ADJ3 sicknes) OR (chronicpulmonary ADJ3 sickleave) OR (chronicpulmonary ADJ3 sickens) OR (chronicpulmonary ADJ3 sicklaneny) OR (chronicpulmonary ADJ3 sicklepod) OR (chronicpulmonary ADJ3 sickliness) OR (chronicpulmonary ADJ3 sickenss) OR (chronicpulmonary ADJ3 sicklemia) OR (chroniclymphocytic ADJ3 disease) OR (chroniclymphocytic ADJ3 diseases) OR (chroniclymphocytic ADJ3 diseased) OR (chroniclymphocytic ADJ3 diseasefree) OR (chroniclymphocytic ADJ3 disease0) OR (chroniclymphocytic ADJ3 diseasewas) OR (chroniclymphocytic ADJ3 diseasein) OR (chroniclymphocytic ADJ3 diseaserenin) OR (chroniclymphocytic ADJ3 diseaserelated) OR (chroniclymphocytic ADJ3 diseasemodifying) OR (chroniclymphocytic ADJ3 diseasemeth) OR (chroniclymphocytic ADJ3 diseaseavancando) OR (chroniclymphocytic ADJ3 diseaseprevention) OR (chroniclymphocytic ADJ3 disease19) OR (chroniclymphocytic ADJ3 diseases1) OR (chroniclymphocytic ADJ3 diseasehepatitis) OR (chroniclymphocytic ADJ3 diseaseses) OR (chroniclymphocytic ADJ3 diseasebefore) OR (chroniclymphocytic ADJ3 diseasesthe) OR (chroniclymphocytic ADJ3 diseasesincluding) OR (chroniclymphocytic ADJ3 diseasest) OR (chroniclymphocytic ADJ3 diseaseprogression) OR (chroniclymphocytic ADJ3 diseaseci) OR (chroniclymphocytic ADJ3 diseasedjoint) OR (chroniclymphocytic ADJ3 diseaseand) OR (chroniclymphocytic ADJ3 diseasewith) OR (chroniclymphocytic ADJ3 diseaseful) OR (chroniclymphocytic ADJ3 diseasemanagement) OR (chroniclymphocytic ADJ3 diseasefor) OR (chroniclymphocytic ADJ3 diseasepatients) OR (chroniclymphocytic ADJ3 diseasethat) OR (chroniclymphocytic ADJ3 conditions) OR (chroniclymphocytic ADJ3 conditionally) OR (chroniclymphocytic ADJ3 condition) OR (chroniclymphocytic ADJ3 conditioner) OR (chroniclymphocytic ADJ3 conditioners) OR (chroniclymphocytic ADJ3 conditional) OR (chroniclymphocytic ADJ3 conditioning) OR (chroniclymphocytic ADJ3 conditionality) OR (chroniclymphocytic ADJ3 conditioned) OR (chroniclymphocytic ADJ3 conditionings) OR (chroniclymphocytic ADJ3 conditionalities) OR (chroniclymphocytic ADJ3 conditionals) OR (chroniclymphocytic ADJ3 conditionallyhealthy) OR (chroniclymphocytic ADJ3 conditionalization) OR (chroniclymphocytic ADJ3 conditionstrial) OR (chroniclymphocytic ADJ3 conditionsin) OR (chroniclymphocytic ADJ3 conditionsns) OR (chroniclymphocytic ADJ3 conditionsby) OR (chroniclymphocytic ADJ3 conditionning) OR (chroniclymphocytic ADJ3 conditioningcoaches) OR (chroniclymphocytic ADJ3 conditionining) OR (chroniclymphocytic ADJ3 conditionsthe) OR (chroniclymphocytic ADJ3 conditioncategory) OR (chroniclymphocytic ADJ3 conditionsglass) OR (chroniclymphocytic ADJ3 conditionized) OR (chroniclymphocytic ADJ3 conditionstrained) OR (chroniclymphocytic ADJ3 conditions4) OR (chroniclymphocytic ADJ3 conditionsto) OR (chroniclymphocytic ADJ3 conditionsfor) OR (chroniclymphocytic ADJ3 conditionn) OR (chroniclymphocytic ADJ3 conditionary) OR (chroniclymphocytic ADJ3 conditionability) OR (chroniclymphocytic ADJ3 conditiong) OR (chroniclymphocytic ADJ3 illness) OR (chroniclymphocytic ADJ3 illnesses) OR (chroniclymphocytic ADJ3 illnesspmh) OR (chroniclymphocytic ADJ3 illnessnwb) OR (chroniclymphocytic ADJ3 illnessnew) OR (chroniclymphocytic ADJ3 disorder) OR (chroniclymphocytic ADJ3 disorders) OR (chroniclymphocytic ADJ3 disorderly) OR (chroniclymphocytic ADJ3 disordering) OR (chroniclymphocytic ADJ3 disordered) OR (chroniclymphocytic ADJ3 disorderliness) OR (chroniclymphocytic ADJ3 disorderswhich) OR (chroniclymphocytic ADJ3 disorderof) OR (chroniclymphocytic ADJ3 disorderd) OR (chroniclymphocytic ADJ3 disordercould) OR (chroniclymphocytic ADJ3 disorderis) OR (chroniclymphocytic ADJ3 disorderrs) OR (chroniclymphocytic ADJ3 sicker) OR (chroniclymphocytic ADJ3 sickest) OR (chroniclymphocytic ADJ3 sickle) OR (chroniclymphocytic ADJ3 sickkids) OR (chroniclymphocytic ADJ3 sicknesses) OR (chroniclymphocytic ADJ3 sickness) OR (chroniclymphocytic ADJ3 sickening) OR (chroniclymphocytic ADJ3 sicklefin) OR (chroniclymphocytic ADJ3 sickened) OR (chroniclymphocytic ADJ3 sickling) OR (chroniclymphocytic ADJ3 sickbed) OR (chroniclymphocytic ADJ3 sick) OR (chroniclymphocytic ADJ3 sickly) OR (chroniclymphocytic ADJ3 sicko) OR (chroniclymphocytic ADJ3 sickles) OR (chroniclymphocytic ADJ3 sickel) OR (chroniclymphocytic ADJ3 sickroom) OR (chroniclymphocytic ADJ3 sicks) OR (chroniclymphocytic ADJ3 sickbay) OR (chroniclymphocytic ADJ3 sicktime) OR (chroniclymphocytic ADJ3 sickie) OR (chroniclymphocytic ADJ3 sicknes) OR (chroniclymphocytic ADJ3 sickleave) OR (chroniclymphocytic ADJ3 sickens) OR (chroniclymphocytic ADJ3 sicklaneny) OR (chroniclymphocytic ADJ3 sicklepod) OR (chroniclymphocytic ADJ3 sickliness) OR (chroniclymphocytic ADJ3 sickenss) OR (chroniclymphocytic ADJ3 sicklemia) OR (long-term ADJ3 disease) OR (long-term ADJ3 diseases) OR (long-term ADJ3 diseased) OR (long-term ADJ3 diseasefree) OR (long-term ADJ3 disease0) OR (long-term ADJ3 diseasewas) OR (long-term ADJ3 diseasein) OR (long-term ADJ3 diseaserenin) OR (long-term ADJ3 diseaserelated) OR (long-term ADJ3 diseasemodifying) OR (long-term ADJ3 diseasemeth) OR (long-term ADJ3 diseaseavancando) OR (long-term ADJ3 diseaseprevention) OR (long-term ADJ3 disease19) OR (long-term ADJ3 diseases1) OR (long-term ADJ3 diseasehepatitis) OR (long-term ADJ3 diseaseses) OR (long-term ADJ3 diseasebefore) OR (long-term ADJ3 diseasesthe) OR (long-term ADJ3 diseasesincluding) OR (long-term ADJ3 diseasest) OR (long-term ADJ3 diseaseprogression) OR (long-term ADJ3 diseaseci) OR (long-term ADJ3 diseasedjoint) OR (long-term ADJ3 diseaseand) OR (long-term ADJ3 diseasewith) OR (long-term ADJ3 diseaseful) OR (long-term ADJ3 diseasemanagement) OR (long-term ADJ3 diseasefor) OR (long-term ADJ3 diseasepatients) OR (long-term ADJ3 diseasethat) OR (long-term ADJ3 conditions) OR (long-term ADJ3 conditionally) OR (long-term ADJ3 condition) OR (long-term ADJ3 conditioner) OR (long-term ADJ3 conditioners) OR (long-term ADJ3 conditional) OR (long-term ADJ3 conditioning) OR (long-term ADJ3 conditionality) OR (long-term ADJ3 conditioned) OR (long-term ADJ3 conditionings) OR (long-term ADJ3 conditionalities) OR (long-term ADJ3 conditionals) OR (long-term ADJ3 conditionallyhealthy) OR (long-term ADJ3 conditionalization) OR (long-term ADJ3 conditionstrial) OR (long-term ADJ3 conditionsin) OR (long-term ADJ3 conditionsns) OR (long-term ADJ3 conditionsby) OR (long-term ADJ3 conditionning) OR (long-term ADJ3 conditioningcoaches) OR (long-term ADJ3 conditionining) OR (long-term ADJ3 conditionsthe) OR (long-term ADJ3 conditioncategory) OR (long-term ADJ3 conditionsglass) OR (long-term ADJ3 conditionized) OR (long-term ADJ3 conditionstrained) OR (long-term ADJ3 conditions4) OR (long-term ADJ3 conditionsto) OR (long-term ADJ3 conditionsfor) OR (long-term ADJ3 conditionn) OR (long-term ADJ3 conditionary) OR (long-term ADJ3 conditionability) OR (long-term ADJ3 conditiong) OR (long-term ADJ3 illness) OR (long-term ADJ3 illnesses) OR (long-term ADJ3 illnesspmh) OR (long-term ADJ3 illnessnwb) OR (long-term ADJ3 illnessnew) OR (long-term ADJ3 disorder) OR (long-term ADJ3 disorders) OR (long-term ADJ3 disorderly) OR (long-term ADJ3 disordering) OR (long-term ADJ3 disordered) OR (long-term ADJ3 disorderliness) OR (long-term ADJ3 disorderswhich) OR (long-term ADJ3 disorderof) OR (long-term ADJ3 disorderd) OR (long-term ADJ3 disordercould) OR (long-term ADJ3 disorderis) OR (long-term ADJ3 disorderrs) OR (long-term ADJ3 sicker) OR (long-term ADJ3 sickest) OR (long-term ADJ3 sickle) OR (long-term ADJ3 sickkids) OR (long-term ADJ3 sicknesses) OR (long-term ADJ3 sickness) OR (long-term ADJ3 sickening) OR (long-term ADJ3 sicklefin) OR (long-term ADJ3 sickened) OR (long-term ADJ3 sickling) OR (long-term ADJ3 sickbed) OR (long-term ADJ3 sick) OR (long-term ADJ3 sickly) OR (long-term ADJ3 sicko) OR (long-term ADJ3 sickles) OR (long-term ADJ3 sickel) OR (long-term ADJ3 sickroom) OR (long-term ADJ3 sicks) OR (long-term ADJ3 sickbay) OR (long-term ADJ3 sicktime) OR (long-term ADJ3 sickie) OR (long-term ADJ3 sicknes) OR (long-term ADJ3 sickleave) OR (long-term ADJ3 sickens) OR (long-term ADJ3 sicklaneny) OR (long-term ADJ3 sicklepod) OR (long-term ADJ3 sickliness) OR (long-term ADJ3 sickenss) OR (long-term ADJ3 sicklemia) OR (ongoing ADJ3 disease) OR (ongoing ADJ3 diseases) OR (ongoing ADJ3 diseased) OR (ongoing ADJ3 diseasefree) OR (ongoing ADJ3 disease0) OR (ongoing ADJ3 diseasewas) OR (ongoing ADJ3 diseasein) OR (ongoing ADJ3 diseaserenin) OR (ongoing ADJ3 diseaserelated) OR (ongoing ADJ3 diseasemodifying) OR (ongoing ADJ3 diseasemeth) OR (ongoing ADJ3 diseaseavancando) OR (ongoing ADJ3 diseaseprevention) OR (ongoing ADJ3 disease19) OR (ongoing ADJ3 diseases1) OR (ongoing ADJ3 diseasehepatitis) OR (ongoing ADJ3 diseaseses) OR (ongoing ADJ3 diseasebefore) OR (ongoing ADJ3 diseasesthe) OR (ongoing ADJ3 diseasesincluding) OR (ongoing ADJ3 diseasest) OR (ongoing ADJ3 diseaseprogression) OR (ongoing ADJ3 diseaseci) OR (ongoing ADJ3 diseasedjoint) OR (ongoing ADJ3 diseaseand) OR (ongoing ADJ3 diseasewith) OR (ongoing ADJ3 diseaseful) OR (ongoing ADJ3 diseasemanagement) OR (ongoing ADJ3 diseasefor) OR (ongoing ADJ3 diseasepatients) OR (ongoing ADJ3 diseasethat) OR (ongoing ADJ3 conditions) OR (ongoing ADJ3 conditionally) OR (ongoing ADJ3 condition) OR (ongoing ADJ3 conditioner) OR (ongoing ADJ3 conditioners) OR (ongoing ADJ3 conditional) OR (ongoing ADJ3 conditioning) OR (ongoing ADJ3 conditionality) OR (ongoing ADJ3 conditioned) OR (ongoing ADJ3 conditionings) OR (ongoing ADJ3 conditionalities) OR (ongoing ADJ3 conditionals) OR (ongoing ADJ3 conditionallyhealthy) OR (ongoing ADJ3 conditionalization) OR (ongoing ADJ3 conditionstrial) OR (ongoing ADJ3 conditionsin) OR (ongoing ADJ3 conditionsns) OR (ongoing ADJ3 conditionsby) OR (ongoing ADJ3 conditionning) OR (ongoing ADJ3 conditioningcoaches) OR (ongoing ADJ3 conditionining) OR (ongoing ADJ3 conditionsthe) OR (ongoing ADJ3 conditioncategory) OR (ongoing ADJ3 conditionsglass) OR (ongoing ADJ3 conditionized) OR (ongoing ADJ3 conditionstrained) OR (ongoing ADJ3 conditions4) OR (ongoing ADJ3 conditionsto) OR (ongoing ADJ3 conditionsfor) OR (ongoing ADJ3 conditionn) OR (ongoing ADJ3 conditionary) OR (ongoing ADJ3 conditionability) OR (ongoing ADJ3 conditiong) OR (ongoing ADJ3 illness) OR (ongoing ADJ3 illnesses) OR (ongoing ADJ3 illnesspmh) OR (ongoing ADJ3 illnessnwb) OR (ongoing ADJ3 illnessnew) OR (ongoing ADJ3 disorder) OR (ongoing ADJ3 disorders) OR (ongoing ADJ3 disorderly) OR (ongoing ADJ3 disordering) OR (ongoing ADJ3 disordered) OR (ongoing ADJ3 disorderliness) OR (ongoing ADJ3 disorderswhich) OR (ongoing ADJ3 disorderof) OR (ongoing ADJ3 disorderd) OR (ongoing ADJ3 disordercould) OR (ongoing ADJ3 disorderis) OR (ongoing ADJ3 disorderrs) OR (ongoing ADJ3 sicker) OR (ongoing ADJ3 sickest) OR (ongoing ADJ3 sickle) OR (ongoing ADJ3 sickkids) OR (ongoing ADJ3 sicknesses) OR (ongoing ADJ3 sickness) OR (ongoing ADJ3 sickening) OR (ongoing ADJ3 sicklefin) OR (ongoing ADJ3 sickened) OR (ongoing ADJ3 sickling) OR (ongoing ADJ3 sickbed) OR (ongoing ADJ3 sick) OR (ongoing ADJ3 sickly) OR (ongoing ADJ3 sicko) OR (ongoing ADJ3 sickles) OR (ongoing ADJ3 sickel) OR (ongoing ADJ3 sickroom) OR (ongoing ADJ3 sicks) OR (ongoing ADJ3 sickbay) OR (ongoing ADJ3 sicktime) OR (ongoing ADJ3 sickie) OR (ongoing ADJ3 sicknes) OR (ongoing ADJ3 sickleave) OR (ongoing ADJ3 sickens) OR (ongoing ADJ3 sicklaneny) OR (ongoing ADJ3 sicklepod) OR (ongoing ADJ3 sickliness) OR (ongoing ADJ3 sickenss) OR (ongoing ADJ3 sicklemia) OR (continuous ADJ3 disease) OR (continuous ADJ3 diseases) OR (continuous ADJ3 diseased) OR (continuous ADJ3 diseasefree) OR (continuous ADJ3 disease0) OR (continuous ADJ3 diseasewas) OR (continuous ADJ3 diseasein) OR (continuous ADJ3 diseaserenin) OR (continuous ADJ3 diseaserelated) OR (continuous ADJ3 diseasemodifying) OR (continuous ADJ3 diseasemeth) OR (continuous ADJ3 diseaseavancando) OR (continuous ADJ3 diseaseprevention) OR (continuous ADJ3 disease19) OR (continuous ADJ3 diseases1) OR (continuous ADJ3 diseasehepatitis) OR (continuous ADJ3 diseaseses) OR (continuous ADJ3 diseasebefore) OR (continuous ADJ3 diseasesthe) OR (continuous ADJ3 diseasesincluding) OR (continuous ADJ3 diseasest) OR (continuous ADJ3 diseaseprogression) OR (continuous ADJ3 diseaseci) OR (continuous ADJ3 diseasedjoint) OR (continuous ADJ3 diseaseand) OR (continuous ADJ3 diseasewith) OR (continuous ADJ3 diseaseful) OR (continuous ADJ3 diseasemanagement) OR (continuous ADJ3 diseasefor) OR (continuous ADJ3 diseasepatients) OR (continuous ADJ3 diseasethat) OR (continuous ADJ3 conditions) OR (continuous ADJ3 conditionally) OR (continuous ADJ3 condition) OR (continuous ADJ3 conditioner) OR (continuous ADJ3 conditioners) OR (continuous ADJ3 conditional) OR (continuous ADJ3 conditioning) OR (continuous ADJ3 conditionality) OR (continuous ADJ3 conditioned) OR (continuous ADJ3 conditionings) OR (continuous ADJ3 conditionalities) OR (continuous ADJ3 conditionals) OR (continuous ADJ3 conditionallyhealthy) OR (continuous ADJ3 conditionalization) OR (continuous ADJ3 conditionstrial) OR (continuous ADJ3 conditionsin) OR (continuous ADJ3 conditionsns) OR (continuous ADJ3 conditionsby) OR (continuous ADJ3 conditionning) OR (continuous ADJ3 conditioningcoaches) OR (continuous ADJ3 conditionining) OR (continuous ADJ3 conditionsthe) OR (continuous ADJ3 conditioncategory) OR (continuous ADJ3 conditionsglass) OR (continuous ADJ3 conditionized) OR (continuous ADJ3 conditionstrained) OR (continuous ADJ3 conditions4) OR (continuous ADJ3 conditionsto) OR (continuous ADJ3 conditionsfor) OR (continuous ADJ3 conditionn) OR (continuous ADJ3 conditionary) OR (continuous ADJ3 conditionability) OR (continuous ADJ3 conditiong) OR (continuous ADJ3 illness) OR (continuous ADJ3 illnesses) OR (continuous ADJ3 illnesspmh) OR (continuous ADJ3 illnessnwb) OR (continuous ADJ3 illnessnew) OR (continuous ADJ3 disorder) OR (continuous ADJ3 disorders) OR (continuous ADJ3 disorderly) OR (continuous ADJ3 disordering) OR (continuous ADJ3 disordered) OR (continuous ADJ3 disorderliness) OR (continuous ADJ3 disorderswhich) OR (continuous ADJ3 disorderof) OR (continuous ADJ3 disorderd) OR (continuous ADJ3 disordercould) OR (continuous ADJ3 disorderis) OR (continuous ADJ3 disorderrs) OR (continuous ADJ3 sicker) OR (continuous ADJ3 sickest) OR (continuous ADJ3 sickle) OR (continuous ADJ3 sickkids) OR (continuous ADJ3 sicknesses) OR (continuous ADJ3 sickness) OR (continuous ADJ3 sickening) OR (continuous ADJ3 sicklefin) OR (continuous ADJ3 sickened) OR (continuous ADJ3 sickling) OR (continuous ADJ3 sickbed) OR (continuous ADJ3 sick) OR (continuous ADJ3 sickly) OR (continuous ADJ3 sicko) OR (continuous ADJ3 sickles) OR (continuous ADJ3 sickel) OR (continuous ADJ3 sickroom) OR (continuous ADJ3 sicks) OR (continuous ADJ3 sickbay) OR (continuous ADJ3 sicktime) OR (continuous ADJ3 sickie) OR (continuous ADJ3 sicknes) OR (continuous ADJ3 sickleave) OR (continuous ADJ3 sickens) OR (continuous ADJ3 sicklaneny) OR (continuous ADJ3 sicklepod) OR (continuous ADJ3 sickliness) OR (continuous ADJ3 sickenss) OR (continuous ADJ3 sicklemia) OR (non ADJ3 communicable ADJ3 disease) OR (non ADJ3 communicable ADJ3 diseases) OR (non ADJ3 communicable ADJ3 diseased) OR (non ADJ3 communicable ADJ3 diseasefree) OR (non ADJ3 communicable ADJ3 disease0) OR (non ADJ3 communicable ADJ3 diseasewas) OR (non ADJ3 communicable ADJ3 diseasein) OR (non ADJ3 communicable ADJ3 diseaserenin) OR (non ADJ3 communicable ADJ3 diseaserelated) OR (non ADJ3 communicable ADJ3 diseasemodifying) OR (non ADJ3 communicable ADJ3 diseasemeth) OR (non ADJ3 communicable ADJ3 diseaseavancando) OR (non ADJ3 communicable ADJ3 diseaseprevention) OR (non ADJ3 communicable ADJ3 disease19) OR (non ADJ3 communicable ADJ3 diseases1) OR (non ADJ3 communicable ADJ3 diseasehepatitis) OR (non ADJ3 communicable ADJ3 diseaseses) OR (non ADJ3 communicable ADJ3 diseasebefore) OR (non ADJ3 communicable ADJ3 diseasesthe) OR (non ADJ3 communicable ADJ3 diseasesincluding) OR (non ADJ3 communicable ADJ3 diseasest) OR (non ADJ3 communicable ADJ3 diseaseprogression) OR (non ADJ3 communicable ADJ3 diseaseci) OR (non ADJ3 communicable ADJ3 diseasedjoint) OR (non ADJ3 communicable ADJ3 diseaseand) OR (non ADJ3 communicable ADJ3 diseasewith) OR (non ADJ3 communicable ADJ3 diseaseful) OR (non ADJ3 communicable ADJ3 diseasemanagement) OR (non ADJ3 communicable ADJ3 diseasefor) OR (non ADJ3 communicable ADJ3 diseasepatients) OR (non ADJ3 communicable ADJ3 diseasethat) OR (non ADJ3 communicable ADJ3 conditions) OR (non ADJ3 communicable ADJ3 conditionally) OR (non ADJ3 communicable ADJ3 condition) OR (non ADJ3 communicable ADJ3 conditioner) OR (non ADJ3 communicable ADJ3 conditioners) OR (non ADJ3 communicable ADJ3 conditional) OR (non ADJ3 communicable ADJ3 conditioning) OR (non ADJ3 communicable ADJ3 conditionality) OR (non ADJ3 communicable ADJ3 conditioned) OR (non ADJ3 communicable ADJ3 conditionings) OR (non ADJ3 communicable ADJ3 conditionalities) OR (non ADJ3 communicable ADJ3 conditionals) OR (non ADJ3 communicable ADJ3 conditionallyhealthy) OR (non ADJ3 communicable ADJ3 conditionalization) OR (non ADJ3 communicable ADJ3 conditionstrial) OR (non ADJ3 communicable ADJ3 conditionsin) OR (non ADJ3 communicable ADJ3 conditionsns) OR (non ADJ3 communicable ADJ3 conditionsby) OR (non ADJ3 communicable ADJ3 conditionning) OR (non ADJ3 communicable ADJ3 conditioningcoaches) OR (non ADJ3 communicable ADJ3 conditionining) OR (non ADJ3 communicable ADJ3 conditionsthe) OR (non ADJ3 communicable ADJ3 conditioncategory) OR (non ADJ3 communicable ADJ3 conditionsglass) OR (non ADJ3 communicable ADJ3 conditionized) OR (non ADJ3 communicable ADJ3 conditionstrained) OR (non ADJ3 communicable ADJ3 conditions4) OR (non ADJ3 communicable ADJ3 conditionsto) OR (non ADJ3 communicable ADJ3 conditionsfor) OR (non ADJ3 communicable ADJ3 conditionn) OR (non ADJ3 communicable ADJ3 conditionary) OR (non ADJ3 communicable ADJ3 conditionability) OR (non ADJ3 communicable ADJ3 conditiong) OR (non ADJ3 communicable ADJ3 illness) OR (non ADJ3 communicable ADJ3 illnesses) OR (non ADJ3 communicable ADJ3 illnesspmh) OR (non ADJ3 communicable ADJ3 illnessnwb) OR (non ADJ3 communicable ADJ3 illnessnew) OR (non ADJ3 communicable ADJ3 disorder) OR (non ADJ3 communicable ADJ3 disorders) OR (non ADJ3 communicable ADJ3 disorderly) OR (non ADJ3 communicable ADJ3 disordering) OR (non ADJ3 communicable ADJ3 disordered) OR (non ADJ3 communicable ADJ3 disorderliness) OR (non ADJ3 communicable ADJ3 disorderswhich) OR (non ADJ3 communicable ADJ3 disorderof) OR (non ADJ3 communicable ADJ3 disorderd) OR (non ADJ3 communicable ADJ3 disordercould) OR (non ADJ3 communicable ADJ3 disorderis) OR (non ADJ3 communicable ADJ3 disorderrs) OR (non ADJ3 communicable ADJ3 sicker) OR (non ADJ3 communicable ADJ3 sickest) OR (non ADJ3 communicable ADJ3 sickle) OR (non ADJ3 communicable ADJ3 sickkids) OR (non ADJ3 communicable ADJ3 sicknesses) OR (non ADJ3 communicable ADJ3 sickness) OR (non ADJ3 communicable ADJ3 sickening) OR (non ADJ3 communicable ADJ3 sicklefin) OR (non ADJ3 communicable ADJ3 sickened) OR (non ADJ3 communicable ADJ3 sickling) OR (non ADJ3 communicable ADJ3 sickbed) OR (non ADJ3 communicable ADJ3 sick) OR (non ADJ3 communicable ADJ3 sickly) OR (non ADJ3 communicable ADJ3 sicko) OR (non ADJ3 communicable ADJ3 sickles) OR (non ADJ3 communicable ADJ3 sickel) OR (non ADJ3 communicable ADJ3 sickroom) OR (non ADJ3 communicable ADJ3 sicks) OR (non ADJ3 communicable ADJ3 sickbay) OR (non ADJ3 communicable ADJ3 sicktime) OR (non ADJ3 communicable ADJ3 sickie) OR (non ADJ3 communicable ADJ3 sicknes) OR (non ADJ3 communicable ADJ3 sickleave) OR (non ADJ3 communicable ADJ3 sickens) OR (non ADJ3 communicable ADJ3 sicklaneny) OR (non ADJ3 communicable ADJ3 sicklepod) OR (non ADJ3 communicable ADJ3 sickliness) OR (non ADJ3 communicable ADJ3 sickenss) OR (non ADJ3 communicable ADJ3 sicklemia) OR (noncommunicable ADJ3 disease) OR (noncommunicable ADJ3 diseases) OR (noncommunicable ADJ3 diseased) OR (noncommunicable ADJ3 diseasefree) OR (noncommunicable ADJ3 disease0) OR (noncommunicable ADJ3 diseasewas) OR (noncommunicable ADJ3 diseasein) OR (noncommunicable ADJ3 diseaserenin) OR (noncommunicable ADJ3 diseaserelated) OR (noncommunicable ADJ3 diseasemodifying) OR (noncommunicable ADJ3 diseasemeth) OR (noncommunicable ADJ3 diseaseavancando) OR (noncommunicable ADJ3 diseaseprevention) OR (noncommunicable ADJ3 disease19) OR (noncommunicable ADJ3 diseases1) OR (noncommunicable ADJ3 diseasehepatitis) OR (noncommunicable ADJ3 diseaseses) OR (noncommunicable ADJ3 diseasebefore) OR (noncommunicable ADJ3 diseasesthe) OR (noncommunicable ADJ3 diseasesincluding) OR (noncommunicable ADJ3 diseasest) OR (noncommunicable ADJ3 diseaseprogression) OR (noncommunicable ADJ3 diseaseci) OR (noncommunicable ADJ3 diseasedjoint) OR (noncommunicable ADJ3 diseaseand) OR (noncommunicable ADJ3 diseasewith) OR (noncommunicable ADJ3 diseaseful) OR (noncommunicable ADJ3 diseasemanagement) OR (noncommunicable ADJ3 diseasefor) OR (noncommunicable ADJ3 diseasepatients) OR (noncommunicable ADJ3 diseasethat) OR (noncommunicable ADJ3 conditions) OR (noncommunicable ADJ3 conditionally) OR (noncommunicable ADJ3 condition) OR (noncommunicable ADJ3 conditioner) OR (noncommunicable ADJ3 conditioners) OR (noncommunicable ADJ3 conditional) OR (noncommunicable ADJ3 conditioning) OR (noncommunicable ADJ3 conditionality) OR (noncommunicable ADJ3 conditioned) OR (noncommunicable ADJ3 conditionings) OR (noncommunicable ADJ3 conditionalities) OR (noncommunicable ADJ3 conditionals) OR (noncommunicable ADJ3 conditionallyhealthy) OR (noncommunicable ADJ3 conditionalization) OR (noncommunicable ADJ3 conditionstrial) OR (noncommunicable ADJ3 conditionsin) OR (noncommunicable ADJ3 conditionsns) OR (noncommunicable ADJ3 conditionsby) OR (noncommunicable ADJ3 conditionning) OR (noncommunicable ADJ3 conditioningcoaches) OR (noncommunicable ADJ3 conditionining) OR (noncommunicable ADJ3 conditionsthe) OR (noncommunicable ADJ3 conditioncategory) OR (noncommunicable ADJ3 conditionsglass) OR (noncommunicable ADJ3 conditionized) OR (noncommunicable ADJ3 conditionstrained) OR (noncommunicable ADJ3 conditions4) OR (noncommunicable ADJ3 conditionsto) OR (noncommunicable ADJ3 conditionsfor) OR (noncommunicable ADJ3 conditionn) OR (noncommunicable ADJ3 conditionary) OR (noncommunicable ADJ3 conditionability) OR (noncommunicable ADJ3 conditiong) OR (noncommunicable ADJ3 illness) OR (noncommunicable ADJ3 illnesses) OR (noncommunicable ADJ3 illnesspmh) OR (noncommunicable ADJ3 illnessnwb) OR (noncommunicable ADJ3 illnessnew) OR (noncommunicable ADJ3 disorder) OR (noncommunicable ADJ3 disorders) OR (noncommunicable ADJ3 disorderly) OR (noncommunicable ADJ3 disordering) OR (noncommunicable ADJ3 disordered) OR (noncommunicable ADJ3 disorderliness) OR (noncommunicable ADJ3 disorderswhich) OR (noncommunicable ADJ3 disorderof) OR (noncommunicable ADJ3 disorderd) OR (noncommunicable ADJ3 disordercould) OR (noncommunicable ADJ3 disorderis) OR (noncommunicable ADJ3 disorderrs) OR (noncommunicable ADJ3 sicker) OR (noncommunicable ADJ3 sickest) OR (noncommunicable ADJ3 sickle) OR (noncommunicable ADJ3 sickkids) OR (noncommunicable ADJ3 sicknesses) OR (noncommunicable ADJ3 sickness) OR (noncommunicable ADJ3 sickening) OR (noncommunicable ADJ3 sicklefin) OR (noncommunicable ADJ3 sickened) OR (noncommunicable ADJ3 sickling) OR (noncommunicable ADJ3 sickbed) OR (noncommunicable ADJ3 sick) OR (noncommunicable ADJ3 sickly) OR (noncommunicable ADJ3 sicko) OR (noncommunicable ADJ3 sickles) OR (noncommunicable ADJ3 sickel) OR (noncommunicable ADJ3 sickroom) OR (noncommunicable ADJ3 sicks) OR (noncommunicable ADJ3 sickbay) OR (noncommunicable ADJ3 sicktime) OR (noncommunicable ADJ3 sickie) OR (noncommunicable ADJ3 sicknes) OR (noncommunicable ADJ3 sickleave) OR (noncommunicable ADJ3 sickens) OR (noncommunicable ADJ3 sicklaneny) OR (noncommunicable ADJ3 sicklepod) OR (noncommunicable ADJ3 sickliness) OR (noncommunicable ADJ3 sickenss) OR (noncommunicable ADJ3 sicklemia) OR (persistent ADJ3 disease) OR (persistent ADJ3 diseases) OR (persistent ADJ3 diseased) OR (persistent ADJ3 diseasefree) OR (persistent ADJ3 disease0) OR (persistent ADJ3 diseasewas) OR (persistent ADJ3 diseasein) OR (persistent ADJ3 diseaserenin) OR (persistent ADJ3 diseaserelated) OR (persistent ADJ3 diseasemodifying) OR (persistent ADJ3 diseasemeth) OR (persistent ADJ3 diseaseavancando) OR (persistent ADJ3 diseaseprevention) OR (persistent ADJ3 disease19) OR (persistent ADJ3 diseases1) OR (persistent ADJ3 diseasehepatitis) OR (persistent ADJ3 diseaseses) OR (persistent ADJ3 diseasebefore) OR (persistent ADJ3 diseasesthe) OR (persistent ADJ3 diseasesincluding) OR (persistent ADJ3 diseasest) OR (persistent ADJ3 diseaseprogression) OR (persistent ADJ3 diseaseci) OR (persistent ADJ3 diseasedjoint) OR (persistent ADJ3 diseaseand) OR (persistent ADJ3 diseasewith) OR (persistent ADJ3 diseaseful) OR (persistent ADJ3 diseasemanagement) OR (persistent ADJ3 diseasefor) OR (persistent ADJ3 diseasepatients) OR (persistent ADJ3 diseasethat) OR (persistent ADJ3 conditions) OR (persistent ADJ3 conditionally) OR (persistent ADJ3 condition) OR (persistent ADJ3 conditioner) OR (persistent ADJ3 conditioners) OR (persistent ADJ3 conditional) OR (persistent ADJ3 conditioning) OR (persistent ADJ3 conditionality) OR (persistent ADJ3 conditioned) OR (persistent ADJ3 conditionings) OR (persistent ADJ3 conditionalities) OR (persistent ADJ3 conditionals) OR (persistent ADJ3 conditionallyhealthy) OR (persistent ADJ3 conditionalization) OR (persistent ADJ3 conditionstrial) OR (persistent ADJ3 conditionsin) OR (persistent ADJ3 conditionsns) OR (persistent ADJ3 conditionsby) OR (persistent ADJ3 conditionning) OR (persistent ADJ3 conditioningcoaches) OR (persistent ADJ3 conditionining) OR (persistent ADJ3 conditionsthe) OR (persistent ADJ3 conditioncategory) OR (persistent ADJ3 conditionsglass) OR (persistent ADJ3 conditionized) OR (persistent ADJ3 conditionstrained) OR (persistent ADJ3 conditions4) OR (persistent ADJ3 conditionsto) OR (persistent ADJ3 conditionsfor) OR (persistent ADJ3 conditionn) OR (persistent ADJ3 conditionary) OR (persistent ADJ3 conditionability) OR (persistent ADJ3 conditiong) OR (persistent ADJ3 illness) OR (persistent ADJ3 illnesses) OR (persistent ADJ3 illnesspmh) OR (persistent ADJ3 illnessnwb) OR (persistent ADJ3 illnessnew) OR (persistent ADJ3 disorder) OR (persistent ADJ3 disorders) OR (persistent ADJ3 disorderly) OR (persistent ADJ3 disordering) OR (persistent ADJ3 disordered) OR (persistent ADJ3 disorderliness) OR (persistent ADJ3 disorderswhich) OR (persistent ADJ3 disorderof) OR (persistent ADJ3 disorderd) OR (persistent ADJ3 disordercould) OR (persistent ADJ3 disorderis) OR (persistent ADJ3 disorderrs) OR (persistent ADJ3 sicker) OR (persistent ADJ3 sickest) OR (persistent ADJ3 sickle) OR (persistent ADJ3 sickkids) OR (persistent ADJ3 sicknesses) OR (persistent ADJ3 sickness) OR (persistent ADJ3 sickening) OR (persistent ADJ3 sicklefin) OR (persistent ADJ3 sickened) OR (persistent ADJ3 sickling) OR (persistent ADJ3 sickbed) OR (persistent ADJ3 sick) OR (persistent ADJ3 sickly) OR (persistent ADJ3 sicko) OR (persistent ADJ3 sickles) OR (persistent ADJ3 sickel) OR (persistent ADJ3 sickroom) OR (persistent ADJ3 sicks) OR (persistent ADJ3 sickbay) OR (persistent ADJ3 sicktime) OR (persistent ADJ3 sickie) OR (persistent ADJ3 sicknes) OR (persistent ADJ3 sickleave) OR (persistent ADJ3 sickens) OR (persistent ADJ3 sicklaneny) OR (persistent ADJ3 sicklepod) OR (persistent ADJ3 sickliness) OR (persistent ADJ3 sickenss) OR (persistent ADJ3 sicklemia) OR (enduring ADJ3 disease) OR (enduring ADJ3 diseases) OR (enduring ADJ3 diseased) OR (enduring ADJ3 diseasefree) OR (enduring ADJ3 disease0) OR (enduring ADJ3 diseasewas) OR (enduring ADJ3 diseasein) OR (enduring ADJ3 diseaserenin) OR (enduring ADJ3 diseaserelated) OR (enduring ADJ3 diseasemodifying) OR (enduring ADJ3 diseasemeth) OR (enduring ADJ3 diseaseavancando) OR (enduring ADJ3 diseaseprevention) OR (enduring ADJ3 disease19) OR (enduring ADJ3 diseases1) OR (enduring ADJ3 diseasehepatitis) OR (enduring ADJ3 diseaseses) OR (enduring ADJ3 diseasebefore) OR (enduring ADJ3 diseasesthe) OR (enduring ADJ3 diseasesincluding) OR (enduring ADJ3 diseasest) OR (enduring ADJ3 diseaseprogression) OR (enduring ADJ3 diseaseci) OR (enduring ADJ3 diseasedjoint) OR (enduring ADJ3 diseaseand) OR (enduring ADJ3 diseasewith) OR (enduring ADJ3 diseaseful) OR (enduring ADJ3 diseasemanagement) OR (enduring ADJ3 diseasefor) OR (enduring ADJ3 diseasepatients) OR (enduring ADJ3 diseasethat) OR (enduring ADJ3 conditions) OR (enduring ADJ3 conditionally) OR (enduring ADJ3 condition) OR (enduring ADJ3 conditioner) OR (enduring ADJ3 conditioners) OR (enduring ADJ3 conditional) OR (enduring ADJ3 conditioning) OR (enduring ADJ3 conditionality) OR (enduring ADJ3 conditioned) OR (enduring ADJ3 conditionings) OR (enduring ADJ3 conditionalities) OR (enduring ADJ3 conditionals) OR (enduring ADJ3 conditionallyhealthy) OR (enduring ADJ3 conditionalization) OR (enduring ADJ3 conditionstrial) OR (enduring ADJ3 conditionsin) OR (enduring ADJ3 conditionsns) OR (enduring ADJ3 conditionsby) OR (enduring ADJ3 conditionning) OR (enduring ADJ3 conditioningcoaches) OR (enduring ADJ3 conditionining) OR (enduring ADJ3 conditionsthe) OR (enduring ADJ3 conditioncategory) OR (enduring ADJ3 conditionsglass) OR (enduring ADJ3 conditionized) OR (enduring ADJ3 conditionstrained) OR (enduring ADJ3 conditions4) OR (enduring ADJ3 conditionsto) OR (enduring ADJ3 conditionsfor) OR (enduring ADJ3 conditionn) OR (enduring ADJ3 conditionary) OR (enduring ADJ3 conditionability) OR (enduring ADJ3 conditiong) OR (enduring ADJ3 illness) OR (enduring ADJ3 illnesses) OR (enduring ADJ3 illnesspmh) OR (enduring ADJ3 illnessnwb) OR (enduring ADJ3 illnessnew) OR (enduring ADJ3 disorder) OR (enduring ADJ3 disorders) OR (enduring ADJ3 disorderly) OR (enduring ADJ3 disordering) OR (enduring ADJ3 disordered) OR (enduring ADJ3 disorderliness) OR (enduring ADJ3 disorderswhich) OR (enduring ADJ3 disorderof) OR (enduring ADJ3 disorderd) OR (enduring ADJ3 disordercould) OR (enduring ADJ3 disorderis) OR (enduring ADJ3 disorderrs) OR (enduring ADJ3 sicker) OR (enduring ADJ3 sickest) OR (enduring ADJ3 sickle) OR (enduring ADJ3 sickkids) OR (enduring ADJ3 sicknesses) OR (enduring ADJ3 sickness) OR (enduring ADJ3 sickening) OR (enduring ADJ3 sicklefin) OR (enduring ADJ3 sickened) OR (enduring ADJ3 sickling) OR (enduring ADJ3 sickbed) OR (enduring ADJ3 sick) OR (enduring ADJ3 sickly) OR (enduring ADJ3 sicko) OR (enduring ADJ3 sickles) OR (enduring ADJ3 sickel) OR (enduring ADJ3 sickroom) OR (enduring ADJ3 sicks) OR (enduring ADJ3 sickbay) OR (enduring ADJ3 sicktime) OR (enduring ADJ3 sickie) OR (enduring ADJ3 sicknes) OR (enduring ADJ3 sickleave) OR (enduring ADJ3 sickens) OR (enduring ADJ3 sicklaneny) OR (enduring ADJ3 sicklepod) OR (enduring ADJ3 sickliness) OR (enduring ADJ3 sickenss) OR (enduring ADJ3 sicklemia))).ti,ab,kf.) AND (exp "Patient participation"/ or exp "Self-management"/ or (("Self manag*" or "Self care" or “Person centered care” or ((patient ADJ3 empowerment) OR (patient ADJ3 empowering) OR (patient ADJ3 empower) OR (patient ADJ3 empowered) OR (patient ADJ3 empowers) OR (patient ADJ3 empowerd) OR (patient ADJ3 empowermen) OR (patient ADJ3 empowerer) OR (patient ADJ3 empowerments) OR (patient ADJ3 empowerers) OR (patient ADJ3 empowerstats) OR (patient ADJ3 empowerclinicians) OR (patient ADJ3 empowerstat) OR (patient ADJ3 empowerrf) OR (patient ADJ3 empowerplus) OR (patient ADJ3 empowernment) OR (patients ADJ3 empowerment) OR (patients ADJ3 empowering) OR (patients ADJ3 empower) OR (patients ADJ3 empowered) OR (patients ADJ3 empowers) OR (patients ADJ3 empowerd) OR (patients ADJ3 empowermen) OR (patients ADJ3 empowerer) OR (patients ADJ3 empowerments) OR (patients ADJ3 empowerers) OR (patients ADJ3 empowerstats) OR (patients ADJ3 empowerclinicians) OR (patients ADJ3 empowerstat) OR (patients ADJ3 empowerrf) OR (patients ADJ3 empowerplus) OR (patients ADJ3 empowernment) OR (patientenakte ADJ3 empowerment) OR (patientenakte ADJ3 empowering) OR (patientenakte ADJ3 empower) OR (patientenakte ADJ3 empowered) OR (patientenakte ADJ3 empowers) OR (patientenakte ADJ3 empowerd) OR (patientenakte ADJ3 empowermen) OR (patientenakte ADJ3 empowerer) OR (patientenakte ADJ3 empowerments) OR (patientenakte ADJ3 empowerers) OR (patientenakte ADJ3 empowerstats) OR (patientenakte ADJ3 empowerclinicians) OR (patientenakte ADJ3 empowerstat) OR (patientenakte ADJ3 empowerrf) OR (patientenakte ADJ3 empowerplus) OR (patientenakte ADJ3 empowernment) OR (patientslikeme ADJ3 empowerment) OR (patientslikeme ADJ3 empowering) OR (patientslikeme ADJ3 empower) OR (patientslikeme ADJ3 empowered) OR (patientslikeme ADJ3 empowers) OR (patientslikeme ADJ3 empowerd) OR (patientslikeme ADJ3 empowermen) OR (patientslikeme ADJ3 empowerer) OR (patientslikeme ADJ3 empowerments) OR (patientslikeme ADJ3 empowerers) OR (patientslikeme ADJ3 empowerstats) OR (patientslikeme ADJ3 empowerclinicians) OR (patientslikeme ADJ3 empowerstat) OR (patientslikeme ADJ3 empowerrf) OR (patientslikeme ADJ3 empowerplus) OR (patientslikeme ADJ3 empowernment) OR (patientgpt ADJ3 empowerment) OR (patientgpt ADJ3 empowering) OR (patientgpt ADJ3 empower) OR (patientgpt ADJ3 empowered) OR (patientgpt ADJ3 empowers) OR (patientgpt ADJ3 empowerd) OR (patientgpt ADJ3 empowermen) OR (patientgpt ADJ3 empowerer) OR (patientgpt ADJ3 empowerments) OR (patientgpt ADJ3 empowerers) OR (patientgpt ADJ3 empowerstats) OR (patientgpt ADJ3 empowerclinicians) OR (patientgpt ADJ3 empowerstat) OR (patientgpt ADJ3 empowerrf) OR (patientgpt ADJ3 empowerplus) OR (patientgpt ADJ3 empowernment) OR (patientsand ADJ3 empowerment) OR (patientsand ADJ3 empowering) OR (patientsand ADJ3 empower) OR (patientsand ADJ3 empowered) OR (patientsand ADJ3 empowers) OR (patientsand ADJ3 empowerd) OR (patientsand ADJ3 empowermen) OR (patientsand ADJ3 empowerer) OR (patientsand ADJ3 empowerments) OR (patientsand ADJ3 empowerers) OR (patientsand ADJ3 empowerstats) OR (patientsand ADJ3 empowerclinicians) OR (patientsand ADJ3 empowerstat) OR (patientsand ADJ3 empowerrf) OR (patientsand ADJ3 empowerplus) OR (patientsand ADJ3 empowernment) OR (patienthood ADJ3 empowerment) OR (patienthood ADJ3 empowering) OR (patienthood ADJ3 empower) OR (patienthood ADJ3 empowered) OR (patienthood ADJ3 empowers) OR (patienthood ADJ3 empowerd) OR (patienthood ADJ3 empowermen) OR (patienthood ADJ3 empowerer) OR (patienthood ADJ3 empowerments) OR (patienthood ADJ3 empowerers) OR (patienthood ADJ3 empowerstats) OR (patienthood ADJ3 empowerclinicians) OR (patienthood ADJ3 empowerstat) OR (patienthood ADJ3 empowerrf) OR (patienthood ADJ3 empowerplus) OR (patienthood ADJ3 empowernment) OR (patiently ADJ3 empowerment) OR (patiently ADJ3 empowering) OR (patiently ADJ3 empower) OR (patiently ADJ3 empowered) OR (patiently ADJ3 empowers) OR (patiently ADJ3 empowerd) OR (patiently ADJ3 empowermen) OR (patiently ADJ3 empowerer) OR (patiently ADJ3 empowerments) OR (patiently ADJ3 empowerers) OR (patiently ADJ3 empowerstats) OR (patiently ADJ3 empowerclinicians) OR (patiently ADJ3 empowerstat) OR (patiently ADJ3 empowerrf) OR (patiently ADJ3 empowerplus) OR (patiently ADJ3 empowernment) OR (patientenverfugung ADJ3 empowerment) OR (patientenverfugung ADJ3 empowering) OR (patientenverfugung ADJ3 empower) OR (patientenverfugung ADJ3 empowered) OR (patientenverfugung ADJ3 empowers) OR (patientenverfugung ADJ3 empowerd) OR (patientenverfugung ADJ3 empowermen) OR (patientenverfugung ADJ3 empowerer) OR (patientenverfugung ADJ3 empowerments) OR (patientenverfugung ADJ3 empowerers) OR (patientenverfugung ADJ3 empowerstats) OR (patientenverfugung ADJ3 empowerclinicians) OR (patientenverfugung ADJ3 empowerstat) OR (patientenverfugung ADJ3 empowerrf) OR (patientenverfugung ADJ3 empowerplus) OR (patientenverfugung ADJ3 empowernment) OR (patienten ADJ3 empowerment) OR (patienten ADJ3 empowering) OR (patienten ADJ3 empower) OR (patienten ADJ3 empowered) OR (patienten ADJ3 empowers) OR (patienten ADJ3 empowerd) OR (patienten ADJ3 empowermen) OR (patienten ADJ3 empowerer) OR (patienten ADJ3 empowerments) OR (patienten ADJ3 empowerers) OR (patienten ADJ3 empowerstats) OR (patienten ADJ3 empowerclinicians) OR (patienten ADJ3 empowerstat) OR (patienten ADJ3 empowerrf) OR (patienten ADJ3 empowerplus) OR (patienten ADJ3 empowernment))

).ti,ab,kf. or ((patient ADJ3 activation) OR (patient ADJ3 activating) OR (patient ADJ3 activations) OR (patient ADJ3 activationsthrough) OR (patient ADJ3 activationrelated) OR (patient ADJ3 activational) OR (patient ADJ3 activatibility) OR (patient ADJ3 activative) OR (patient ADJ3 engagement) OR (patient ADJ3 engagements) OR (patient ADJ3 engage) OR (patient ADJ3 engaging) OR (patient ADJ3 engaged) OR (patient ADJ3 engagedmd) OR (patient ADJ3 engages) OR (patient ADJ3 engager) OR (patient ADJ3 engagers) OR (patient ADJ3 engagespark) OR (patient ADJ3 engagment) OR (patient ADJ3 engagemodel) OR (patient ADJ3 engagability) OR (patient ADJ3 engageability) OR (patient ADJ3 engageness) OR (patient ADJ3 engageable) OR (patient ADJ3 engagedness) OR (patient ADJ3 involvement) OR (patient ADJ3 involving) OR (patient ADJ3 involvements) OR (patient ADJ3 involved) OR (patient ADJ3 involves) OR (patient ADJ3 involve) OR (patient ADJ3 involveme) OR (patient ADJ3 involvment) OR (patient ADJ3 involvine) OR (patient ADJ3 involvemet) OR (patient ADJ3 involvingcomplete) OR (patient ADJ3 involving4046) OR (patient ADJ3 involvihttng) OR (patient ADJ3 involvingthe) OR (patient ADJ3 involved240) OR (patient ADJ3 involvementwith) OR (patient ADJ3 involvementconfirmed) OR (patient ADJ3 involvingcontext) OR (patient ADJ3 involvementmethods) OR (patient ADJ3 involvement1) OR (patient ADJ3 involvementa) OR (patient ADJ3 involv) OR (patient ADJ3 involveda) OR (patient ADJ3 involvation) OR (patient ADJ3 involvments) OR (patient ADJ3 involvesregular) OR (patient ADJ3 involvementby) OR (patient ADJ3 involvemtation) OR (patient ADJ3 participation) OR (patient ADJ3 participating) OR (patient ADJ3 participations) OR (patient ADJ3 participative) OR (patient ADJ3 participational) OR (patient ADJ3 contribution) OR (patient ADJ3 contributive) OR (patient ADJ3 contributing) OR (patient ADJ3 contributions) OR (patient ADJ3 contributivity) OR (patient ADJ3 contributionthe) OR (patient ADJ3 contributionelderly) OR (patient ADJ3 contributingto) OR (patient ADJ3 contributively) OR (patient ADJ3 contributiontowards) OR (patient ADJ3 contributional) OR (patient ADJ3 contributiing) OR (patient ADJ3 collaborated) OR (patient ADJ3 collaboration) OR (patient ADJ3 collaboratives) OR (patient ADJ3 collaborator) OR (patient ADJ3 collaborative) OR (patient ADJ3 collaboratory) OR (patient ADJ3 collaborates) OR (patient ADJ3 collaborate) OR (patient ADJ3 collaborations) OR (patient ADJ3 collaboratively) OR (patient ADJ3 collaborativeness) OR (patient ADJ3 collaborators) OR (patient ADJ3 collaborathon) OR (patient ADJ3 collaborativo) OR (patient ADJ3 collaborating) OR (patient ADJ3 collaboratingclinics) OR (patient ADJ3 collaborati) OR (patient ADJ3 collaborationism)).ti,kf.)) AND (exp "cardiovascular diseases"/ or exp "Osteoarthritis"/ or exp "Respiration disorders"/ or exp "respiratory tract diseases"/ or exp "diabetes mellitus"/ or ("cardiovascular*" or "cardiac*" or "heart" or "osteoarth*" or "respirat*” or “pulmon*" or "lung*" or "breathing" or "diabet*" OR DM OR T2DM OR T1DM).ti,ab,kf.)
